# The epidemiology and direct medical costs of diseases associated with human papillomavirus infection among men in Manitoba, Canada

**DOI:** 10.1101/2020.12.02.20242891

**Authors:** Christiaan H. Righolt, Gurpreet Pabla, Salaheddin M. Mahmud

## Abstract

**Background:** There is little information on the economic burden of human papillomavirus-related diseases (HPV-RDs) among men. We used province-wide clinical, administrative and accounting databases to measure the direct medical costs of HPV infections in men in Manitoba (Canada).

**Methods:** We included all males aged 9 years and older with health insurance coverage in Manitoba between January 1997 and December 2016. We identified HPV-RD patient cohorts and matched each patient to HPV-RD-free men. We estimated the net direct medical cost (excess cost of hospitalizations, outpatient visits, and prescription drugs) of patients compared to their matches for anogenital warts (AGWs) and HPV-caused cancers. We adjusted costs to 2017 Canadian dollars. For each condition, we attributed costs to HPV based on the etiological fraction caused by HPV infection.

**Results:** We found that the median net direct medical cost was about $250 for AGW patients and $16,000 for invasive cancer patients. The total cost was about $49 million or $2.6 million per year. Overall, 54%-67% ($26-$33 million) was attributable to HPV infection according different estimates of the attributable fraction. The net annual attributable cost was $2.37-$2.95 per male resident and $161-$200 per male newborn. The estimated potential savings was 30% for the bivalent vaccine and 56%-60% for the quadrivalent and nonavalent vaccines.

**Conclusions:** Overall, HPV’s economic burden on males remains significant, the average cost of treating all conditions attributable to HPV was about $180 per male newborn. Invasive cancer accounted for the majority of these costs.

## 1 Introduction

Anogenital infection with the human papillomavirus (HPV) is the most common sexually transmitted infection in Canada and worldwide [1–3]. Although HPV infection is as prevalent among men [4], the focus of most research in this field has been on women due to its strong association with cervical cancer, a more common and devastating cancer. However, persistent infection with 13 or so high-risk oncogenic HPV genotypes is known to causes most (90%) anal cancers and a significant proportion of oropharyngeal cancers (12%) in both men and women [5–7]. About 40% of penile cancers are also caused by HPV infection. In addition, “low-risk” types 6 and 11 are responsible for most external anogenital warts (AGWs) in both genders [8]. With the availability of prophylactic vaccines against HPV infection in males, it is important to understand both the epidemiologic and economic male burden associated with these human papillomavirus-related diseases (HPV-RDs).

Little is known about the epidemiology and long-term trends of anal and penile cancer among Canadian men. Because of their lower incidence, they are not regularly reported in national and provincial cancer surveillance reports. In addition, patterns of treatment of these cancers have not been characterized at the population level in Canada. Most studies that estimated HPV-related medical costs were not based on nationally representative data as information was often obtained for individual organizations (hospitals, health maintenance organizations, etc.) or from patient surveys [9–11]. Some studies used diseases incidence rates derived from mathematical modeling, these rates were then combined with estimates of average costs per case/service as derived from the literature to arrive at total expected costs [12,13]. Most studies were limited to costs associated with the prevention and treatment of one or two conditions and were focused on the cost of all HPV-RDs. We previously reported on the direct medical cost for both men and women in Manitoba and found that two-thirds of costs were for cervical screening; because of this, we reported limited data specific to males [14].

Information on the economic burden of disease is essential for planning and evaluating prevention and control programs [15]. Up-to-date cost estimates are essential to inform decisions on priorities, evaluate existing prevention and treatment programs, forecast expenditures, and estimate the impact of new interventions [16]. Estimating cost-effectiveness requires detailed knowledge of costs to facilitate decision-making on whether to expand HPV vaccine programs or to switch to using the nonavalent vaccine [17,18].

We aimed to assess the direct medical costs associated with the prevention and treatment of conditions attributable to HPV infection among men in Manitoba, Canada.

## 2 Methods

We used several Manitoba Health (MH) clinical and administrative databases to identify all males diagnosed with an HPV-RD in Manitoba between January 1997 and December 2016 and measure all costs incurred by MH in relation to the diagnosis and treatment of these conditions (*direct medical costs*).

### 2.1 Data sources

MH is the publicly funded health insurance agency providing comprehensive health insurance, including coverage for hospital and outpatient physician services, to the province’s 1.3 million residents. Coverage is universal, with no eligibility distinction based on age or income, and participation rates are very high (>99%) [19]. Insured services include hospital, physician and preventive services including vaccinations. MH maintains several centralized, administrative electronic databases that are linkable using a unique personal health identification number (PHIN). The completeness and accuracy of these databases are well established [20,21]. These databases have been used extensively in studies of disease surveillance and post-marketing evaluation of various vaccines and drugs [22–24].

The Manitoba Health Population Registry (MHPR) tracks addresses and dates of birth, death and insurance coverage for all insured persons. Since 1971, the Hospital Abstracts Database (HAD) recorded virtually all services provided by hospitals in the province, including admissions and day surgeries [20]. The data collected comprise demographic as well as diagnosis and treatment information including primary diagnosis and service or procedure codes, coded using the International Classification of Diseases, Ninth Revision, Clinical Modification (ICD-9-CM) before April, 2004, and the ICD-10-CA (Canadian adaptation of the ICD-10) and the Canadian Classification of Health Interventions (CCI) afterwards. The Medical Services Database (MSD), also in operation since 1971, collects similar information, based on physician fee-for-service or shadow billing, on services provided by physicians in offices, hospitals and outpatient departments across the province [20]. The Drug Program Information Network (DPIN), in operation since 1995, records all prescription drugs dispensed to Manitoba residents, including most personal care home residents [25]. The DPIN database captures data from pharmacy claims for formulary drugs dispensed to all Manitobans even those without prescription drug insurance [25].

CancerCare Manitoba (CCMB) maintains one of the oldest population-based cancer registries in the world (Manitoba Cancer Registry [MCR], in operation since 1956). Reporting of cancer cases is mandated by provincial regulations and required for payments of physicians’ service claims. The MCR is regularly audited by the North American Association of Central Cancer Registries. The quality of cancer registration has been consistently very high [21]. Most cases are pathologically-confirmed (94% for cases registered between 2003 and 2007) and less than 2% of registrations originate from death certificates [21]. It also tracks all cancer-specific treatment using ICD-9 procedure codes prior to 2005 and ICD-10 Canadian Classification of Health Interventions codes since then. The Emergency Department Information System (EDIS) captures all visits to Winnipeg (Manitoba’s main urban center with over half the provincial population and the province’s only tertiary care hospitals) emergency departments since April 1999.

### 2.2 Eligibility and study cohort

All males aged 9 years and older who were registered with MH at any point between January 1997 and December 2016 (the *study period*) were eligible for inclusion in this study. Males joined the population cohort on the latest of their 9^th^ birthdate, date of immigration to Manitoba or January 1, 1997. They exited the population cohort on the earliest of December 31, 2016 or the date of loss of MH coverage for any reason, including death or emigration.

To build the study (HPV-RD) cohort, we identified all eligible males who were diagnosed with an incident HPV-RD during the study period. We matched each HPV-RD patient on age (±1 year), length of MH coverage (±1 year) and region of residence to up to 3 eligible males who met all the inclusion criteria but did not have a HPV-RD at the diagnosis date of the case.

We identified males diagnosed with invasive cancer or carcinoma *in situ* of the penis, anus, oral cavity, or oropharynx by linking with the MCR. We used ICD-O-3 codes to identify these cases (Supplementary Table 1). For carcinoma *in situ* (CIS) and cancer patients, the index date was the date of diagnosis as reported in the MCR (defined as the earliest of symptom onset, laboratory diagnosis or cancer treatment date).

We obtained information on incident medically-attended AGWs from the HAD and MSD using previously validated algorithms [26]. In short, persons treated for AGWs were identified from the MSD using tariff codes specific to condyloma (Supplementary Table 2) or from the HAD using period-dependent diagnostic and procedure codes (Supplementary Table 3-4). Because patients can have multiple diagnoses of AGWs, each episode of care had to be separated by 12 months without any related claims to be considered an incident episode (Supplementary Table 6).

We added direct medical costs and the number of healthcare encounters for disease-specific periods: from 180 days prior to the diagnosis date to 730 days after diagnosis for invasive cancer, and from the diagnosis date to 365 days after diagnosis for AGWs and CIS.

### 2.3 Costs and healthcare utilization

We summed the costs, obtained from the administrative databases, of all prescription drugs (including dispensing fees), in-hospital and outpatient diagnostic and therapeutic procedures, and physician visits for each HPV-RD case and their matches. We attributed all costs to the year of diagnosis of the case and indexed costs to 2017 Canadian dollars using the all-item and prescription drugs consumer price indices. We counted all hospitalizations, physician visits, emergency department visits, and dispensed prescriptions for each HPV-RD case and their matches. We attributed all healthcare utilization to the year of diagnosis of the case.

### 2.5 Analysis

We calculated crude and age-standardized annual incidence rates of each HPV-RD using the 2016 Canada census population as the standard population. We calculated incidence rates, age-standardized when appropriate, for different subsets of the population, defined by age group, region of residence, and year of diagnosis.

We compared costs for each HPV-RD patient with their matches. We reported individual net costs (the difference in cost between the case and their specific matches) for all HPV-RDs and for each HPV-RD separately for the entire population and for specific age groups, regions of residence and years of diagnosis. To estimate the net cost attributable to HPV infection, we repeated all analyses after multiplying the cost differential of each HPV-RD by the HPV attributable fraction of that condition (Supplementary Table 7) using two sets of estimates derived from two detailed systematic reviews [27,28]. To estimate potential savings from various vaccines (bivalent, quadrivalent and nonavalent), we multiplied the cost differential of each HPV-RD by the percentage of that HPV-RD attributable to the HPV types included in the specific vaccine using estimates derived from Saraiya et al. [28] (Supplementary Table 8). We repeated the above methods for each patient’s healthcare utilization.

We used SAS 9.4 (SAS Institute, Cary, North Carolina) and Stata 16 (StataCorp, College Station, Texas) for all analysis. This study was approved by the University of Manitoba Research Ethics Board and by MH’s Health Information Privacy Committee.

## 3 Results

During the study period, AGWs were the mostly commonly treated HPV-RD with an overall rate of 149/100,000 person-years (Table 1). AGW rates were highest among 18-30 year-olds (367/100,000). Invasive cancer of the oropharynx was the most common HPV-related cancer (5.2/100,000), peaking among 51-64 year-olds at 13.4/100,000. The HPV-RD cohorts consisted of 14,784 AGW and 1,236 CIS/invasive cancer patients matched to 43,330 and 2,960 HPV-RD-free men (Table 2). About two-thirds of HPV-RD patients lived in Winnipeg, where incidence was highest in the southern suburbs. Incidence rates varied by age group, birth cohort, region of residence, and year of diagnosis (Supplementary Tables 9-33).

**Table 1.**
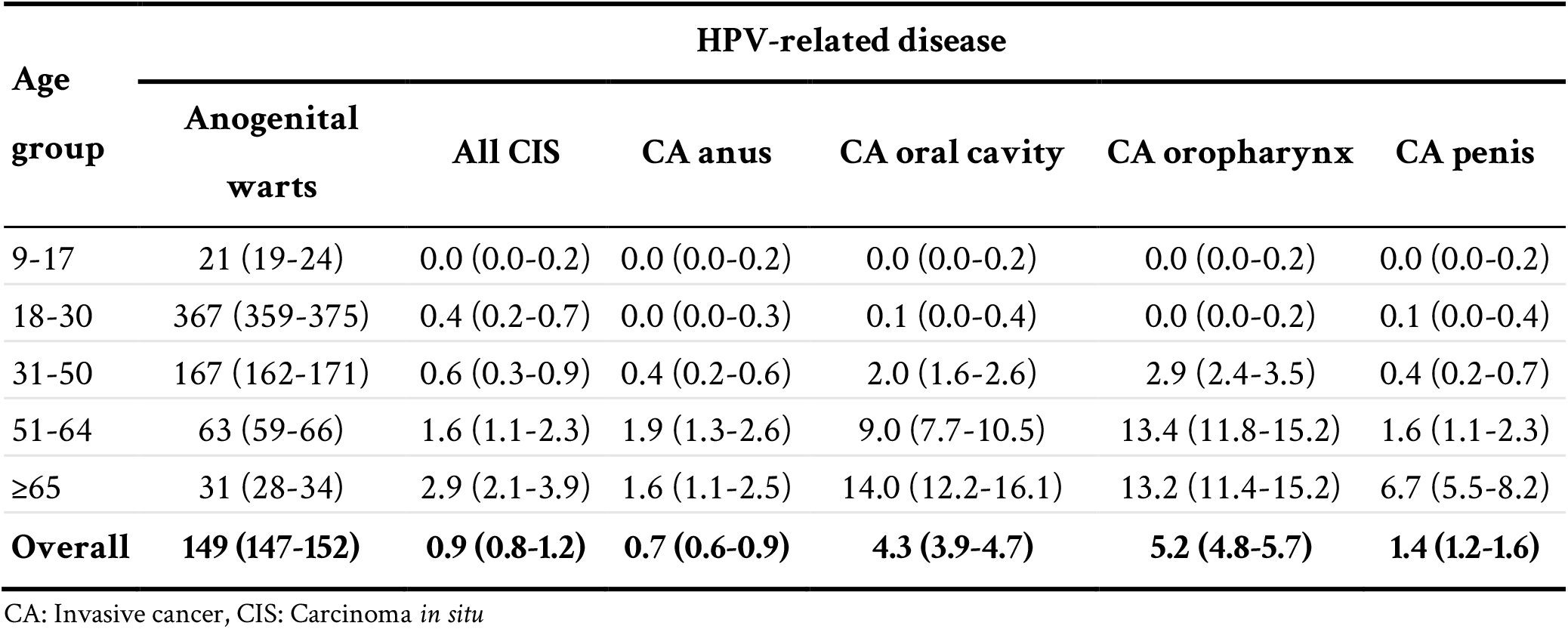
Crude incidence rates (95% confidence interval) per 100,000 person -years of HPV-related diseases among Manitoba males by age group (1997 - 2016).

| Age group | HPV-related disease |  |  |  |  |  |
| --- | --- | --- | --- | --- | --- | --- |
|  | Anogenital warts | All CIS | CA anus | CA oral cavity | CA oropharynx | CA penis |
| 9-17 | 21 (19-24) | 0.0 (0.0-0.2) | 0.0 (0.0-0.2) | 0.0 (0.0-0.2) | 0.0 (0.0-0.2) | 0.0 (0.0-0.2) |
| 18-30 | 367 (359-375) | 0.4 (0.2-0.7) | 0.0 (0.0-0.3) | 0.1 (0.0-0.4) | 0.0 (0.0-0.2) | 0.1 (0.0-0.4) |
| 31-50 | 167 (162-171) | 0.6 (0.3-0.9) | 0.4 (0.2-0.6) | 2.0 (1.6-2.6) | 2.9 (2.4-3.5) | 0.4 (0.2-0.7) |
| 51-64 | 63 (59-66) | 1.6 (1.1-2.3) | 1.9 (1.3-2.6) | 9.0 (7.7-10.5) | 13.4 (11.8-15.2) | 1.6 (1.1-2.3) |
| ≥65 | 31 (28-34) | 2.9 (2.1-3.9) | 1.6 (1.1-2.5) | 14.0 (12.2-16.1) | 13.2 (11.4-15.2) | 6.7 (5.5-8.2) |
| **Overall** | **149 (147-152)** | **0.9 (0.8-1.2)** | **0.7 (0.6-0.9)** | **4.3 (3.9-4.7)** | **5.2 (4.8-5.7)** | **1.4 (1.2-1.6)** |

CA: Invasive cancer, CIS: Carcinoma *in situ*

**Table 2.** Number of HPV-related disease patients and their matches among Manitoba males by certain socio-economic and clinical characteristics

|  | Anogenital warts |  | CIS/Invasive cancer |  |
| --- | --- | --- | --- | --- |
|  | Patients | Matches | Patients | Matches |
|  | (N=14,784) | (N=43,330) | (N=1,236) | (N=2,960) |
| Age (years) |  |  |  |  |
| 9 - 17 | 305 (2.1%) | 904 (2.1%) | 0 (0.0%) | 0 (0.0%) |
| 18 - 30 | 7,589 (51.3%) | 22,265 (51.4%) | 13 (1.1%) | 38 (1.3%) |
| 31 - 50 | 5,409 (36.6%) | 15,936 (36.8%) | 202 (16.3%) | 554 (18.7%) |
| 51 - 64 | 1,107 (7.5%) | 3,231 (7.5%) | 504 (40.8%) | 1,263 (42.7%) |
| ≥ 65 | 374 (2.5%) | 994 (2.3%) | 517 (41.8%) | 1,105 (37.3%) |
| Income quintile |  |  |  |  |
| Q1 (lowest) | 3,086 (20.9%) | 9,111 (21.0%) | 272 (22.0%) | 609 (20.6%) |
| Q2 | 3,160 (21.4%) | 9,234 (21.3%) | 245 (19.8%) | 598 (20.2%) |
| Q3 | 3,133 (21.2%) | 8,760 (20.2%) | 250 (20.2%) | 611 (20.6%) |
| Q4 | 2,707 (18.3%) | 8,015 (18.5%) | 244 (19.7%) | 562 (19.0%) |
| Q5 (highest) | 2,610 (17.7%) | 7,885 (18.2%) | 199 (16.1%) | 533 (18.0%) |
| Unknown | 88 (0.6%) | 325 (0.8%) | 26 (2.1%) | 47 (1.6%) |
| Residence |  |  |  |  |
| Rural | 3,484 (23.6%) | 9,943 (22.9%) | 418 (33.8%) | 944 (31.9%) |
| Urban | 11,271 (76.2%) | 33,317 (76.9%) | 804 (65.0%) | 1,997 (67.5%) |
| Unknown | 29 (0.2%) | 70 (0.2%) | 14 (1.1%) | 19 (0.6%) |
| Regional health authority of residence |  |  |  |  |
| Winnipeg | 10,551 (71.4%) | 31,251 (72.1%) | 753 (60.9%) | 1,879 (63.5%) |
| Interlake-Eastern | 1,025 (6.9%) | 2,955 (6.8%) | 133 (10.8%) | 322 (10.9%) |
| Northern | 376 (2.5%) | 1,023 (2.4%) | 54 (4.4%) | 114 (3.9%) |
| Southern | 1,312 (8.9%) | 3,722 (8.6%) | 110 (8.9%) | 227 (7.7%) |
| Prairie Mountain | 1,491 (10.1%) | 4,309 (9.9%) | 172 (13.9%) | 399 (13.5%) |
| Public Trustee/In CFS care | 29 (0.2%) | 70 (0.2%) | 14 (1.1%) | 19 (0.6%) |
| Winnipeg region of residence |  |  |  |  |
| Northern suburbs | 3,096 (20.9%) | 9,132 (21.1%) | 210 (17.0%) | 515 (17.4%) |
| Inner city | 1,941 (13.1%) | 5,771 (13.3%) | 149 (12.1%) | 380 (12.8%) |
| Southern suburbs | 5,502 (37.2%) | 16,337 (37.7%) | 394 (31.9%) | 984 (33.2%) |
| Non-Winnipeg | 4,216 (28.5%) | 12,020 (27.7%) | 469 (37.9%) | 1,062 (35.9%) |
| Public Trustee/In CFS care | 29 (0.2%) | 70 (0.2%) | 14 (1.1%) | 19 (0.6%) |
| No. of physician visits in the past year |  |  |  |  |
| 0 - 4 | 7,537 (51.0%) | 30,796 (71.1%) | 258 (20.9%) | 1,258 (42.5%) |
| 5 - 11 | 4,767 (32.2%) | 8,606 (19.9%) | 458 (37.1%) | 984 (33.2%) |
| 12+ | 2,480 (16.8%) | 3,928 (9.1%) | 520 (42.1%) | 718 (24.3%) |
| No. of physician visits 2-5 years before the diagnosis date |  |  |  |  |
| 0 - 14 | 8,153 (55.1%) | 30,540 (70.5%) | 478 (38.7%) | 1,222 (41.3%) |
| 15 - 49 | 5,488 (37.1%) | 10,954 (25.3%) | 502 (40.6%) | 1,265 (42.7%) |
| 50+ | 1,143 (7.7%) | 1,836 (4.2%) | 256 (20.7%) | 473 (16.0%) |
| No. of hospitalizations in the past year |  |  |  |  |
| None | 14,200 (96.0%) | 42,070 (97.1%) | 1,008 (81.6%) | 2,698 (91.1%) |
| 1 | 493 (3.3%) | 1,070 (2.5%) | 170 (13.8%) | 185 (6.3%) |
| 2+ | 91 (0.6%) | 190 (0.4%) | 58 (4.7%) | 77 (2.6%) |
| No. of hospitalizations 2-5 years before the diagnosis date |  |  |  |  |
| None | 13,190 (89.2%) | 39,961 (92.2%) | 885 (71.6%) | 2,348 (79.3%) |
| 1 | 1,206 (8.2%) | 2,563 (5.9%) | 187 (15.1%) | 401 (13.5%) |
| 2+ | 388 (2.6%) | 806 (1.9%) | 164 (13.3%) | 211 (7.1%) |
| One or more chronic conditions | 1,886 (12.8%) | 4,035 (9.3%) | 552 (44.7%) | 1,072 (36.2%) |
| Diabetes | 454 (3.1%) | 1,231 (2.8%) | 160 (12.9%) | 458 (15.5%) |
| Chronic cardiovascular disease (excluding hypertension) | 460 (3.1%) | 1,119 (2.6%) | 301 (24.4%) | 584 (19.7%) |
| Peripheral cardiovascular disease | 491 (3.3%) | 717 (1.7%) | 110 (8.9%) | 157 (5.3%) |
| Chronic respiratory disease (excluding asthma) | 750 (5.1%) | 1,480 (3.4%) | 211 (17.1%) | 263 (8.9%) |
| Chronic renal failure | 67 (0.5%) | 111 (0.3%) | 36 (2.9%) | 49 (1.7%) |
| Chronic liver disease | 47 (0.3%) | 119 (0.3%) | 34 (2.8%) | 28 (0.9%) |
| Year of diagnosis |  |  |  |  |
| 1997 | 771 (5.2%) | 2,246 (5.2%) | 48 (3.9%) | 105 (3.5%) |
| 1998 | 649 (4.4%) | 1,903 (4.4%) | 51 (4.1%) | 119 (4.0%) |
| 1999 | 587 (4.0%) | 1,734 (4.0%) | 47 (3.8%) | 113 (3.8%) |
| 2000 | 624 (4.2%) | 1,807 (4.2%) | 56 (4.5%) | 128 (4.3%) |
| 2001 | 598 (4.0%) | 1,764 (4.1%) | 60 (4.9%) | 135 (4.6%) |
| 2002 | 691 (4.7%) | 2,025 (4.7%) | 61 (4.9%) | 127 (4.3%) |
| 2003 | 718 (4.9%) | 2,103 (4.9%) | 52 (4.2%) | 117 (4.0%) |
| 2004 | 757 (5.1%) | 2,193 (5.1%) | 65 (5.3%) | 154 (5.2%) |
| 2005 | 803 (5.4%) | 2,349 (5.4%) | 53 (4.3%) | 119 (4.0%) |
| 2006 | 776 (5.2%) | 2,250 (5.2%) | 58 (4.7%) | 158 (5.3%) |
| 2007 | 731 (4.9%) | 2,153 (5.0%) | 72 (5.8%) | 159 (5.4%) |
| 2008 | 828 (5.6%) | 2,421 (5.6%) | 71 (5.7%) | 171 (5.8%) |
| 2009 | 842 (5.7%) | 2,490 (5.7%) | 69 (5.6%) | 159 (5.4%) |
| 2010 | 917 (6.2%) | 2,701 (6.2%) | 72 (5.8%) | 171 (5.8%) |
| 2011 | 918 (6.2%) | 2,705 (6.2%) | 85 (6.9%) | 210 (7.1%) |
| 2012 | 935 (6.3%) | 2,744 (6.3%) | 64 (5.2%) | 161 (5.4%) |
| 2013 | 828 (5.6%) | 2,419 (5.6%) | 73 (5.9%) | 198 (6.7%) |
| 2014 | 907 (6.1%) | 2,664 (6.1%) | 79 (6.4%) | 205 (6.9%) |
| 2015 | 904 (6.1%) | 2,659 (6.1%) | 100 (8.1%) | 251 (8.5%) |

CIS: Carcinoma *in situ*

Invasive cancer patients have the highest average direct medical cost (about $42,000; Table 3). Among these cancers, anal ($51,000) and oral cavity ($53,000) cancer patients incurred the most costs. The average cost for AGW patients was about $2,100. The median net cost (compared to matched men) of invasive cancer was about $16,000 (Table 4). The highest median net cost was for the cancer of the oral cavity ($27,000). Hospitalization was the biggest contributor to costs for invasive cancer, but outpatient costs were similar for CIS and dominant for AGWs. Overall, 65% of the net cost was due to hospitalization, the cost increased during the study period (more than doubling over 2 decades after inflation-adjustment) and varied by age group and region of residence (Supplementary Tables 34-61).

**Table 3.** Average direct medical cost^1^ of HPV-related disease patients and their matches in 2017 Canadian dollars among Manitoba males (1997-2015)

| HPV-related disease | Patients |  | Matches |  |
| --- | --- | --- | --- | --- |
|  | Number | Average cost | Number | Average cost |
| Anogenital warts | 14,784 | \$2,103 | 43,330 | \$1,048 |
| All HPV-related carcinoma *in situ* | 86 | \$6,144 | 234 | \$4,640 |
| All HPV-related invasive cancers | 1,150 | \$42,484 | 2,726 | \$9,685 |
| Invasive cancer of the anus | 72 | \$51,082 | 187 | \$7,371 |
| Invasive cancer of the oral cavity | 430 | \$52,565 | 1,009 | \$9,533 |
| Invasive cancer of the oropharynx | 510 | \$35,996 | 1,207 | \$9,694 |
| Invasive cancer of the penis | 138 | \$30,564 | 323 | \$11,470 |
| **Total** | **16,020** | **\$5,023** | **46,290** | **\$1,575** |

<sup>1</sup> The periods for which costs were added were 0 to 365 days after diagnosis for AGW and carcinoma *in situ* and 180 days before to 730 days after diagnosis for invasive cancer

**Table 4.** Distribution of net direct medical cost ^1^ of HPV-related diseases (for patients compared to their matches) in 2017 Canadian dollars among Manitoba males (1997-2015)

| HPV-related disease | Average | SD<sup>2</sup> | Median | Q1 - Q3<sup>3</sup> |
| --- | --- | --- | --- | --- |
| **Net total cost** |  |  |  |  |
| Anogenital warts | \$958 | \$10,883 | \$256 | \$-86 - \$941 |
| All HPV-related carcinoma *in situ* | \$1,466 | \$11,483 | \$1,253 | \$-45 - \$4,638 |
| All HPV-related invasive cancers | \$30,382 | \$72,834 | \$16,411 | \$4,527 - \$43,189 |
| Invasive cancer of the anus | \$43,629 | \$56,467 | \$23,712 | \$10,213 - \$72,533 |
| Invasive cancer of the oral cavity | \$40,669 | \$67,962 | \$27,175 | \$7,307 - \$57,202 |
| Invasive cancer of the oropharynx | \$23,530 | \$84,366 | \$12,551 | \$3,683 - \$29,873 |
| Invasive cancer of the penis | \$16,486 | \$33,336 | \$9,417 | \$2,806 - \$21,572 |
| **Net outpatient cost<sup>4</sup>** |  |  |  |  |
| Anogenital warts | \$415 | \$1,115 | \$230 | \$29 - \$575 |
| All HPV-related carcinoma *in situ* | \$987 | \$1,631 | \$632 | \$139 - \$1,467 |
| All HPV-related invasive cancers | \$6,127 | \$6,697 | \$4,672 | \$2,059 - \$8,806 |
| Invasive cancer of the anus | \$6,987 | \$5,871 | \$5,770 | \$3,615 - \$10,543 |
| Invasive cancer of the oral cavity | \$7,952 | \$8,216 | \$5,825 | \$2,203 - \$12,342 |
| Invasive cancer of the oropharynx | \$5,381 | \$5,086 | \$4,693 | \$2,360 - \$7,546 |
| Invasive cancer of the penis | \$2,719 | \$5,183 | \$2,185 | \$689 - \$4,343 |
| **Net hospital cost** |  |  |  |  |
| Anogenital warts | \$352 | \$9,541 | \$0 | \$0 - \$0 |
| All HPV-related carcinoma *in situ* | \$6 | \$10,019 | \$898 | \$-130 - \$2,133 |
| All HPV-related invasive cancers | \$23,245 | \$68,538 | \$10,842 | \$1,553 - \$30,716 |
| Invasive cancer of the anus | \$30,365 | \$49,784 | \$12,780 | \$3,297 - \$46,112 |
| Invasive cancer of the oral cavity | \$31,868 | \$61,562 | \$17,400 | \$4,601 - \$43,137 |
| Invasive cancer of the oropharynx | \$17,805 | \$81,918 | \$6,825 | \$497 - \$23,303 |
| Invasive cancer of the penis | \$12,641 | \$28,358 | \$6,648 | \$742 - \$18,973 |
| **Net drug cost** |  |  |  |  |
| Anogenital warts | \$191 | \$3,613 | \$0 | \$-74 - \$116 |
| All HPV-related carcinoma *in situ* | \$473 | \$1,852 | \$66 | \$-519 - \$835 |
| All HPV-related invasive cancers | \$1,009 | \$6,096 | \$246 | \$-925 - \$1,735 |
| Invasive cancer of the anus | \$6,277 | \$12,320 | \$1,005 | \$-318 - \$6,058 |
| Invasive cancer of the oral cavity | \$849 | \$5,615 | \$246 | \$-1,020 - \$1,721 |
| Invasive cancer of the oropharynx | \$345 | \$4,659 | \$192 | \$-771 - \$1,532 |
| Invasive cancer of the penis | \$1,125 | \$5,915 | \$34 | \$-1,691 - \$1,942 |

<sup>1</sup> The periods for which costs were added were 0 to 365 days after diagnosis for AGW and carcinoma *in situ* and 180 days before to 730 days after diagnosis for invasive cancer; <sup>2</sup> Standard deviation; <sup>3</sup> Q1 and Q3 are the first and third quartile respectively; <sup>4</sup> Outpatient cost includes diagnostic and therapeutic procedures.

Annual healthcare utilization (outpatient visits, hospitalizations, ED visits, and prescriptions) varied by age group, region of residence, and year of diagnosis (Supplementary Tables 62-153). Overall, HPV-RD patients had about 11 outpatient visits and 13 prescriptions per year. These patients had less than 1 annual hospitalization or ED visit. As expected, healthcare utilization was highest for invasive cancer patients.

During 1997–2015, the total net cost of HPV-RDs was about $49 million or about $2.6 million per year. Overall, about 54% (about $26 million) of the total net direct medical costs were attributed to treatment of HPV-caused diseases according to attributable fraction estimates by Volesky *et al*. [27] and about 67% (about $33 million) according to estimates by Saraiya *et al*. [28] (Table 5). The difference was mostly due to different attributable fractions of invasive cancer of the oral cavity. The net attributable cost of AGWs was about $14 million and invasive diseases accounted for $12-$19 million. About half the net attributable cost of HPV-caused cancer was for the treatment of invasive cancer of the oropharynx.

**Table 5.** Total net direct medical cost^1^ of HPV-related diseases (for patients compared to their matches) in 2017 Canadian dollars among Manitoba males according to different estimates of the attributable fraction (1997-2015)

| HPV-related disease | Total cost | Volesky et al. [27] |  | Saraiya et al. [28] |  |
| --- | --- | --- | --- | --- | --- |
|  |  | AF<sup>2</sup> | Attributable cost | AF<sup>2</sup> | Attributable cost |
| Anogenital warts | \$14,164,001 | 100% | \$14,164,001 | 100% | \$14,164,001 |
| All HPV-related carcinoma *in situ* | \$136,963 | <sup>3</sup> | \$25,979 | <sup>4</sup> | \$56,510 |
| All HPV-related invasive cancers | \$34,904,549 | | \$12,250,926 | | \$18,640,337 |
| Invasive cancer of the anus | \$3,141,291 | 88% | \$2,764,336 | 89% | \$2,795,749 |
| Invasive cancer of the oral cavity | \$17,487,701 | 8% | \$1,399,016 | 33% | \$5,770,941 |
| Invasive cancer of the oropharynx | \$12,000,506 | 60% | \$7,200,303 | 72% | \$8,640,364 |
| Invasive cancer of the penis | \$2,275,051 | 39% | \$887,270 | 63% | \$1,433,282 |
| **Total** | **\$49,205,513** | | **\$26,440,906** | | **\$32,860,848** |

<sup>1</sup> The periods for which costs were added were 0 to 365 days after diagnosis for AGW and carcinoma *in situ* and 180 days before to 730 days after diagnosis for invasive cancer; <sup>2</sup> AF = Fraction of disease attributable to HPV; <sup>3</sup> Carcinoma *in situ* AF: 88% for anus, 8% for oral cavity, 60% for oropharynx, and 39% for penis; <sup>4</sup> Carcinoma *in situ* AF: 89% for anus, 33% for oral cavity, 72% for oropharynx, and 63% for penis

The net attributable annual cost was $2.37-$2.95 per male resident and $161-$200 per male newborn over the course of study period (Table 6), although these costs varied by year and were higher at the end of the study period compared to the start.

**Table 6.** Total net direct medical cost^1^ of HPV-related diseases (for patients compared to their matches) in 2017 Canadian dollars among Manitoba males by calendar year according to different estimates of the attributable fraction

| Year | Volesky et al. [27] |  |  | Saraiya et al. [28] |  |  |
| --- | --- | --- | --- | --- | --- | --- |
|  | Total | Per resident<sup>2</sup> | Per newborn<sup>3</sup> | Total | Per resident<sup>2</sup> | Per newborn<sup>3</sup> |
| 1997 | \$850,506 | \$1.73 | \$114 | \$1,098,184 | \$2.24 | \$148 |
| 1998 | \$1,140,336 | \$2.32 | \$152 | \$1,393,839 | \$2.84 | \$186 |
| 1999 | \$990,485 | \$2.01 | \$134 | \$1,244,052 | \$2.52 | \$169 |
| 2000 | \$1,361,579 | \$2.74 | \$189 | \$1,588,603 | \$3.20 | \$221 |
| 2001 | \$1,630,016 | \$3.26 | \$230 | \$2,057,660 | \$4.12 | \$291 |
| 2002 | \$775,782 | \$1.54 | \$110 | \$1,034,121 | \$2.06 | \$146 |
| 2003 | \$1,792,275 | \$3.54 | \$251 | \$1,982,786 | \$3.91 | \$278 |
| 2004 | \$1,289,146 | \$2.52 | \$180 | \$1,683,654 | \$3.30 | \$235 |
| 2005 | \$1,475,677 | \$2.87 | \$200 | \$1,804,581 | \$3.51 | \$245 |
| 2006 | \$581,370 | \$1.13 | \$77 | \$870,593 | \$1.69 | \$116 |
| 2007 | \$1,701,715 | \$3.28 | \$217 | \$2,099,957 | \$4.04 | \$268 |
| 2008 | \$1,839,979 | \$3.50 | \$231 | \$2,342,395 | \$4.46 | \$294 |
| 2009 | \$1,543,021 | \$2.90 | \$190 | \$1,876,320 | \$3.53 | \$232 |
| 2010 | \$1,201,898 | \$2.23 | \$149 | \$1,695,952 | \$3.15 | \$210 |
| 2011 | \$1,903,646 | \$3.48 | \$232 | \$2,335,098 | \$4.27 | \$285 |
| 2012 | \$1,558,377 | \$2.80 | \$184 | \$1,952,947 | \$3.51 | \$230 |
| 2013 | \$1,371,231 | \$2.42 | \$163 | \$1,613,200 | \$2.85 | \$191 |
| 2014 | \$1,617,387 | \$2.81 | \$187 | \$1,981,467 | \$3.45 | \$229 |
| 2015 | \$1,816,481 | \$3.13 | \$212 | \$2,205,438 | \$3.80 | \$258 |
| **Total** | **\$26,440,906** | **\$2.37** | **\$161** | **\$32,860,848** | **\$2.95** | **\$200** |

<sup>1</sup> The periods for which costs were added were 0 to 365 days after diagnosis for AGW and carcinoma *in situ* and 180 days before to 730 days after diagnosis for invasive cancer; <sup>2</sup> Male residents $\geq 9$ years of age; <sup>3</sup> Male newborns

The estimated potential saving from various HPV vaccines was about 30% (about $15 million) for the bivalent vaccine, 56% (about $28 million) for the quadrivalent vaccine, and 60% (about $29 million) for the nonavalent vaccine, according to attributable fractions estimated by Saraiya *et al*. [28] (Table 7). The major difference in these amounts is the exclusion of AGW-causing HPV types in the bivalent vaccine.

**Table 7.** Total net direct medical cost^1^ of HPV-related diseases (for patients compared to their matches) in 2017 Canadian dollars among Manitoba males attributable to the HPV types included in different HPV vaccines (1997-2015)

| HPV-related diseases | Total cost | Saraiya et al. [28] |  |  |  |  |  |
| --- | --- | --- | --- | --- | --- | --- | --- |
|  |  | Bivalent |  | Quadrivalent |  | Nonavalent |  |
|  |  | AF<sup>2</sup> | Attributable cost | AF<sup>2</sup> | Attributable cost | AF<sup>2</sup> | Attributable cost |
| Anogenital warts | \$14,164,001 | 0% | \$0 | 90% | \$12,747,601 | 90% | \$12,747,601 |
| All HPV-related carcinoma *in situ* | \$136,963 | <sup>3</sup> | \$41,480 | <sup>4</sup> | \$41,480 | <sup>5</sup> | \$47,382 |
| All HPV-related invasive cancers | \$34,904,549 | | \$14,981,257 | | \$14,981,257 | | \$16,611,197 |
| Invasive cancer of the anus | \$3,141,291 | 79% | \$2,481,620 | 79% | \$2,481,620 | 83% | \$2,607,272 |
| Invasive cancer of the oral cavity | \$17,487,701 | 22% | \$3,847,294 | 22% | \$3,847,294 | 26% | \$4,546,802 |
| Invasive cancer of the oropharynx | \$12,000,506 | 63% | \$7,560,319 | 63% | \$7,560,319 | 68% | \$8,160,344 |
| Invasive cancer of the penis | \$2,275,051 | 48% | \$1,092,025 | 48% | \$1,092,025 | 57% | \$1,296,779 |
| **Total** | **\$49,205,513** | | **\$15,022,737** | | **\$27,770,338** | | **\$29,406,180** |

<sup>1</sup> The periods for which costs were added were 0 to 365 days after diagnosis for AGW and carcinoma *in situ* and 180 days before to 730 days after diagnosis for invasive cancer; <sup>2</sup> AF = Fraction of disease attributable to HPV; <sup>3</sup> Carcinoma *in situ* AF: 79% for anus, 22% for oral cavity, 63% for oropharynx, and 48% for penis; <sup>4</sup> Carcinoma *in situ* AF: 79% for anus, 22% for oral cavity, 63% for oropharynx, and 48% for penis; <sup>5</sup> Carcinoma *in situ* AF: 83% for anus, 26% for oral cavity, 68% for oropharynx, and 57% for penis

## 4 Discussion

We found that the average cost of treating all conditions attributable to HPV was about $2.60 per male resident per year or about $180 per male newborn (in 2017 Canadian dollars). Invasive cancer accounted for about half of these costs and AGW for roughly the other half. The costs of treating HPV-related cancers was about $16,000; others have estimated the annual costs for common chronic conditions (in 2006 Canadian dollars) at $3,400 for coronary heart disease, $3,800 for diabetes, and $6,000 for stroke [29] (corresponding to $4,100, $4,500, and $7,200 respectively in 2017 Canadian dollars).

The literature on the total economic burden of HPV infections is quite limited, especially when it comes to males. The costs of HPV infections are much higher than any other sexually transmitted infection, except for HIV/AIDS [30]. Differences in HPV-RDs examined, study periods, populations, currencies, methods, and assumptions make it difficult to compare individual studies [10]. Costs are often estimated for men and women combined. For example, the cost per case for oropharyngeal and anal cancer for both genders was estimated as $43,200 and $36,200 (in 2010 US dollars) from a literature review [12]. The same review estimated a cost of $19,800 (in 2010 US dollars) for penile cancer and $810 for AGW (men and women combined) [12]. The costs per anal cancer patient in the US was estimated as $60,913 (in 2014 US dollars) for 66+ Medicare-covered men [31] and $127,531 (2015 US dollars) for <65 commercially insured patients (men and women) [32]. The annual purchasing power parity-adjusted per-capita health expenditure over the last 15 years averaged about $3,800 in Canada and $7,400 in the US in 2010 US dollars [33]. This means the differences between Canada and the US could be explained by the different healthcare systems.

European health systems differ from Canada, so cost estimates from European studies are hard to compare to our results. An Italian study pegged the medical cost attributable to HPV 6, 11, 16, and 18 infections in men at €112.9 million (in 2011 Euros) based on a literature review [34]; they estimated €10,000 in medical cost per penile cancer case and €470 per male AGW case [34]. A French study found that over 95% of costs of HPV-related cancers in men was attributable to head and neck cancer [35]; we found 85% of cancer costs were for cancer of the oral cavity and oropharynx. The French cost for specific HPV-related cancers were estimated between €3,800 and €6,800 (in 2006/07 Euros) for men and women combined [35]. In Spain the male cost of HPV-related cancers ranged between €7,000 and €9,000 (in 2017 Euros), depending on the specific site [36]. These European costs per case all seem lower than the costs we reported from Manitoba, although we used a different methodology.

### 4.1 Strengths and limitations

A major strength of this study is the availability of high quality, comprehensive, population-based healthcare utilization databases and disease registries maintained by the sole health insurer in Manitoba. Since the original purpose of these databases was tracking payment for healthcare providers, our cost estimates were based on direct measurement of actual paid amounts rather than on using average cost estimates derived from micro-costing studies or surveys.

We may not have identified all HPV-RD patients, because of the limitations of retrospective studies and administrative databases. Patients may have been misdiagnosed or the diagnosis date may not have been correct. Disease misclassification could also have occurred due to under-reporting and coding errors (e.g., using the wrong ICD code). Because of the high accuracy of the MCR [21] and the prior validation study of AGW codes [26], we believe disease misclassification was rare in this study.

Our study is limited to the costs that are recorded in the MH datasets, which excludes costs of treating cancer patients in the community, e.g., home intravenous chemotherapy. We do not believe that these excluded costs were significant, because these programs were only recently introduced. We also did not include indirect costs (e.g., loss of employment) of costs incurred by patients (e.g., unpaid time off work), the total costs to all payers is certainly higher than the direct medical costs we report.

The HPV-attributable cost estimates are limited by the accuracy of the estimates of HPV-attributable fractions of disease. We used different estimates of attributable fractions from the literature to account for this uncertainty. Although this leads to roughly similar estimates, analysis based on our results should account for the range of results we report.

## 5 Conclusions

Overall, HPV’s economic burden on males remains significant, the average cost of treating all conditions attributable to HPV was about $2.60 per male resident per year or about $180 per male newborn. Invasive cancer accounted for about half of these costs.

## Data Availability

Data used in this article was derived from administrative health and social data as a secondary use. The data was provided under specific data sharing agreements only for approved use at the Manitoba Centre for Health Policy (MCHP). The original source data is not owned by the researchers or MCHP and as such cannot be provided to a public repository. The original data source and approval for use has been noted in the acknowledgments of the article. Where necessary, source data specific to this article or project may be reviewed at MCHP with the consent of the original data providers, along with the required privacy and ethical review bodies.

## Acknowledgements

The authors acknowledge the Manitoba Centre for Health Policy for use of data contained in the Population Health Research Data Repository under project # 2019-008 (HIPC # 2018/2019-23; REB # HS21956 (H2018:265); RRIC #2018-028). The results and conclusions are those of the authors and no official endorsement by the Manitoba Centre for Health Policy, Manitoba Health, or the Winnipeg Regional Health Authority is intended or should be inferred.

## Conflict of interest statement

CHR has received an unrestricted research grant from Pfizer for an unrelated study. SMM has received unrestricted research grants from Merck, GlaxoSmithKline, Sanofi Pasteur, Pfizer and Roche-Assurex for unrelated studies. SMM has received fees as an advisory board member for Sanofi Pasteur. None of the other authors has any conflicts of interest to disclose.

## Funding sources

This work was supported by the Merck Investigator Studies Program with a grant to the Biomedical Association of Manitoba (BAM). SMM’s work is supported, in part, by funding from the Canada Research Chair Program. The sponsors had no role in the design or conduct of the study, including but not limited to, data identification, collection, management, analysis and interpretation, or preparation, review, or approval of the final results. The opinions presented in the report do not necessarily reflect those of the sponsors.

## Author’s contributions

CHR and SMM designed the study, GP and CHR analyzed the data, CHR, GP and SM interpreted the data, CHR and GP drafted the manuscript. All authors revised the manuscript and approved the final version.

## List of abbreviations

AGW: anogenital wart
CCMB: CancerCare Manitoba
CIS: carcinoma *in situ*
DPIN: Drug Program Information Network
ED: emergency department
EDIS: Emergency Department Information System
HAD: Hospital Abstracts Database
HPV: human papillomavirus
HPV-RD: human papillomavirus-related disease
MCHP: Manitoba Centre for Health Policy
MCR: Manitoba Cancer Registry
MH: Manitoba Health
MHPR: Manitoba Health Population Registry
MSD: Medical Services Database
PHIN: Personal Health Identification Number
WRHA: Winnipeg regional health authority

## Notes

### Author Declarations

This study was approved by the University of Manitoba Research Ethics Board (HS21956 (H2018:265)) and by MH's Health Information Privacy Committee (2018/2019-23).

