## Supplement for "The epidemiology and direct medical costs of diseases associated with human papillomavirus infection among men in Manitoba, Canada"

Supplementary Table 15 Crude incidence rates (95% confidence interval) per 100,000 person-years of all HPV-related invasive cancers among Manitoba males by age group and year of diagnosis. 19

Supplementary Table 22 Crude incidence rates (95% confidence interval) per 100,000 person-years of all HPV-related invasive cancers among Manitoba males by birth year and year of diagnosis. 25

Supplementary Table 119 Distribution of excess number of ER visits[1,2] (per year) due to invasive cancer of the penis (for patients compared to their matches) among Manitoba males by regional health authority and Winnipeg region of residence (1997-2015). 86

Supplementary Table 120 Distribution of excess number of prescriptions[1] (per year) due to invasive cancer of the penis (for patients compared to their matches) among Manitoba males by regional health authority and Winnipeg region of residence (1997-2015). 86

Excess healthcare utilization by year of diagnosis 87

Supplementary Table 121 Distribution of excess number of clinician visits[1] (per year) due to anogenital warts (for patients compared to their matches) among Manitoba males by year of diagnosis (1997-2015). 87

Supplementary Table 122 Distribution of excess number of hospital admissions[1] (per year) due to anogenital warts (for patients compared to their matches) among Manitoba males by year of diagnosis (1997-2015). 88

Supplementary Table 123 Distribution of excess number of ER visits[1,2] (per year) due to anogenital warts (for patients compared to their matches) among Manitoba males by year of diagnosis (1997-2015). 89

Supplementary Table 124 Distribution of excess number of prescriptions[1] (per year) due to anogenital warts (for patients compared to their matches) among Manitoba males by year of diagnosis (1997-2015). 90

Supplementary Table 125 Distribution of excess number of clinician visits[1] (per year) due to HPV-related carcinoma *in situ* (for patients compared to their matches) among Manitoba males by year of diagnosis (1997-2015). 91

Supplementary Table 126 Distribution of excess number of hospital admissions[1] (per year) due to HPV-related carcinoma *in situ* (for patients compared to their matches) among Manitoba males by year of diagnosis (1997-2015). 92

Supplementary Table 127 Distribution of excess number of ER visits[1,2] (per year) due to HPV-related carcinoma *in situ* (for patients compared to their matches) among Manitoba males by year of diagnosis (1997-2015). 93

Supplementary Table 128 Distribution of excess number of prescriptions[1] (per year) due to HPV-related carcinoma *in situ* (for patients compared to their matches) among Manitoba males by year of diagnosis (1997-2015). 94

Supplementary Table 129 Distribution of excess number of clinician visits[1] (per year) due to HPV-related invasive cancer (for patients compared to their matches) among Manitoba males by year of diagnosis (1997-2015). 95

Supplementary Table 130 Distribution of excess number of hospital admissions[1] (per year) due to HPV-related invasive cancer (for patients compared to their matches) among Manitoba males by year of diagnosis (1997-2015). 96

Supplementary Table 131 Distribution of excess number of ER visits[1,2] (per year) due to HPV-related invasive cancer (for patients compared to their matches) among Manitoba males by year of diagnosis (1997-2015). 97

Supplementary Table 132 Distribution of excess number of prescriptions[1] (per year) due to HPV-related invasive cancer (for patients compared to their matches) among Manitoba males by year of diagnosis (1997-2015). 98

### Supplementary tables

Supplementary Table 1: **ICD-O-3 topography and morphology codes for HPV-related cancers of interest**

| Cancer | ICD-O-3 Topography | ICD-O-3 Morphology |
|---|---|---|
| Penile | C60* | Squamous cell carcinoma (8050-8084, 8120-8131) |
| Anal | C21* | Squamous cell carcinoma (8050-8084, 8120-8131; exclude 8077/2) |
| Oropharyngeal | C01*, C02.4, C02.8, C09*, C10.2, C10.8, C10.9, C14* | Squamous cell carcinoma (8050-8084, 8120-8131) |
| Oral cavity | C00.3, C00.4, C00.5, C02.0, C02.1, C02.2, C02.3, C02.9, C03* - C06* | Squamous cell carcinoma (8050-8084, 8120-8131) |

Supplementary Table 2: **Tariff codes used to identify male patients with anogenital warts in the medical claims** [1]

| Code | Description |
|---|---|
| 3372 | Anus, condyloma, single or multiple, internal or external, destruction, in hospital |
| 3433 | Anus, condyloma, external, electrodessication, initial, per sitting |
| 3434 | Anus, condyloma, external, electrodessication, subsequent, per sitting |
| 4120 | Penis, penile skin lesion, including warts, local excision or fulguration, per sitting |

Supplementary Table 3: **Identification of hospitalized patients with anogenital warts** [1]

| Date | ICD version | Criteria | Number of fields[1] |
|---|---|---|---|
| 04/1994-03/2004 | 9 | 078.11 diagnosis OR (078.10 / 078.19 diagnosis AND related procedure) | 16 diagnosis and 12 procedure |
| >03/2004 | 10 | A630 diagnosis OR (B07 diagnosis AND related procedure) | 25 diagnosis and 20 procedure |

[1] All diagnostic and procedure fields were included.

Supplementary Table 4: **ICD-9-CM procedure codes used to assist in the identification of a male with anogenital warts** [1]

| Code | Description |
|---|---|
| 48.82 | Excision of perirectal tissue |
| 49.04 | Other excision of perianal tissue |
| 49.3 | Local excision or destruction of other lesion or tissue of anus |
| 58.3 | Excision or destruction of lesion or tissue of urethra |
| 61.3 | Excision or destruction of lesion or tissue of scrotum |
| 64.2 | Local excision or destruction of lesion of penis |

Supplementary Table 5: **ICD-10 procedure codes used to assist in the identification of a male with anogenital warts** [1]

| Code | Description |
|---|---|
| 1.NT.59.CA.GX | Destruction anus using per orifice approach and device NEC |
| 1.PQ.59.LA.GX | Destruction urethra using open approach and device NEC |
| 1.RY.87.LA | Excision, partial perineum |

Supplementary Table 6: **Tariff codes for treatment of anogenital warts used to assist in the identification of a male with anogenital warts** [1]

| Code | Description |
|---|---|
| 0253 | Excision & simple closure – single lesion, any location |
| 0254 | Excision & simple closure – each additional lesion to a maximum of four |
| 0255 | Excision & closure – multiple lesions, extensive |
| 0397 | Laser vaporization, other than face, one lesion |
| 0398 | Laser vaporization, other than face, two lesions |
| 0399 | Laser vaporization, other than face, three or more lesions |
| 0401 | Cautery (electro, chemo, or simple surgical excision, one lesion) elsewhere |
| 0402 | Warts & fibrocutanous tags - simple |
| 0404 | Cryocautery, etc., of benign lesion of skin, etc., second lesion |
| 0405 | Cryocautery, etc., of benign lesion of skin, etc., subsequent lesions (each) |
| 0406 | Cryocautery, etc., of benign lesion of skin, etc., complicated lesions |
| 3300 | Rectum, villous papilloma of rectum, extensive, local excision |
| 3301 | Rectum, unlisted or unusually complicated |
| 3311 | Rectum, proctosigmoidoscopy |
| 3315 | Rectum, proctosigmoidoscopy with removal of polyp or papilloma, single |
| 3317 | Rectum, proctosigmoidoscopy with removal of polyp or papilloma, multiple |
| 3429 | Anus, unlisted or unusually complicated |
| 3994 | Urethroscopy, therapeutic, polyps, urethral, excision of fulguration with or without urethroscopy |
| 4000 | Urethra, urethroscopy, diagnostic, initial or subsequent |
| 4221 | Scrotum, skin lesion, local excision |
| 4229 | Scrotum, unlisted or unusually complicated |
| 8501 | Office visits, regional, history and examination |
| 8502 | Office visits, complete or extensive re-examination for same illness |
| 8507 | Office visits, subsequent visit |

Supplementary Table 7 **Fraction of HPV-related diseases attributable to the HPV types included in different HPV vaccines** [2]

| **HPV-related disease** | **Bivalent**[1] | **Quadrivalent**[2] | **Nonavalent**[3] |
|---|---|---|---|
| Anogenital warts | 0% | 90% | 90% |
| Carcinoma *in situ* of the anus | 79% | 79% | 83% |
| Carcinoma *in situ* of the oral cavity | 22% | 22% | 26% |
| Carcinoma *in situ* of the oropharynx | 63% | 63% | 68% |
| Carcinoma *in situ* of the penis | 48% | 48% | 57% |
| Invasive cancer of the anus | 79% | 79% | 83% |
| Invasive cancer of the oral cavity | 22% | 22% | 26% |
| Invasive cancer of the oropharynx | 63% | 63% | 68% |
| Invasive cancer of the penis | 48% | 48% | 57% |

[1] Includes HPV type 16 and 18; [2] Includes HPV-type 6, 11, 16, and 18; [3] Includes HPV-type 6, 11, 16, 18, 31, 33, 45, 52, and 58

Supplementary Table 8 **Overall fraction of HPV-related diseases attributable to any HPV type**

| **HPV-related disease** | **Volesky et al.** [3] | **Saraiya et al.** [2] |
|---|---|---|
| Anogenital warts | 100% | 100% |
| Carcinoma *in situ* of the anus | 88% | 89% |
| Carcinoma *in situ* of the oral cavity | 8% | 33% |
| Carcinoma *in situ* of the oropharynx | 60% | 72% |
| Carcinoma *in situ* of the penis | 39% | 63% |
| Invasive cancer of the anus | 88% | 89% |
| Invasive cancer of the oral cavity | 8% | 33% |
| Invasive cancer of the oropharynx | 60% | 72% |
| Invasive cancer of the penis | 39% | 63% |

#### Incidence Rates

Supplementary Table 9 **Age-standardized incidence rates (95% confidence interval) per 100,000 person-years of HPV-related diseases among Manitoba males by regional health authority of residence (1997 - 2016).**

| Regional health authority of residence | Diseases | | | | | |
|---|---|---|---|---|---|---|
| | Anogenital warts | All CIS | CA anus | CA oral cavity | CA oropharynx | CA penis |
| Winnipeg | 174 (170-177) | 1.4 (1.0-1.7) | 1.0 (0.7-1.3) | 5.6 (5.0-6.3) | 6.9 (6.2-7.7) | 1.4 (1.0-1.7) |
| Interlake-Eastern | 105 (98-111) | 1.0 (0.4-1.6) | 0.8 (0.2-1.3) | 4.8 (3.4-6.1) | 6.1 (4.6-7.7) | 1.6 (0.8-2.4) |
| Northern | 59 (53-65) | 2.1 (0.4-3.8) | 1.1 (0.0-2.5) | 3.6 (1.5-5.6) | 6.2 (3.5-9.0) | 2.5 (0.7-4.3) |
| Southern | 91 (86-96) | 0.5 (0.1-0.9) | 0.4 (0.0-0.7) | 4.5 (3.2-5.7) | 3.8 (2.7-4.9) | 1.3 (0.6-1.9) |
| Prairie Mountain | 113 (108-119) | 0.5 (0.1-0.9) | 0.6 (0.2-1.0) | 4.0 (2.9-5.0) | 5.3 (4.1-6.6) | 2.9 (2.0-3.8) |
| Public Trustee/In CFS care | 70 (41-98) | 1.5 (0.0-4.4) | 0.0 (0.0-0.0) | 13.4 (3.8-23.1) | 9.3 (0.0-19.8) | 3.0 (0.0-7.2) |
| **Overall** | **141 (139-143)** | **1.1 (0.9-1.3)** | **0.8 (0.6-1.0)** | **5.1 (4.6-5.6)** | **6.2 (5.7-6.7)** | **1.7 (1.4-2.0)** |

CA: Invasive cancer, CIS: Carcinoma *in situ*

Supplementary Table 10 **Age-standardized incidence rates (95% confidence interval) per 100,000 person-years of HPV-related diseases among Manitoba males by Winnipeg region of residence (1997 - 2016).**

| Winnipeg region of residence | Diseases | | | | | |
|---|---|---|---|---|---|---|
| | Anogenital warts | All CIS | CA anus | CA oral cavity | CA oropharynx | CA penis |
| Northern suburbs | 154 (149-160) | 1.2 (0.7-1.7) | 0.7 (0.3-1.2) | 4.3 (3.3-5.4) | 6.6 (5.3-7.9) | 1.0 (0.5-1.5) |
| Inner city | 174 (166-181) | 1.9 (0.9-3.0) | 1.5 (0.7-2.4) | 7.5 (5.4-9.5) | 7.5 (5.6-9.5) | 2.8 (1.5-4.1) |
| Southern suburbs | 188 (183-192) | 1.3 (0.9-1.8) | 1.0 (0.6-1.4) | 5.9 (5.0-6.9) | 7.0 (6.0-8.0) | 1.2 (0.8-1.6) |
| **Overall** | **174 (171-177)** | **1.4 (1.0-1.7)** | **1.0 (0.7-1.3)** | **5.6 (5.0-6.3)** | **7.0 (6.2-7.7)** | **1.4 (1.0-1.7)** |

CA: Invasive cancer, CIS: Carcinoma *in situ*

Supplementary Table 11 **Age-standardized incidence rates (95% confidence interval) per 100,000 person-years of HPV-related diseases among Manitoba males by year of diagnosis.**

| Year of diagnosis | Diseases | | | | | |
|---|---|---|---|---|---|---|
| | Anogenital warts | All CIS | CA anus | CA oral cavity | CA oropharynx | CA penis |
| 1997 | 144 (134-154) | 0.5 (0.0-1.2) | 0.5 (0.0-1.2) | 6.4 (3.7-9.0) | 4.9 (2.6-7.1) | 0.4 (0.0-1.1) |
| 1998 | 121 (112-131) | 0.7 (0.0-1.6) | 0.8 (0.0-1.7) | 5.8 (3.4-8.3) | 4.6 (2.4-6.8) | 1.4 (0.2-2.7) |
| 1999 | 110 (101-119) | 0.5 (0.0-1.1) | 0.7 (0.0-1.5) | 5.1 (2.8-7.4) | 3.7 (1.8-5.6) | 1.7 (0.4-3.0) |
| 2000 | 117 (108-126) | 1.2 (0.1-2.3) | 0.6 (0.0-1.3) | 5.2 (3.0-7.5) | 5.1 (2.9-7.3) | 1.6 (0.4-2.8) |
| 2001 | 113 (104-122) | 0.8 (0.0-1.7) | 0.3 (0.0-0.8) | 5.5 (3.2-7.8) | 5.7 (3.4-8.1) | 2.2 (0.7-3.6) |
| 2002 | 131 (121-140) | 2.1 (0.6-3.6) | 0.7 (0.0-1.6) | 5.3 (3.0-7.6) | 5.9 (3.5-8.3) | 1.3 (0.2-2.5) |
| 2003 | 135 (125-144) | 0.5 (0.0-1.2) | 0.5 (0.0-1.2) | 4.2 (2.2-6.3) | 6.0 (3.6-8.4) | 1.7 (0.4-3.0) |
| 2004 | 141 (130-151) | 2.3 (0.9-3.8) | 1.0 (0.1-1.9) | 4.5 (2.5-6.6) | 6.1 (3.8-8.5) | 1.2 (0.1-2.2) |
| 2005 | 148 (138-158) | 1.0 (0.0-2.0) | 0.6 (0.0-1.3) | 4.7 (2.6-6.8) | 5.0 (2.9-7.1) | 1.0 (0.0-2.0) |
| 2006 | 143 (133-153) | 0.7 (0.0-1.5) | 0.7 (0.0-1.5) | 5.3 (3.1-7.5) | 6.0 (3.7-8.3) | 0.8 (0.0-1.6) |
| 2007 | 135 (125-145) | 1.3 (0.3-2.4) | 1.4 (0.3-2.5) | 4.7 (2.7-6.8) | 7.2 (4.7-9.8) | 2.2 (0.8-3.7) |
| 2008 | 152 (142-163) | 0.5 (0.0-1.1) | 1.2 (0.1-2.2) | 5.2 (3.1-7.4) | 6.4 (4.1-8.8) | 2.7 (1.2-4.3) |
| 2009 | 153 (143-163) | 1.5 (0.4-2.6) | 0.8 (0.0-1.6) | 5.4 (3.3-7.6) | 6.2 (3.9-8.5) | 1.4 (0.3-2.5) |
| 2010 | 163 (152-173) | 1.1 (0.1-2.0) | 0.9 (0.0-1.7) | 6.5 (4.2-8.9) | 6.7 (4.3-9.1) | 0.5 (0.0-1.1) |
| 2011 | 160 (150-171) | 1.0 (0.1-1.9) | 1.1 (0.1-2.0) | 5.7 (3.5-7.8) | 8.6 (6.0-11.2) | 1.1 (0.1-2.1) |
| 2012 | 158 (148-168) | 0.6 (0.0-1.3) | 0.4 (0.0-0.9) | 5.5 (3.4-7.6) | 5.5 (3.4-7.6) | 1.1 (0.1-2.0) |
| 2013 | 139 (129-148) | 1.1 (0.2-2.0) | 1.4 (0.4-2.4) | 3.4 (1.8-5.1) | 6.4 (4.2-8.6) | 2.0 (0.8-3.2) |
| 2014 | 150 (140-160) | 1.3 (0.3-2.2) | 1.2 (0.2-2.1) | 3.4 (1.8-5.0) | 6.4 (4.2-8.6) | 3.0 (1.5-4.5) |
| 2015 | 149 (140-159) | 1.1 (0.2-2.0) | 1.1 (0.2-2.0) | 6.3 (4.1-8.4) | 6.8 (4.6-9.0) | 3.9 (2.2-5.6) |
| 2016 | 144 (135-154) | 2.0 (0.8-3.2) | 0.4 (0.0-0.9) | 4.1 (2.4-5.8) | 7.8 (5.4-10.1) | 1.7 (0.6-2.8) |
| **Overall** | **141 (139-143)** | **1.1 (0.9-1.3)** | **0.8 (0.6-1.0)** | **5.1 (4.6-5.6)** | **6.2 (5.7-6.7)** | **1.7 (1.4-2.0)** |

CA: Invasive cancer, CIS: Carcinoma *in situ*

Supplementary Table 12 **Crude incidence rates (95% confidence interval) per 100,000 person-years of anogenital warts among Manitoba males by age group and regional health authority and Winnipeg region of residence (1997 - 2016).**

| **Region** | **Age group** | | | | | |
|---|---|---|---|---|---|---|
| | **9-17** | **18-30** | **31-50** | **51-64** | **≥65** | **Overall** |
| Winnipeg | 24 (21-28) | 439 (427-451) | 212 (205-218) | 82 (76-87) | 38 (33-42) | 188 (185-192) |
| Northern suburbs | 21 (16-28) | 396 (377-416) | 187 (177-198) | 69 (60-78) | 34 (27-42) | 166 (160-172) |
| Inner city | 27 (19-38) | 397 (372-423) | 220 (205-235) | 101 (86-117) | 43 (32-58) | 199 (190-208) |
| Southern suburbs | 25 (21-31) | 486 (468-504) | 226 (216-235) | 84 (77-92) | 38 (33-44) | 200 (195-205) |
| Interlake-Eastern | 15 (10-23) | 277 (254-302) | 122 (110-134) | 44 (36-54) | 25 (18-33) | 102 (96-109) |
| Northern | 15 (9-24) | 140 (122-161) | 64 (54-77) | 33 (23-48) | 23 (11-44) | 65 (59-72) |
| Southern | 14 (10-19) | 249 (232-268) | 103 (94-113) | 33 (26-41) | 23 (16-30) | 96 (91-101) |
| Prairie Mountain | 23 (17-30) | 357 (335-380) | 102 (92-112) | 41 (34-49) | 21 (15-27) | 114 (108-119) |
| Public Trustee/In CFS care | 64 (32-114) | 226 (117-394) | 35 (7-102) | 13 (0-73) | 34 (9-88) | 61 (42-87) |
| Overall | 21 (19-24) | 367 (359-375) | 167 (162-171) | 63 (59-66) | 31 (28-34) | 149 (147-152) |

#### Incidence Rates by age and year of diagnosis

Supplementary Table 13 **Crude incidence rates (95% confidence interval) per 100,000 person-years of anogenital warts among Manitoba males by age group and year of diagnosis.**

| Age group | Year of diagnosis | | | | |
|---|---|---|---|---|---|
| | 1997-2001 | 2002-2006 | 2007-2011 | 2012-2016 | Overall |
| 9-17 | 19 (15-24) | 22 (17-27) | 20 (16-25) | 23 (19-29) | 21 (19-24) |
| 18-30 | 314 (299-330) | 370 (354-387) | 411 (394-428) | 370 (355-386) | 367 (359-375) |
| 31-50 | 150 (142-158) | 166 (157-174) | 172 (163-181) | 180 (171-189) | 167 (163-171) |
| 51-64 | 49 (42-57) | 55 (48-63) | 70 (63-78) | 71 (64-78) | 63 (59-67) |
| ≥65 | 24 (19-30) | 30 (25-37) | 32 (27-39) | 35 (30-41) | 31 (28-34) |
| **Overall** | **131 (127-136)** | **148 (143-152)** | **160 (155-165)** | **156 (152-161)** | **149 (147-152)** |

Supplementary Table 14 **Crude incidence rates (95% confidence interval) per 100,000 person-years of all HPV-related carcinoma *in situ* among Manitoba males by age group and year of diagnosis.**

| Age group | Year of diagnosis | | | | |
|---|---|---|---|---|---|
| | 1997-2001 | 2002-2006 | 2007-2011 | 2012-2016 | Overall |
| 9-17 | 0.0 (0.0-1.0) | 0.0 (0.0-0.9) | 0.0 (0.0-1.0) | 0.0 (0.0-1.0) | 0.0 (0.0-0.2) |
| 18-30 | 0.4 (0.0-1.4) | 0.2 (0.0-1.1) | 0.4 (0.0-1.3) | 0.5 (0.1-1.4) | 0.4 (0.2-0.7) |
| 31-50 | 0.3 (0.1-1.0) | 0.2 (0.0-0.8) | 0.6 (0.2-1.4) | 1.0 (0.5-2.0) | 0.6 (0.3-0.9) |
| 51-64 | 1.6 (0.6-3.5) | 1.8 (0.8-3.5) | 1.1 (0.4-2.5) | 1.9 (0.9-3.3) | 1.6 (1.1-2.3) |
| ≥65 | 1.2 (0.3-3.1) | 4.7 (2.7-7.7) | 3.3 (1.7-5.8) | 2.4 (1.1-4.3) | 2.9 (2.1-3.9) |
| **Overall** | **0.6 (0.3-1.0)** | **1.1 (0.7-1.5)** | **0.9 (0.6-1.4)** | **1.2 (0.8-1.6)** | **0.9 (0.8-1.2)** |

Supplementary Table 15 **Crude incidence rates (95% confidence interval) per 100,000 person-years of all HPV-related invasive cancers among Manitoba males by age group and year of diagnosis.**

| Age group | Year of diagnosis | | | | |
|---|---|---|---|---|---|
| | 1997-2001 | 2002-2006 | 2007-2011 | 2012-2016 | Overall |
| 9-17 | 0.0 (0.0-1.0) | 0.0 (0.0-0.9) | 0.0 (0.0-1.0) | 0.0 (0.0-1.0) | 0.0 (0.0-0.2) |

| | | | | | |
|---|---|---|---|---|---|
| 18-30 | 0.4 (0.0-1.4) | 0.4 (0.0-1.4) | 0.2 (0.0-1.0) | 0.3 (0.0-1.2) | 0.3 (0.1-0.7) |
| 31-50 | 7.0 (5.4-9.0) | 5.5 (4.0-7.3) | 4.8 (3.5-6.6) | 5.7 (4.2-7.5) | 5.8 (5.0-6.6) |
| 51-64 | 22.5 (18.0-27.8) | 21.5 (17.4-26.3) | 31.1 (26.5-36.3) | 26.8 (22.8-31.3) | 25.9 (23.7-28.3) |
| ≥65 | 30.1 (24.4-36.6) | 34.6 (28.6-41.5) | 38.7 (32.6-45.7) | 38.0 (32.4-44.4) | 35.6 (32.6-38.8) |
| **Overall** | **10.0 (8.8-11.3)** | **10.3 (9.1-11.6)** | **13.0 (11.7-14.4)** | **12.9 (11.6-14.3)** | **11.6 (11.0-12.3)** |

Supplementary Table 16 **Crude incidence rates (95% confidence interval) per 100,000 person-years of invasive cancer of the anus among Manitoba males by age group and year of diagnosis.**

| Age group | Year of diagnosis | | | | |
|---|---|---|---|---|---|
| | 1997-2001 | 2002-2006 | 2007-2011 | 2012-2016 | Overall |
| 9-17 | 0.0 (0.0-1.0) | 0.0 (0.0-0.9) | 0.0 (0.0-1.0) | 0.0 (0.0-1.0) | 0.0 (0.0-0.2) |
| 18-30 | 0.0 (0.0-0.7) | 0.2 (0.0-1.1) | 0.0 (0.0-0.7) | 0.0 (0.0-0.6) | 0.0 (0.0-0.3) |
| 31-50 | 0.3 (0.1-1.0) | 0.7 (0.3-1.5) | 0.2 (0.0-0.9) | 0.2 (0.0-0.8) | 0.4 (0.2-0.6) |
| 51-64 | 1.3 (0.4-3.1) | 0.9 (0.2-2.3) | 2.5 (1.3-4.2) | 2.4 (1.3-4.0) | 1.9 (1.3-2.6) |
| ≥65 | 0.9 (0.2-2.7) | 1.5 (0.5-3.5) | 2.5 (1.1-4.7) | 1.7 (0.7-3.4) | 1.6 (1.1-2.5) |
| **Overall** | **0.4 (0.2-0.8)** | **0.6 (0.4-1.0)** | **0.9 (0.6-1.3)** | **0.8 (0.5-1.2)** | **0.7 (0.6-0.9)** |

Supplementary Table 17 **Crude incidence rates (95% confidence interval) per 100,000 person-years of invasive cancer of the oral cavity among Manitoba males by age group and year of diagnosis.**

| Age group | Year of diagnosis | | | | |
|---|---|---|---|---|---|
| | 1997-2001 | 2002-2006 | 2007-2011 | 2012-2016 | Overall |
| 9-17 | 0.0 (0.0-1.0) | 0.0 (0.0-0.9) | 0.0 (0.0-1.0) | 0.0 (0.0-1.0) | 0.0 (0.0-0.2) |
| 18-30 | 0.0 (0.0-0.7) | 0.2 (0.0-1.1) | 0.2 (0.0-1.0) | 0.2 (0.0-0.9) | 0.1 (0.0-0.4) |
| 31-50 | 2.9 (1.9-4.2) | 1.6 (0.9-2.7) | 1.3 (0.6-2.3) | 2.3 (1.4-3.6) | 2.0 (1.6-2.6) |
| 51-64 | 9.8 (6.9-13.5) | 9.2 (6.6-12.5) | 11.1 (8.4-14.3) | 6.6 (4.7-9.0) | 9.0 (7.7-10.5) |
| ≥65 | 14.6 (10.7-19.3) | 13.0 (9.5-17.5) | 15.1 (11.4-19.7) | 13.5 (10.2-17.4) | 14.0 (12.2-16.1) |
| **Overall** | **4.5 (3.7-5.4)** | **3.9 (3.2-4.8)** | **4.7 (3.9-5.6)** | **4.1 (3.4-4.9)** | **4.3 (3.9-4.7)** |

Supplementary Table 18 **Crude incidence rates (95% confidence interval) per 100,000 person-years of invasive cancer of the oropharynx among Manitoba males by age group and year of diagnosis.**

| Age group | Year of diagnosis | | | | |
|---|---|---|---|---|---|
| | 1997-2001 | 2002-2006 | 2007-2011 | 2012-2016 | Overall |
| 9-17 | 0.0 (0.0-1.0) | 0.0 (0.0-0.9) | 0.0 (0.0-1.0) | 0.0 (0.0-1.0) | 0.0 (0.0-0.2) |

| | | | | | |
|---|---|---|---|---|---|
| 18-30 | 0.0 (0.0-0.7) | 0.0 (0.0-0.7) | 0.0 (0.0-0.7) | 0.0 (0.0-0.6) | 0.0 (0.0-0.2) |
| 31-50 | 3.0 (1.9-4.4) | 2.9 (1.9-4.3) | 3.1 (2.0-4.5) | 2.7 (1.7-4.0) | 2.9 (2.4-3.5) |
| 51-64 | 9.3 (6.5-12.9) | 10.8 (7.9-14.3) | 16.4 (13.1-20.3) | 15.4 (12.4-18.9) | 13.4 (11.8-15.2) |
| ≥65 | 10.6 (7.4-14.8) | 14.5 (10.7-19.2) | 14.0 (10.4-18.4) | 13.5 (10.2-17.4) | 13.2 (11.4-15.2) |
| **Overall** | **3.9 (3.1-4.7)** | **4.8 (4.0-5.7)** | **6.1 (5.2-7.1)** | **6.0 (5.1-6.9)** | **5.2 (4.8-5.7)** |

Supplementary Table 19 **Crude incidence rates (95% confidence interval) per 100,000 person-years of invasive cancer of the penis among Manitoba males by age group and year of diagnosis.**

| Age group | Year of diagnosis | | | | |
|---|---|---|---|---|---|
| | 1997-2001 | 2002-2006 | 2007-2011 | 2012-2016 | Overall |
| 9-17 | 0.0 (0.0-1.0) | 0.0 (0.0-0.9) | 0.0 (0.0-1.0) | 0.0 (0.0-1.0) | 0.0 (0.0-0.2) |
| 18-30 | 0.4 (0.0-1.4) | 0.0 (0.0-0.7) | 0.0 (0.0-0.7) | 0.2 (0.0-0.9) | 0.1 (0.0-0.4) |
| 31-50 | 0.8 (0.3-1.7) | 0.2 (0.0-0.8) | 0.2 (0.0-0.9) | 0.5 (0.1-1.2) | 0.4 (0.2-0.7) |
| 51-64 | 2.1 (0.9-4.2) | 0.7 (0.1-2.0) | 1.1 (0.4-2.5) | 2.4 (1.3-4.0) | 1.6 (1.1-2.3) |
| ≥65 | 3.9 (2.1-6.7) | 5.6 (3.4-8.8) | 7.1 (4.7-10.5) | 9.4 (6.7-12.9) | 6.7 (5.5-8.2) |
| **Overall** | **1.2 (0.8-1.7)** | **0.9 (0.6-1.4)** | **1.3 (0.9-1.8)** | **2.1 (1.6-2.7)** | **1.4 (1.2-1.6)** |

#### Incidence Rates by birth year cohort and year of diagnosis

Supplementary Table 20 **Crude incidence rates (95% confidence interval) per 100,000 person-years of anogenital warts among Manitoba males by birth year and year of diagnosis.**

| Birth year | Year of diagnosis | | | | |
|---|---|---|---|---|---|
| | 1997-2001 | 2002-2006 | 2007-2011 | 2012-2016 | Overall |
| 1920-1929 | 25 (18-34) | 25 (17-35) | 20 (12-33) | 12 (5-27) | 22 (18-28) |
| 1930-1939 | 38 (30-47) | 39 (31-50) | 37 (29-48) | 22 (15-32) | 35 (31-40) |
| 1940-1949 | 59 (51-68) | 55 (47-64) | 48 (41-57) | 45 (37-54) | 52 (48-57) |
| 1950-1959 | 106 (97-117) | 84 (76-93) | 80 (72-89) | 65 (57-73) | 84 (80-89) |
| 1960-1969 | 226 (213-241) | 174 (162-186) | 123 (112-133) | 110 (101-120) | 158 (152-164) |
| 1970-1979 | 332 (314-351) | 342 (324-361) | 274 (258-291) | 180 (168-193) | 280 (272-288) |
| 1980-1989 | 40 (34-47) | 253 (238-269) | 439 (419-459) | 357 (340-375) | 276 (268-284) |
| 1990-1999 | 15 (6-33) | 5 (2-8) | 61 (54-69) | 248 (234-262) | 123 (117-130) |
| **Overall** | **131 (127-136)** | **148 (143-152)** | **160 (155-165)** | **156 (152-161)** | **149 (147-152)** |

Supplementary Table 21 **Crude incidence rates (95% confidence interval) per 100,000 person-years of all HPV-related carcinoma *in situ* among Manitoba males by birth year and year of diagnosis.**

| Birth year | Year of diagnosis | | | | |
|---|---|---|---|---|---|
| | 1997-2001 | 2002-2006 | 2007-2011 | 2012-2016 | Overall |
| 1920-1929 | 1.9 (0.4-5.5) | 4.9 (1.8-10.7) | 2.4 (0.3-8.6) | 2.1 (0.1-11.5) | 2.9 (1.5-5.1) |
| 1930-1939 | 1.4 (0.3-4.2) | 4.8 (2.2-9.1) | 4.4 (1.8-9.0) | 3.1 (0.8-7.9) | 3.3 (2.1-5.0) |
| 1940-1949 | 1.0 (0.2-2.8) | 1.3 (0.4-3.4) | 1.4 (0.4-3.7) | 1.9 (0.6-4.5) | 1.4 (0.8-2.3) |
| 1950-1959 | 0.5 (0.1-1.7) | 0.7 (0.1-2.1) | 1.4 (0.5-3.1) | 2.2 (1.0-4.2) | 1.2 (0.7-1.8) |
| 1960-1969 | 0.2 (0.0-1.3) | 0.2 (0.0-1.3) | 0.5 (0.1-1.6) | 0.9 (0.2-2.3) | 0.5 (0.2-0.9) |
| 1970-1979 | 0.5 (0.1-1.8) | 0.3 (0.0-1.5) | 0.5 (0.1-1.8) | 0.5 (0.1-1.7) | 0.4 (0.2-0.9) |
| 1980-1989 | 0.0 (0.0-0.9) | 0.2 (0.0-1.3) | 0.5 (0.1-1.7) | 1.5 (0.6-3.2) | 0.6 (0.3-1.1) |
| 1990-1999 | 0.0 (0.0-9.3) | 0.0 (0.0-1.5) | 0.0 (0.0-0.9) | 0.2 (0.0-1.2) | 0.1 (0.0-0.5) |
| **Overall** | **0.6 (0.3-1.0)** | **1.1 (0.7-1.5)** | **0.9 (0.6-1.4)** | **1.2 (0.8-1.6)** | **0.9 (0.8-1.2)** |

Supplementary Table 22 **Crude incidence rates (95% confidence interval) per 100,000 person-years of all HPV-related invasive cancers among Manitoba males by birth year and year of diagnosis.**

| Birth year | Year of diagnosis | | | | |
|---|---|---|---|---|---|
| | **1997-2001** | **2002-2006** | **2007-2011** | **2012-2016** | **Overall** |
| 1920-1929 | 27.6 (20.0-37.0) | 37.6 (27.6-50.2) | 31.1 (20.3-45.5) | 24.8 (12.8-43.3) | 30.9 (25.8-36.8) |
| 1930-1939 | 25.2 (18.9-32.9) | 32.5 (24.9-41.8) | 38.6 (29.6-49.5) | 32.4 (23.3-43.7) | 31.7 (27.6-36.2) |
| 1940-1949 | 20.1 (15.4-25.7) | 23.8 (18.6-30.0) | 43.9 (36.5-52.3) | 41.6 (34.1-50.2) | 31.7 (28.5-35.1) |
| 1950-1959 | 12.0 (9.0-15.7) | 13.1 (9.9-17.1) | 22.1 (17.8-27.1) | 32.3 (27.0-38.4) | 19.7 (17.6-21.9) |
| 1960-1969 | 0.7 (0.1-2.0) | 3.2 (1.8-5.4) | 6.3 (4.2-9.1) | 12.0 (9.0-15.6) | 5.6 (4.5-6.8) |
| 1970-1979 | 0.5 (0.1-1.8) | 1.3 (0.4-3.0) | 2.0 (0.9-4.0) | 3.6 (2.0-5.9) | 1.9 (1.3-2.7) |
| 1980-1989 | 0.0 (0.0-0.9) | 0.0 (0.0-0.9) | 0.2 (0.0-1.3) | 1.8 (0.8-3.5) | 0.5 (0.2-1.0) |
| 1990-1999 | 0.0 (0.0-9.3) | 0.0 (0.0-1.5) | 0.0 (0.0-0.9) | 0.2 (0.0-1.2) | 0.1 (0.0-0.5) |
| **Overall** | **10.0 (8.8-11.3)** | **10.3 (9.1-11.6)** | **13.0 (11.7-14.4)** | **12.9 (11.6-14.3)** | **11.6 (11.0-12.3)** |

Supplementary Table 23 **Crude incidence rates (95% confidence interval) per 100,000 person-years of invasive cancer of the anus among Manitoba males by birth year and year of diagnosis.**

| Birth year | Year of diagnosis | | | | |
|---|---|---|---|---|---|
| | **1997-2001** | **2002-2006** | **2007-2011** | **2012-2016** | **Overall** |
| 1920-1929 | 0.6 (0.0-3.5) | 3.3 (0.9-8.4) | 1.2 (0.0-6.7) | 0.0 (0.0-7.6) | 1.4 (0.5-3.2) |
| 1930-1939 | 1.9 (0.5-4.9) | 0.5 (0.0-3.0) | 3.7 (1.4-8.1) | 2.3 (0.5-6.8) | 2.0 (1.1-3.4) |
| 1940-1949 | 0.6 (0.1-2.3) | 1.0 (0.2-2.9) | 2.5 (1.0-5.1) | 1.6 (0.4-4.0) | 1.4 (0.8-2.3) |
| 1950-1959 | 0.7 (0.1-2.0) | 0.5 (0.1-1.7) | 1.9 (0.8-3.8) | 3.0 (1.5-5.2) | 1.5 (1.0-2.2) |
| 1960-1969 | 0.0 (0.0-0.8) | 0.9 (0.2-2.3) | 0.2 (0.0-1.3) | 0.7 (0.1-1.9) | 0.5 (0.2-0.9) |
| 1970-1979 | 0.0 (0.0-0.9) | 0.5 (0.1-1.9) | 0.3 (0.0-1.4) | 0.0 (0.0-0.9) | 0.2 (0.0-0.6) |
| 1980-1989 | 0.0 (0.0-0.9) | 0.0 (0.0-0.9) | 0.0 (0.0-0.9) | 0.2 (0.0-1.2) | 0.1 (0.0-0.3) |
| 1990-1999 | 0.0 (0.0-9.3) | 0.0 (0.0-1.5) | 0.0 (0.0-0.9) | 0.0 (0.0-0.8) | 0.0 (0.0-0.3) |
| **Overall** | **0.4 (0.2-0.8)** | **0.6 (0.4-1.0)** | **0.9 (0.6-1.3)** | **0.8 (0.5-1.2)** | **0.7 (0.6-0.9)** |

Supplementary Table 24 **Crude incidence rates (95% confidence interval) per 100,000 person-years of invasive cancer of the oral cavity among Manitoba males by birth year and year of diagnosis.**

| Birth year | Year of diagnosis | | | | |
|---|---|---|---|---|---|
| | 1997-2001 | 2002-2006 | 2007-2011 | 2012-2016 | Overall |
| 1920-1929 | 12.5 (7.7-19.3) | 14.7 (8.7-23.3) | 13.1 (6.6-23.5) | 12.4 (4.6-27.0) | 13.3 (10.0-17.3) |
| 1930-1939 | 12.8 (8.4-18.7) | 11.2 (6.9-17.1) | 13.7 (8.6-20.8) | 11.6 (6.5-19.1) | 12.3 (9.9-15.3) |
| 1940-1949 | 9.2 (6.2-13.3) | 9.4 (6.2-13.5) | 15.7 (11.4-21.1) | 14.8 (10.4-20.3) | 12.1 (10.2-14.3) |
| 1950-1959 | 4.4 (2.6-6.8) | 5.6 (3.6-8.4) | 8.2 (5.7-11.4) | 7.9 (5.4-11.2) | 6.5 (5.3-7.8) |
| 1960-1969 | 0.2 (0.0-1.3) | 0.7 (0.1-2.0) | 1.8 (0.8-3.6) | 2.9 (1.5-4.9) | 1.4 (0.9-2.1) |
| 1970-1979 | 0.0 (0.0-0.9) | 0.5 (0.1-1.9) | 0.5 (0.1-1.8) | 2.1 (1.0-4.1) | 0.8 (0.4-1.4) |
| 1980-1989 | 0.0 (0.0-0.9) | 0.0 (0.0-0.9) | 0.2 (0.0-1.3) | 0.9 (0.2-2.2) | 0.3 (0.1-0.7) |
| 1990-1999 | 0.0 (0.0-9.3) | 0.0 (0.0-1.5) | 0.0 (0.0-0.9) | 0.0 (0.0-0.8) | 0.0 (0.0-0.3) |
| **Overall** | **4.5 (3.7-5.4)** | **3.9 (3.2-4.8)** | **4.7 (3.9-5.6)** | **4.1 (3.4-4.9)** | **4.3 (3.9-4.7)** |

Supplementary Table 25 **Crude incidence rates (95% confidence interval) per 100,000 person-years of invasive cancer of the oropharynx among Manitoba males by birth year and year of diagnosis.**

| Birth year | Year of diagnosis | | | | |
|---|---|---|---|---|---|
| | 1997-2001 | 2002-2006 | 2007-2011 | 2012-2016 | Overall |
| 1920-1929 | 13.2 (8.1-20.1) | 11.5 (6.3-19.2) | 9.6 (4.1-18.8) | 4.1 (0.5-14.9) | 10.9 (7.9-14.5) |
| 1930-1939 | 7.6 (4.3-12.3) | 17.6 (12.1-24.7) | 13.1 (8.1-20.0) | 6.2 (2.7-12.1) | 11.3 (9.0-14.1) |
| 1940-1949 | 8.6 (5.7-12.5) | 12.4 (8.7-17.1) | 22.5 (17.3-28.7) | 17.1 (12.4-22.9) | 14.9 (12.7-17.3) |
| 1950-1959 | 5.3 (3.4-8.0) | 6.6 (4.4-9.5) | 11.3 (8.3-15.0) | 18.8 (14.8-23.5) | 10.3 (8.9-12.0) |
| 1960-1969 | 0.5 (0.1-1.6) | 1.4 (0.5-3.0) | 4.1 (2.4-6.4) | 7.5 (5.2-10.5) | 3.4 (2.6-4.4) |
| 1970-1979 | 0.0 (0.0-0.9) | 0.3 (0.0-1.5) | 1.3 (0.4-3.0) | 1.2 (0.4-2.8) | 0.7 (0.3-1.2) |
| 1980-1989 | 0.0 (0.0-0.9) | 0.0 (0.0-0.9) | 0.0 (0.0-0.9) | 0.4 (0.1-1.6) | 0.1 (0.0-0.4) |
| 1990-1999 | 0.0 (0.0-9.3) | 0.0 (0.0-1.5) | 0.0 (0.0-0.9) | 0.0 (0.0-0.8) | 0.0 (0.0-0.3) |
| **Overall** | **3.9 (3.1-4.7)** | **4.8 (4.0-5.7)** | **6.1 (5.2-7.1)** | **6.0 (5.1-6.9)** | **5.2 (4.8-5.7)** |

Supplementary Table 26 **Crude incidence rates (95% confidence interval) per 100,000 person-years of invasive cancer of the penis among Manitoba males by birth year and year of diagnosis.**

| Birth year | Year of diagnosis | | | | |
|---|---|---|---|---|---|
| | **1997-2001** | **2002-2006** | **2007-2011** | **2012-2016** | **Overall** |
| 1920-1929 | 1.3 (0.2-4.5) | 8.2 (3.9-15.0) | 7.2 (2.6-15.6) | 8.3 (2.3-21.2) | 5.3 (3.3-8.0) |
| 1930-1939 | 2.8 (1.0-6.2) | 3.2 (1.2-7.0) | 8.1 (4.3-13.9) | 12.3 (7.0-20.0) | 6.0 (4.3-8.1) |
| 1940-1949 | 1.6 (0.5-3.7) | 1.0 (0.2-2.9) | 3.2 (1.5-6.1) | 8.2 (5.0-12.5) | 3.3 (2.3-4.5) |
| 1950-1959 | 1.6 (0.6-3.3) | 0.5 (0.1-1.7) | 0.7 (0.1-2.1) | 2.7 (1.4-4.9) | 1.4 (0.9-2.1) |
| 1960-1969 | 0.0 (0.0-0.8) | 0.2 (0.0-1.3) | 0.2 (0.0-1.3) | 0.9 (0.2-2.3) | 0.3 (0.1-0.7) |
| 1970-1979 | 0.5 (0.1-1.8) | 0.0 (0.0-1.0) | 0.0 (0.0-0.9) | 0.2 (0.0-1.3) | 0.2 (0.0-0.6) |
| 1980-1989 | 0.0 (0.0-0.9) | 0.0 (0.0-0.9) | 0.0 (0.0-0.9) | 0.2 (0.0-1.2) | 0.1 (0.0-0.3) |
| 1990-1999 | 0.0 (0.0-9.3) | 0.0 (0.0-1.5) | 0.0 (0.0-0.9) | 0.2 (0.0-1.2) | 0.1 (0.0-0.5) |
| **Overall** | **1.2 (0.8-1.7)** | **0.9 (0.6-1.4)** | **1.3 (0.9-1.8)** | **2.1 (1.6-2.7)** | **1.4 (1.2-1.6)** |

#### Incidence Rates by birth year cohort and age

Supplementary Table 27 **Crude incidence rates (95% confidence interval) per 100,000 person-years of anogenital warts among Manitoba males by birth year and age group.**

| Birth year | Age group | | | | | |
|---|---|---|---|---|---|---|
| | 9-17 | 18-30 | 31-50 | 51-64 | ≥65 | Overall |
| 1920-1929 | n/a | n/a | n/a | n/a | 22 (18-28) | 22 (18-28) |
| 1930-1939 | n/a | n/a | n/a | 44 (34-57) | 33 (28-38) | 35 (31-40) |
| 1940-1949 | n/a | n/a | 91 (69-119) | 53 (48-59) | 43 (37-51) | 52 (48-57) |
| 1950-1959 | n/a | n/a | 102 (95-110) | 68 (63-74) | 52 (21-107) | 84 (80-89) |
| 1960-1969 | n/a | 349 (304-399) | 157 (151-164) | 92 (79-108) | n/a | 158 (152-164) |
| 1970-1979 | 172 (69-355) | 354 (340-368) | 223 (214-233) | n/a | n/a | 280 (272-288) |
| 1980-1989 | 28 (23-33) | 392 (380-404) | 289 (263-316) | n/a | n/a | 276 (268-284) |
| 1990-1999 | 20 (17-23) | 328 (310-347) | n/a | n/a | n/a | 123 (117-130) |
| **Overall** | **21 (19-24)** | **367 (359-375)** | **167 (163-171)** | **63 (59-67)** | **31 (28-34)** | **149 (147-152)** |

Supplementary Table 28 **Crude incidence rates (95% confidence interval) per 100,000 person-years of all HPV-related carcinoma *in situ* among Manitoba males by birth year and age group.**

| Birth year | Age group | | | | | |
|---|---|---|---|---|---|---|
| | 9-17 | 18-30 | 31-50 | 51-64 | ≥65 | Overall |
| 1920-1929 | n/a | n/a | n/a | n/a | 2.9 (1.5-5.1) | 2.9 (1.5-5.1) |
| 1930-1939 | n/a | n/a | n/a | 2.9 (0.8-7.3) | 3.5 (2.1-5.4) | 3.3 (2.1-5.0) |
| 1940-1949 | n/a | n/a | 0.0 (0.0-6.1) | 1.1 (0.5-2.1) | 2.3 (1.0-4.6) | 1.4 (0.8-2.3) |
| 1950-1959 | n/a | n/a | 0.4 (0.1-1.1) | 1.9 (1.1-3.1) | 0.0 (0.0-27.4) | 1.2 (0.7-1.8) |
| 1960-1969 | n/a | 0.0 (0.0-5.9) | 0.4 (0.1-0.9) | 1.2 (0.1-4.2) | n/a | 0.5 (0.2-0.9) |
| 1970-1979 | 0.0 (0.0-90.9) | 0.3 (0.0-1.0) | 0.6 (0.2-1.3) | n/a | n/a | 0.4 (0.2-0.9) |
| 1980-1989 | 0.0 (0.0-0.7) | 0.5 (0.2-1.1) | 3.1 (1.0-7.3) | n/a | n/a | 0.6 (0.3-1.1) |
| 1990-1999 | 0.0 (0.0-0.5) | 0.3 (0.0-1.4) | n/a | n/a | n/a | 0.1 (0.0-0.5) |
| **Overall** | **0.0 (0.0-0.2)** | **0.4 (0.2-0.7)** | **0.6 (0.3-0.9)** | **1.6 (1.1-2.3)** | **2.9 (2.1-3.9)** | **0.9 (0.8-1.2)** |

Supplementary Table 29 **Crude incidence rates (95% confidence interval) per 100,000 person-years of all HPV-related invasive cancers among Manitoba males by birth year and age group.**

| Birth year | Age group | | | | | |
|---|---|---|---|---|---|---|
| | 9-17 | 18-30 | 31-50 | 51-64 | ≥65 | Overall |
| 1920-1929 | n/a | n/a | n/a | n/a | 30.9 (25.8-36.8) | 30.9 (25.8-36.8) |
| 1930-1939 | n/a | n/a | n/a | 24.4 (16.9-34.1) | 33.5 (28.9-38.7) | 31.7 (27.6-36.2) |
| 1940-1949 | n/a | n/a | 13.3 (5.7-26.2) | 27.0 (23.4-31.0) | 45.1 (38.2-52.8) | 31.7 (28.5-35.1) |
| 1950-1959 | n/a | n/a | 10.7 (8.6-13.3) | 27.3 (23.9-31.0) | 51.9 (20.9-107.0) | 19.7 (17.6-21.9) |
| 1960-1969 | n/a | 0.0 (0.0-5.9) | 4.7 (3.7-5.9) | 15.6 (10.3-22.7) | n/a | 5.6 (4.5-6.8) |
| 1970-1979 | 0.0 (0.0-90.9) | 0.6 (0.2-1.5) | 2.9 (1.9-4.3) | n/a | n/a | 1.9 (1.3-2.7) |
| 1980-1989 | 0.0 (0.0-0.7) | 0.2 (0.0-0.7) | 4.4 (1.8-9.0) | n/a | n/a | 0.5 (0.2-1.0) |
| 1990-1999 | 0.0 (0.0-0.5) | 0.3 (0.0-1.4) | n/a | n/a | n/a | 0.1 (0.0-0.5) |
| **Overall** | **0.0 (0.0-0.2)** | **0.3 (0.1-0.7)** | **5.8 (5.0-6.6)** | **25.9 (23.7-28.3)** | **35.6 (32.6-38.8)** | **11.6 (11.0-12.3)** |

Supplementary Table 30 **Crude incidence rates (95% confidence interval) per 100,000 person-years of invasive cancer of the anus among Manitoba males by birth year and age group.**

| Birth year | Age group | | | | | |
|---|---|---|---|---|---|---|
| | 9-17 | 18-30 | 31-50 | 51-64 | ≥65 | Overall |
| 1920-1929 | n/a | n/a | n/a | n/a | 1.4 (0.5-3.2) | 1.4 (0.5-3.2) |
| 1930-1939 | n/a | n/a | n/a | 2.2 (0.4-6.3) | 2.0 (1.0-3.6) | 2.0 (1.1-3.4) |
| 1940-1949 | n/a | n/a | 0.0 (0.0-6.1) | 1.3 (0.6-2.5) | 1.8 (0.6-3.8) | 1.4 (0.8-2.3) |
| 1950-1959 | n/a | n/a | 0.5 (0.1-1.3) | 2.4 (1.5-3.7) | 0.0 (0.0-27.4) | 1.5 (1.0-2.2) |
| 1960-1969 | n/a | 0.0 (0.0-5.9) | 0.4 (0.1-0.9) | 1.2 (0.1-4.2) | n/a | 0.5 (0.2-0.9) |
| 1970-1979 | 0.0 (0.0-90.9) | 0.1 (0.0-0.8) | 0.2 (0.0-0.8) | n/a | n/a | 0.2 (0.0-0.6) |
| 1980-1989 | 0.0 (0.0-0.7) | 0.0 (0.0-0.4) | 0.6 (0.0-3.5) | n/a | n/a | 0.1 (0.0-0.3) |
| 1990-1999 | 0.0 (0.0-0.5) | 0.0 (0.0-0.9) | n/a | n/a | n/a | 0.0 (0.0-0.3) |
| **Overall** | **0.0 (0.0-0.2)** | **0.0 (0.0-0.3)** | **0.4 (0.2-0.6)** | **1.9 (1.3-2.6)** | **1.6 (1.1-2.5)** | **0.7 (0.6-0.9)** |

Supplementary Table 31 **Crude incidence rates (95% confidence interval) per 100,000 person-years of invasive cancer of the oral cavity among Manitoba males by birth year and age group.**

| Birth year | Age group | | | | | |
|---|---|---|---|---|---|---|
| | 9-17 | 18-30 | 31-50 | 51-64 | ≥65 | Overall |
| 1920-1929 | n/a | n/a | n/a | n/a | 13.3 (10.0-17.3) | 13.3 (10.0-17.3) |
| 1930-1939 | n/a | n/a | n/a | 11.5 (6.6-18.6) | 12.6 (9.8-15.9) | 12.3 (9.9-15.3) |
| 1940-1949 | n/a | n/a | 8.3 (2.7-19.4) | 10.3 (8.1-12.9) | 16.7 (12.6-21.6) | 12.1 (10.2-14.3) |
| 1950-1959 | n/a | n/a | 3.8 (2.6-5.4) | 8.8 (6.9-11.0) | 14.8 (1.8-53.6) | 6.5 (5.3-7.8) |
| 1960-1969 | n/a | 0.0 (0.0-5.9) | 1.3 (0.8-2.0) | 2.9 (0.9-6.7) | n/a | 1.4 (0.9-2.1) |
| 1970-1979 | 0.0 (0.0-90.9) | 0.1 (0.0-0.8) | 1.3 (0.7-2.3) | n/a | n/a | 0.8 (0.4-1.4) |
| 1980-1989 | 0.0 (0.0-0.7) | 0.2 (0.0-0.7) | 1.9 (0.4-5.5) | n/a | n/a | 0.3 (0.1-0.7) |
| 1990-1999 | 0.0 (0.0-0.5) | 0.0 (0.0-0.9) | n/a | n/a | n/a | 0.0 (0.0-0.3) |
| **Overall** | **0.0 (0.0-0.2)** | **0.1 (0.0-0.4)** | **2.0 (1.6-2.6)** | **9.0 (7.7-10.5)** | **14.0 (12.2-16.1)** | **4.3 (3.9-4.7)** |

Supplementary Table 32 **Crude incidence rates (95% confidence interval) per 100,000 person-years of invasive cancer of the oropharynx among Manitoba males by birth year and age group.**

| Birth year | Age group | | | | | |
|---|---|---|---|---|---|---|
| | 9-17 | 18-30 | 31-50 | 51-64 | ≥65 | Overall |
| 1920-1929 | n/a | n/a | n/a | n/a | 10.9 (7.9-14.5) | 10.9 (7.9-14.5) |
| 1930-1939 | n/a | n/a | n/a | 8.6 (4.4-15.0) | 12.0 (9.3-15.3) | 11.3 (9.0-14.1) |
| 1940-1949 | n/a | n/a | 5.0 (1.0-14.6) | 13.8 (11.2-16.7) | 19.0 (14.7-24.3) | 14.9 (12.7-17.3) |
| 1950-1959 | n/a | n/a | 5.3 (3.8-7.2) | 14.5 (12.1-17.3) | 37.1 (12.0-86.6) | 10.3 (8.9-12.0) |
| 1960-1969 | n/a | 0.0 (0.0-5.9) | 2.7 (2.0-3.7) | 10.4 (6.2-16.4) | n/a | 3.4 (2.6-4.4) |
| 1970-1979 | 0.0 (0.0-90.9) | 0.0 (0.0-0.5) | 1.2 (0.6-2.2) | n/a | n/a | 0.7 (0.3-1.2) |
| 1980-1989 | 0.0 (0.0-0.7) | 0.0 (0.0-0.4) | 1.2 (0.2-4.5) | n/a | n/a | 0.1 (0.0-0.4) |
| 1990-1999 | 0.0 (0.0-0.5) | 0.0 (0.0-0.9) | n/a | n/a | n/a | 0.0 (0.0-0.3) |
| **Overall** | **0.0 (0.0-0.2)** | **0.0 (0.0-0.2)** | **2.9 (2.4-3.5)** | **13.4 (11.8-15.2)** | **13.2 (11.4-15.2)** | **5.2 (4.8-5.7)** |

Supplementary Table 33 **Crude incidence rates (95% confidence interval) per 100,000 person-years of invasive cancer of the penis among Manitoba males by birth year and age group.**

| Birth year | Age group | | | | | |
|---|---|---|---|---|---|---|
| | 9-17 | 18-30 | 31-50 | 51-64 | ≥65 | Overall |
| 1920-1929 | n/a | n/a | n/a | n/a | 5.3 (3.3-8.0) | 5.3 (3.3-8.0) |
| 1930-1939 | n/a | n/a | n/a | 2.2 (0.4-6.3) | 6.9 (4.9-9.5) | 6.0 (4.3-8.1) |
| 1940-1949 | n/a | n/a | 0.0 (0.0-6.1) | 1.6 (0.8-2.8) | 7.6 (5.0-11.2) | 3.3 (2.3-4.5) |
| 1950-1959 | n/a | n/a | 1.1 (0.5-2.2) | 1.6 (0.9-2.7) | 0.0 (0.0-27.4) | 1.4 (0.9-2.1) |
| 1960-1969 | n/a | 0.0 (0.0-5.9) | 0.3 (0.1-0.7) | 1.2 (0.1-4.2) | n/a | 0.3 (0.1-0.7) |
| 1970-1979 | 0.0 (0.0-90.9) | 0.3 (0.0-1.0) | 0.1 (0.0-0.6) | n/a | n/a | 0.2 (0.0-0.6) |
| 1980-1989 | 0.0 (0.0-0.7) | 0.0 (0.0-0.4) | 0.6 (0.0-3.5) | n/a | n/a | 0.1 (0.0-0.3) |
| 1990-1999 | 0.0 (0.0-0.5) | 0.3 (0.0-1.4) | n/a | n/a | n/a | 0.1 (0.0-0.5) |
| **Overall** | **0.0 (0.0-0.2)** | **0.1 (0.0-0.4)** | **0.4 (0.2-0.7)** | **1.6 (1.1-2.3)** | **6.7 (5.5-8.2)** | **1.4 (1.2-1.6)** |

#### Costing

Supplementary Table 34 **Total net direct medical cost[1] of HPV-related diseases (for patients compared to their matches) in 2017 Canadian dollars among Manitoba males (1997-2015)**

| HPV-related disease | Outpatient[2] | Hospital | Prescription drug | Total |
|---|---|---|---|---|
| Anogenital warts | $6,132,360 (43%) | $5,205,323 (37%) | $2,826,318 (20%) | $14,164,001 (100%) |
| All HPV-related carcinoma *in situ* | $86,339 (63%) | $11,247 (8%) | $39,377 (29%) | $136,963 (100%) |
| All HPV-related invasive cancers | $7,041,975 (20%) | $26,714,551 (77%) | $1,148,023 (3%) | $34,904,549 (100%) |
| Invasive cancer of the anus | $503,078 (16%) | $2,186,303 (70%) | $451,911 (14%) | $3,141,291 (100%) |
| Invasive cancer of the oral cavity | $3,419,404 (20%) | $13,703,381 (78%) | $364,917 (2%) | $17,487,701 (100%) |
| Invasive cancer of the oropharynx | $2,744,239 (23%) | $9,080,347 (76%) | $175,920 (1%) | $12,000,506 (100%) |
| Invasive cancer of the penis | $375,255 (16%) | $1,744,521 (77%) | $155,275 (7%) | $2,275,051 (100%) |
| **Total** | **$13,260,674 (27%)** | **$31,931,122 (65%)** | **$4,013,717 (8%)** | **$49,205,513 (100%)** |

[1] The periods for which costs were added were 0 to 365 days after diagnosis for AGW and carcinoma *in situ* and 180 days before to 730 days after diagnosis for invasive cancer; [2] Outpatient cost includes diagnostic and therapeutic procedures.

Supplementary Table 35 **Total net direct medical cost[1] of HPV-related diseases (for patients compared to their matches) in 2017 Canadian dollars among Manitoba males by age group (1997-2015)**

| HPV-related disease | Age (years) | | | | | |
|---|---|---|---|---|---|---|
| | 9 - 17 | 18 - 30 | 31 - 50 | 51 - 64 | ≥65 | Total |
| Anogenital warts | $306,100 (2%) | $4,032,794 (28%) | $6,768,276 (48%) | $2,179,081 (15%) | $877,750 (6%) | $14,164,001 (100%) |
| All HPV-related carcinoma *in situ* | | $772 (1%) | $10,651 (8%) | $65,867 (48%) | $59,674 (44%) | $136,963 (100%) |
| All HPV-related invasive cancers | | $189,539 (1%) | $6,013,725 (17%) | $16,369,060 (47%) | $12,332,225 (35%) | $34,904,549 (100%) |
| Invasive cancer of the anus | | $43,629 (1%) | $754,659 (24%) | $1,369,170 (44%) | $973,833 (31%) | $3,141,291 (100%) |
| Invasive cancer of the oral cavity | | $88,918 (1%) | $2,571,585 (15%) | $7,627,090 (44%) | $7,200,108 (41%) | $17,487,701 (100%) |
| Invasive cancer of the oropharynx | | | $2,433,804 (20%) | $6,968,059 (58%) | $2,598,643 (22%) | $12,000,506 (100%) |
| Invasive cancer of the penis | | $56,992 (3%) | $253,677 (11%) | $404,741 (18%) | $1,559,642 (69%) | $2,275,051 (100%) |
| **Total** | **$306,100 (1%)** | **$4,223,105 (9%)** | **$12,792,651 (26%)** | **$18,614,008 (38%)** | **$13,269,649 (27%)** | **$49,205,513 (100%)** |

[1] The periods for which costs were added were 0 to 365 days after diagnosis for AGW and carcinoma *in situ* and 180 days before to 730 days after diagnosis for invasive cancer

Supplementary Table 36 **Total net direct medical cost[1] of HPV-related diseases (for patients compared to their matches) in 2017 Canadian dollars among Manitoba males by regional health authority of residence (1997-2015)**

| HPV-related disease | Regional Health Authority of Residence | | | | | | |
|---|---|---|---|---|---|---|---|
| | **Winnipeg** | **Interlake-Eastern** | **Northern** | **Southern** | **Prairie Mountain** | **Public Trustee/In CFS care** | **Total** |
| Anogenital warts | $10,291,905 (73%) | $1,110,305 (8%) | $510,831 (4%) | $843,248 (6%) | $1,273,611 (9%) | $134,102 (1%) | $14,164,001 (100%) |
| All HPV-related carcinoma *in situ* | $98,495 (72%) | $4,050 (3%) | $14,074 (10%) | $8,135 (6%) | $12,098 (9%) | $110 (0%) | $136,963 (100%) |
| All HPV-related invasive cancers | $19,359,700 (55%) | $3,386,209 (10%) | $1,453,136 (4%) | $4,295,468 (12%) | $6,208,382 (18%) | $201,653 (1%) | $34,904,549 (100%) |
| Invasive cancer of the anus | $2,057,537 (65%) | $334,783 (11%) | $115,968 (4%) | $214,838 (7%) | $418,166 (13%) | | $3,141,291 (100%) |
| Invasive cancer of the oral cavity | $9,932,019 (57%) | $1,379,913 (8%) | $745,366 (4%) | $2,832,054 (16%) | $2,483,375 (14%) | $114,975 (1%) | $17,487,701 (100%) |
| Invasive cancer of the oropharynx | $6,544,309 (55%) | $1,320,713 (11%) | $512,859 (4%) | $940,596 (8%) | $2,611,438 (22%) | $70,591 (1%) | $12,000,506 (100%) |
| Invasive cancer of the penis | $825,835 (36%) | $350,801 (15%) | $78,943 (3%) | $307,980 (14%) | $695,404 (31%) | $16,088 (1%) | $2,275,051 (100%) |
| **Total** | **$29,750,100 (60%)** | **$4,500,564 (9%)** | **$1,978,042 (4%)** | **$5,146,850 (10%)** | **$7,494,091 (15%)** | **$335,866 (1%)** | **$49,205,513 (100%)** |

[1] The periods for which costs were added were 0 to 365 days after diagnosis for AGW and carcinoma *in situ* and 180 days before to 730 days after diagnosis for invasive cancer

Supplementary Table 37 **Total net direct medical cost[1] of HPV-related diseases (for patients compared to their matches) in 2017 Canadian dollars among Manitoba males by Winnipeg region of residence (1997-2015)**

| HPV-related disease | Winnipeg Region of Residence | | | |
|---|---|---|---|---|
| | **Northern suburbs** | **Inner city** | **Southern suburbs** | **Total** |
| Anogenital warts | $3,113,466 (30%) | $3,479,377 (34%) | $3,695,021 (36%) | $10,287,864 (100%) |
| All HPV-related carcinoma *in situ* | $26,723 (27%) | $15,928 (16%) | $55,844 (57%) | $98,495 (100%) |
| All HPV-related invasive cancers | $4,923,590 (25%) | $5,167,484 (27%) | $9,268,626 (48%) | $19,359,700 (100%) |

| | | | | |
|---|---|---|---|---|
| Invasive cancer of the anus | $248,425 (12%) | $1,004,216 (49%) | $804,896 (39%) | $2,057,537 (100%) |
| Invasive cancer of the oral cavity | $1,961,605 (20%) | $2,925,407 (29%) | $5,045,007 (51%) | $9,932,019 (100%) |
| Invasive cancer of the oropharynx | $2,439,652 (37%) | $936,449 (14%) | $3,168,207 (48%) | $6,544,309 (100%) |
| Invasive cancer of the penis | $273,907 (33%) | $301,412 (36%) | $250,516 (30%) | $825,835 (100%) |
| **Total** | **$8,063,779 (27%)** | **$8,662,790 (29%)** | **$13,019,491 (44%)** | **$29,746,060 (100%)** |

[1] The periods for which costs were added were 0 to 365 days after diagnosis for AGW and carcinoma *in situ* and 180 days before to 730 days after diagnosis for invasive cancer

Supplementary Table 38 **Total net direct medical cost[1] of HPV-related diseases (for patients compared to their matches) in 2017 Canadian dollars among Manitoba males by year of diagnosis (1997-2015)**

| HPV-related disease | Year of diagnosis | | | | | | | | | | | | | | | | | | | |
|---|---|---|---|---|---|---|---|---|---|---|---|---|---|---|---|---|---|---|---|---|
| | 1997 | 1998 | 1999 | 2000 | 2001 | 2002 | 2003 | 2004 | 2005 | 2006 | 2007 | 2008 | 2009 | 2010 | 2011 | 2012 | 2013 | 2014 | 2015 | Total |
| Anogenital warts | $458,210 (3%) | $651,989 (5%) | $384,438 (3%) | $745,979 (5%) | $436,459 (3%) | $223,102 (2%) | $1,458,779 (10%) | $702,599 (5%) | $932,401 (7%) | $175,078 (1%) | $693,234 (5%) | $1,096,989 (8%) | $688,491 (5%) | $582,797 (4%) | $1,225,564 (9%) | $995,934 (7%) | $618,053 (4%) | $928,885 (7%) | $1,165,018 (8%) | $14,164,001 (100%) |
| All HPV-related carcinoma *in situ* | $5,939 (4%) | $11,768 (9%) | $220 (0%) | $11,988 (9%) | $6,049 (4%) | $8,466 (6%) | $5,939 (4%) | $17,573 (13%) | $11,878 (9%) | $6,049 (4%) | $15,157 (11%) | $220 (0%) | $661 (0%) | $4,282 (3%) | $3,720 (3%) | $331 (0%) | $6,380 (5%) | $6,490 (5%) | $13,854 (10%) | $136,963 (100%) |
| All HPV-related invasive cancers | $1,231,228 (4%) | $1,363,438 (4%) | $1,500,223 (4%) | $1,386,815 (4%) | $2,664,828 (8%) | $1,472,793 (4%) | $999,002 (3%) | $2,010,317 (6%) | $1,731,735 (5%) | $1,452,536 (4%) | $2,421,907 (7%) | $2,555,432 (7%) | $2,006,732 (6%) | $2,495,595 (7%) | $2,213,881 (6%) | $1,967,290 (6%) | $1,619,441 (5%) | $1,840,824 (5%) | $1,970,535 (6%) | $34,904,549 (100%) |
| Invasive cancer of the anus | $66,977 (2%) | $57,913 (2%) | $231,475 (7%) | $76,896 (2%) | $43,629 (1%) | $144,177 (5%) | $67,705 (2%) | $275,674 (9%) | $170,611 (5%) | $50,663 (2%) | $408,308 (13%) | $387,996 (12%) | $254,276 (8%) | $134,525 (4%) | $53,809 (2%) | $65,742 (2%) | $444,210 (14%) | $87,628 (3%) | $119,077 (4%) | $3,141,291 (100%) |
| Invasive cancer of the oral cavity | $615,422 (4%) | $625,013 (4%) | $647,398 (4%) | $443,167 (3%) | $769,585 (4%) | $614,283 (4%) | $515,044 (3%) | $1,328,591 (8%) | $1,033,292 (6%) | $915,372 (5%) | $1,045,909 (6%) | $1,727,168 (10%) | $783,459 (4%) | $1,727,418 (10%) | $1,252,047 (7%) | $1,200,011 (7%) | $627,875 (4%) | $643,446 (4%) | $973,200 (6%) | $17,487,701 (100%) |
| Invasive cancer of the oropharynx | $328,640 (3%) | $577,678 (5%) | $506,862 (4%) | $815,650 (7%) | $1,757,935 (15%) | $519,996 (4%) | $334,081 (3%) | $359,525 (3%) | $487,759 (4%) | $473,744 (4%) | $855,394 (7%) | $382,519 (3%) | $861,457 (7%) | $646,991 (5%) | $823,213 (7%) | $632,060 (5%) | $475,021 (4%) | $574,258 (5%) | $587,723 (5%) | $12,000,506 (100%) |
| Invasive cancer of the penis | $220,189 (10%) | $102,834 (5%) | $114,488 (5%) | $51,101 (2%) | $93,679 (4%) | $194,337 (9%) | $82,173 (4%) | $46,526 (2%) | $40,073 (2%) | $12,758 (1%) | $112,296 (5%) | $57,749 (3%) | $107,540 (5%) | $-13,338 (-1%) | $84,811 (4%) | $69,476 (3%) | $72,334 (3%) | $535,492 (24%) | $290,535 (13%) | $2,275,051 (100%) |
| **Total** | **$1,695,377 (3%)** | **$2,027,195 (4%)** | **$1,884,882 (4%)** | **$2,144,782 (4%)** | **$3,107,336 (6%)** | **$1,704,361 (3%)** | **$2,463,720 (5%)** | **$2,730,489 (6%)** | **$2,676,014 (5%)** | **$1,633,664 (3%)** | **$3,130,297 (6%)** | **$3,652,641 (7%)** | **$2,695,885 (5%)** | **$3,082,675 (6%)** | **$3,443,165 (7%)** | **$2,963,555 (6%)** | **$2,243,873 (5%)** | **$2,776,199 (6%)** | **$3,149,406 (6%)** | **$49,205,513 (100%)** |

[1] The periods for which costs were added were 0 to 365 days after diagnosis for AGW and carcinoma *in situ* and 180 days before to 730 days after diagnosis for invasive cancer

Supplementary Table 39 **Average of net direct medical cost of HPV-related diseases (for patients compared to their matches) in 2017 Canadian dollars among Manitoba males by successive 90-day periods before and after diagnosis (1997-2015)**

| **HPV-related disease** | **-360 to -271 days** | **-270 to -181 days** | **-180 to -91 days** | **-90 to -1 days** | **0 to 90 days** | **91 to 180 days** | **181 to 270 days** | **271 to 360 days** | **361 to 450 days** | **451 to 540 days** | **541 to 630 days** | **631 to 720 days** | **721 to 810 days** | **811 to 900 days** | **901 to 990 days** | **991 to 1080 days** |
|---|---|---|---|---|---|---|---|---|---|---|---|---|---|---|---|---|
| Anogenital warts | $73 | $114 | $73 | $117 | $521 | $118 | $138 | $181 | $151 | $75 | $261 | $131 | $133 | $119 | $105 | $79 |
| All HPV-related carcinoma *in situ* | $-199 | $458 | $-207 | $2,221 | $1,303 | $322 | $-171 | $65 | $29 | $503 | $-77 | $1,266 | $2,700 | $608 | $-383 | $92 |
| All HPV-related invasive cancers | $216 | $118 | $-12 | $537 | $15,241 | $2,784 | $4,383 | $2,305 | $1,826 | $1,580 | $1,540 | $263 | $586 | $828 | $1,220 | $1,239 |
| Invasive cancer of the anus | $634 | $979 | $1,331 | $3,830 | $10,731 | $5,053 | $3,595 | $7,544 | $6,828 | $2,313 | $1,541 | $709 | $2,319 | $886 | $2,468 | $664 |
| Invasive cancer of the oral cavity | $560 | $251 | $191 | $874 | $25,338 | $3,196 | $3,925 | $2,683 | $2,077 | $1,316 | $1,054 | $119 | $265 | $1,828 | $961 | $46 |
| Invasive cancer of the oropharynx | $29 | $-19 | $-178 | $-307 | $9,886 | $2,228 | $5,732 | $1,493 | $1,219 | $1,282 | $1,821 | $448 | $515 | $125 | $727 | $567 |
| Invasive cancer of the penis | $-393 | $-260 | $-765 | $842 | $6,048 | $2,337 | $1,200 | $1,303 | $577 | $3,125 | $2,019 | $-225 | $915 | $289 | $3,200 | $7,830 |

Supplementary Table 40 **Median of net direct medical cost of HPV-related diseases (for patients compared to their matches) in 2017 Canadian dollars among Manitoba males by successive 90-day periods before and after diagnosis (1997-2015)**

| HPV-related disease | -360 to -271 days | -270 to -181 days | -180 to -91 days | -90 to -1 days | 0 to 90 days | 91 to 180 days | 181 to 270 days | 271 to 360 days | 361 to 450 days | 451 to 540 days | 541 to 630 days | 631 to 720 days | 721 to 810 days | 811 to 900 days | 901 to 990 days | 991 to 1080 days |
|---|---|---|---|---|---|---|---|---|---|---|---|---|---|---|---|---|
| Anogenital warts | $0 | $0 | $0 | $17 | $168 | $1 | $0 | $0 | $0 | $0 | $0 | $-1 | $-0 | $-1 | $0 | $0 |
| All HPV-related carcinoma *in situ* | $-45 | $0 | $-8 | $160 | $843 | $49 | $3 | $36 | $34 | $-6 | $-11 | $40 | $0 | $11 | $-8 | $0 |
| All HPV-related invasive cancers | $-34 | $-37 | $-34 | $219 | $6,355 | $401 | $211 | $100 | $87 | $47 | $0 | $0 | $0 | $0 | $0 | $0 |
| Invasive cancer of the anus | $49 | $-3 | $116 | $507 | $4,540 | $1,616 | $690 | $687 | $702 | $175 | $110 | $124 | $153 | $1 | $0 | $0 |
| Invasive cancer of the oral cavity | $-16 | $-23 | $-30 | $116 | $13,197 | $249 | $220 | $84 | $79 | $0 | $0 | $0 | $0 | $0 | $0 | $-5 |
| Invasive cancer of the oropharynx | $-45 | $-44 | $-40 | $266 | $5,104 | $474 | $204 | $88 | $73 | $64 | $0 | $0 | $0 | $0 | $0 | $0 |
| Invasive cancer of the penis | $-75 | $-55 | $-39 | $273 | $3,177 | $164 | $28 | $46 | $29 | $86 | $18 | $0 | $0 | $0 | $0 | $0 |

#### Net cost by age

Supplementary Table 41 **Distribution of net direct medical cost[1] of anogenital warts (for patients compared to their matches) in 2017 Canadian dollars among Manitoba males by age group (1997-2015)**

| Age (years) | Average | SD[2] | Median | Q1 - Q3[3] |
|---|---|---|---|---|
| 9 - 17 | $1,004 | $5,558 | $305 | $28 - $926 |
| 18 - 30 | $527 | $5,762 | $237 | $-3 - $707 |
| 31 - 50 | $1,254 | $14,053 | $266 | $-175 - $1,049 |
| 51 - 64 | $1,983 | $13,999 | $465 | $-646 - $2,384 |
| ≥65 | $2,421 | $23,561 | $789 | $-1,269 - $3,807 |
| **Overall** | **$958** | **$10,883** | **$256** | **$-86 - $941** |

[1] The period for which costs were added was 0 to 365 days after diagnosis for AGW; [2] Standard deviation; [3] Q1 and Q3 are the first and third quartile respectively

Supplementary Table 42 **Distribution of net direct medical cost[1] of HPV-related carcinoma *in situ* (for patients compared to their matches) in 2017 Canadian dollars among Manitoba males by age group (1997-2015)**

| Age (years) | Average | SD[2] | Median | Q1 - Q3[3] |
|---|---|---|---|---|
| 18 - 30 | $-804 | $5,429 | $439 | $361 - $2,511 |
| 31 - 50 | $4,098 | $4,721 | $1,421 | $724 - $8,038 |
| 51 - 64 | $2,408 | $3,563 | $2,042 | $559 - $4,464 |
| ≥65 | $-348 | $18,221 | $1,025 | $-584 - $4,641 |
| **Overall** | **$1,466** | **$11,483** | **$1,253** | **$-45 - $4,638** |

[1] The period for which costs were added was 0 to 365 days after diagnosis for carcinoma *in situ*; [2] Standard deviation; [3] Q1 and Q3 are the first and third quartile respectively

Supplementary Table 43 **Distribution of net direct medical cost[1] of HPV-related invasive cancer (for patients compared to their matches) in 2017 Canadian dollars among Manitoba males by age group (1997-2015)**

| Age (years) | Average | SD[2] | Median | Q1 - Q3[3] |
|---|---|---|---|---|
| 18 - 30 | $29,182 | $18,455 | $27,283 | $25,192 - $32,891 |
| 31 - 50 | $32,275 | $46,358 | $18,343 | $6,226 - $44,239 |
| 51 - 64 | $34,883 | $89,325 | $16,538 | $5,357 - $44,567 |
| ≥65 | $24,703 | $61,976 | $14,644 | $620 - $39,968 |

| | | | | |
|---|---|---|---|---|
| **Overall** | **$30,382** | **$72,834** | **$16,411** | **$4,527 - $43,189** |

[1] The period for which costs were added was 180 days before to 730 days after diagnosis for invasive cancer; [2] Standard deviation; [3] Q1 and Q3 are the first and third quartile respectively

Supplementary Table 44 **Distribution of net direct medical cost[1] of invasive cancer of the anus (for patients compared to their matches) in 2017 Canadian dollars among Manitoba males by age group (1997-2015)**

| Age (years) | Average | SD[2] | Median | Q1 - Q3[3] |
|---|---|---|---|---|
| 31 - 50 | $59,252 | $53,357 | $41,240 | $13,041 - $98,003 |
| 51 - 64 | $40,060 | $48,646 | $23,212 | $5,869 - $62,087 |
| ≥65 | $39,966 | $69,811 | $21,659 | $11,627 - $72,841 |
| **Overall** | **$43,629** | **$56,467** | **$23,712** | **$10,213 - $72,533** |

[1] The period for which costs were added was 180 days before to 730 days after diagnosis for invasive cancer; [2] Standard deviation; [3] Q1 and Q3 are the first and third quartile respectively

Supplementary Table 45 **Distribution of net direct medical cost[1] of invasive cancer of the oral cavity (for patients compared to their matches) in 2017 Canadian dollars among Manitoba males by age group (1997-2015)**

| Age (years) | Average | SD[2] | Median | Q1 - Q3[3] |
|---|---|---|---|---|
| 18 - 30 | $44,459 | $16,360 | $44,459 | $32,891 - $56,028 |
| 31 - 50 | $38,115 | $47,299 | $23,903 | $5,194 - $54,841 |
| 51 - 64 | $45,489 | $74,075 | $30,826 | $9,869 - $59,381 |
| ≥65 | $37,076 | $69,133 | $24,857 | $2,546 - $54,060 |
| **Overall** | **$40,669** | **$67,962** | **$27,175** | **$7,307 - $57,202** |

[1] The period for which costs were added was 180 days before to 730 days after diagnosis for invasive cancer; [2] Standard deviation; [3] Q1 and Q3 are the first and third quartile respectively

Supplementary Table 46 **Distribution of net direct medical cost[1] of invasive cancer of the oropharynx (for patients compared to their matches) in 2017 Canadian dollars among Manitoba males by age group (1997-2015)**

| Age (years) | Average | SD[2] | Median | Q1 - Q3[3] |
|---|---|---|---|---|
| 31 - 50 | $26,622 | $45,871 | $17,320 | $6,835 - $29,638 |
| 51 - 64 | $29,455 | $106,293 | $12,868 | $4,589 - $29,543 |
| ≥65 | $12,270 | $60,209 | $9,168 | $-4,552 - $31,184 |
| **Overall** | **$23,530** | **$84,366** | **$12,551** | **$3,683 - $29,873** |

[1] The period for which costs were added was 180 days before to 730 days after diagnosis for invasive cancer; [2] Standard deviation; [3] Q1 and Q3 are the first and third quartile respectively

Supplementary Table 47 **Distribution of net direct medical cost[1] of invasive cancer of the penis (for patients compared to their matches) in 2017 Canadian dollars among Manitoba males by age group (1997-2015)**

| Age (years) | Average | SD[2] | Median | Q1 - Q3[3] |
|---|---|---|---|---|
| 18 - 30 | $18,997 | $12,584 | $25,192 | $4,517 - $27,283 |
| 31 - 50 | $18,246 | $24,334 | $6,893 | $1,308 - $21,636 |
| 51 - 64 | $13,031 | $26,133 | $7,640 | $4,537 - $17,639 |
| ≥65 | $17,278 | $37,488 | $9,852 | $1,127 - $24,807 |
| **Overall** | **$16,486** | **$33,336** | **$9,417** | **$2,806 - $21,572** |

[1] The period for which costs were added was 180 days before to 730 days after diagnosis for invasive cancer; [2] Standard deviation; [3] Q1 and Q3 are the first and third quartile respectively

#### Net cost by regional health authority and Winnipeg region of residence

Supplementary Table 48 **Distribution of net direct medical cost[1] of anogenital warts (for patients compared to their matches) in 2017 Canadian dollars among Manitoba males by regional health authority and Winnipeg region of residence (1997-2015)**

| Region | Average | SD[2] | Median | Q1 - Q3[3] |
|---|---|---|---|---|
| Winnipeg | $976 | $12,207 | $251 | $-94 - $918 |
| Northern suburbs | $1,006 | $10,467 | $254 | $-83 - $927 |
| Inner city | $1,796 | $20,557 | $248 | $-157 - $1,074 |
| Southern suburbs | $670 | $8,629 | $251 | $-90 - $864 |
| Interlake-Eastern | $1,086 | $6,737 | $273 | $-62 - $1,107 |
| Northern | $1,373 | $7,996 | $277 | $-44 - $1,129 |
| Southern | $635 | $4,786 | $276 | $-27 - $884 |
| Prairie Mountain | $852 | $6,604 | $251 | $-81 - $929 |
| Public Trustee/In CFS care | $5,047 | $16,260 | $127 | $-904 - $1,141 |
| **Overall** | **$958** | **$10,883** | **$256** | **$-86 - $941** |

[1] The period for which costs were added was 0 to 365 days after diagnosis for AGW; [2] Standard deviation; [3] Q1 and Q3 are the first and third quartile respectively

Supplementary Table 49 **Distribution of net direct medical cost[1] of HPV-related carcinoma *in situ* (for patients compared to their matches) in 2017 Canadian dollars among Manitoba males by regional health authority and Winnipeg region of residence (1997-2015)**

| Region | Average | SD[2] | Median | Q1 - Q3[3] |
|---|---|---|---|---|
| Winnipeg | $3,345 | $7,490 | $1,843 | $404 - $4,881 |
| Northern suburbs | $4,849 | $7,587 | $1,989 | $697 - $5,260 |
| Inner city | $1,747 | $7,996 | $626 | $-3,466 - $3,922 |
| Southern suburbs | $3,125 | $7,327 | $2,097 | $310 - $4,495 |
| Interlake-Eastern | $-1,909 | $14,044 | $398 | $-351 - $4,003 |
| Northern | $3,840 | $5,604 | $1,659 | $361 - $4,296 |
| Southern | $-24 | $2,916 | $-584 | $-2,620 - $3,131 |
| Prairie Mountain | $-11,468 | $31,921 | $957 | $-29,690 - $6,754 |
| Public Trustee/In CFS care | S | S | S | S |
| **Overall** | **$1,466** | **$11,483** | **$1,253** | **$-45 - $4,638** |

[1] The period for which costs were added was 0 to 365 days after diagnosis for carcinoma *in situ*; [2] Standard deviation; [3] Q1 and Q3 are the first and third quartile respectively; S = Suppressed

Supplementary Table 50 **Distribution of net direct medical cost[1] of HPV-related invasive cancer (for patients compared to their matches) in 2017 Canadian dollars among Manitoba males by regional health authority and Winnipeg region of residence (1997-2015)**

| Region | Average | SD[2] | Median | Q1 - Q3[3] |
|---|---|---|---|---|
| Winnipeg | $27,698 | $55,513 | $16,538 | $4,299 - $43,078 |
| Northern suburbs | $25,213 | $44,383 | $16,699 | $4,273 - $33,003 |
| Inner city | $38,559 | $83,948 | $29,913 | $9,944 - $65,232 |
| Southern suburbs | $24,687 | $45,324 | $12,820 | $3,576 - $40,018 |
| Interlake-Eastern | $26,191 | $42,774 | $15,460 | $5,136 - $32,411 |
| Northern | $32,670 | $45,228 | $20,044 | $5,357 - $47,807 |
| Southern | $43,908 | $96,860 | $29,986 | $7,374 - $56,805 |
| Prairie Mountain | $38,688 | $128,945 | $12,361 | $4,340 - $42,871 |
| Public Trustee/In CFS care | $1,294 | $36,797 | $-3,688 | $-28,523 - $12,533 |
| **Overall** | **$30,382** | **$72,834** | **$16,411** | **$4,527 - $43,189** |

[1] The period for which costs were added was 180 days before to 730 days after diagnosis for invasive cancer; [2] Standard deviation; [3] Q1 and Q3 are the first and third quartile respectively

Supplementary Table 51 **Distribution of net direct medical cost[1] of invasive cancer of the anus (for patients compared to their matches) in 2017 Canadian dollars among Manitoba males by regional health authority and Winnipeg region of residence (1997-2015)**

| Region | Average | SD[2] | Median | Q1 - Q3[3] |
|---|---|---|---|---|
| Winnipeg | $41,921 | $56,560 | $25,363 | $10,844 - $69,693 |
| Northern suburbs | $22,584 | $37,865 | $13,037 | $3,887 - $24,076 |
| Inner city | $71,730 | $47,536 | $58,765 | $33,308 - $116,772 |
| Southern suburbs | $32,620 | $63,616 | $22,355 | $3,906 - $49,172 |
| Interlake-Eastern | $41,593 | $59,313 | $11,136 | $8,731 - $79,233 |
| Northern | $36,169 | $32,488 | $36,169 | $13,197 - $59,142 |
| Southern | $57,070 | $56,885 | $70,718 | $-5,398 - $105,889 |
| Prairie Mountain | $57,456 | $76,342 | $21,207 | $10,210 - $74,348 |
| **Overall** | **$43,629** | **$56,467** | **$23,712** | **$10,213 - $72,533** |

[1] The period for which costs were added was 180 days before to 730 days after diagnosis for invasive cancer; [2] Standard deviation; [3] Q1 and Q3 are the first and third quartile respectively

Supplementary Table 52 **Distribution of net direct medical cost[1] of invasive cancer of the oral cavity (for patients compared to their matches) in 2017 Canadian dollars among Manitoba males by regional health authority and Winnipeg region of residence (1997-2015)**

| Region | Average | SD[2] | Median | Q1 - Q3[3] |
|---|---|---|---|---|
| Winnipeg | $37,102 | $53,830 | $26,064 | $6,274 - $54,814 |
| Northern suburbs | $26,824 | $38,840 | $22,158 | $6,658 - $35,040 |
| Inner city | $58,042 | $76,895 | $43,695 | $15,391 - $67,897 |
| Southern suburbs | $33,960 | $47,170 | $18,397 | $4,555 - $54,060 |
| Interlake-Eastern | $27,402 | $38,354 | $21,092 | $5,136 - $34,934 |
| Northern | $54,207 | $61,381 | $35,557 | $17,066 - $61,977 |
| Southern | $70,138 | $120,235 | $45,770 | $30,936 - $73,998 |
| Prairie Mountain | $49,445 | $95,581 | $17,924 | $8,027 - $67,908 |
| Public Trustee/In CFS care | $-1,407 | $44,599 | $-5,290 | $-28,523 - $-3,688 |
| **Overall** | **$40,669** | **$67,962** | **$27,175** | **$7,307 - $57,202** |

[1] The period for which costs were added was 180 days before to 730 days after diagnosis for invasive cancer; [2] Standard deviation; [3] Q1 and Q3 are the first and third quartile respectively

Supplementary Table 53 **Distribution of net direct medical cost[1] of invasive cancer of the oropharynx (for patients compared to their matches) in 2017 Canadian dollars among Manitoba males by regional health authority and Winnipeg region of residence (1997-2015)**

| Region | Average | SD[2] | Median | Q1 - Q3[3] |
|---|---|---|---|---|
| Winnipeg | $20,073 | $58,667 | $12,868 | $3,683 - $29,758 |
| Northern suburbs | $25,734 | $51,148 | $14,052 | $3,786 - $29,543 |
| Inner city | $16,055 | $100,981 | $18,937 | $3,053 - $55,912 |
| Southern suburbs | $18,367 | $40,836 | $10,006 | $3,457 - $26,863 |
| Interlake-Eastern | $23,128 | $45,119 | $13,799 | $4,090 - $28,172 |
| Northern | $23,263 | $29,282 | $14,598 | $2,258 - $38,485 |
| Southern | $21,305 | $81,186 | $11,409 | $-1,648 - $32,010 |
| Prairie Mountain | $43,945 | $185,728 | $11,752 | $4,604 - $31,184 |
| **Overall** | **$23,530** | **$84,366** | **$12,551** | **$3,683 - $29,873** |

[1] The period for which costs were added was 180 days before to 730 days after diagnosis for invasive cancer; [2] Standard deviation; [3] Q1 and Q3 are the first and third quartile respectively

Supplementary Table 54 **Distribution of net direct medical cost[1] of invasive cancer of the penis (for patients compared to their matches) in 2017 Canadian dollars among Manitoba males by regional health authority and Winnipeg region of residence (1997-2015)**

| Region | Average | SD[2] | Median | Q1 - Q3[3] |
|---|---|---|---|---|
| Winnipeg | $13,220 | $28,028 | $9,994 | $2,864 - $20,996 |
| Northern suburbs | $17,210 | $29,165 | $16,672 | $2,864 - $32,932 |
| Inner city | $19,174 | $22,974 | $12,495 | $4,537 - $20,996 |
| Southern suburbs | $6,891 | $30,058 | $7,359 | $-2,286 - $16,718 |
| Interlake-Eastern | $26,486 | $38,005 | $12,239 | $5,782 - $23,217 |
| Northern | $3,250 | $23,723 | $9,801 | $-12,874 - $19,373 |
| Southern | $25,001 | $30,176 | $16,264 | $1,127 - $66,345 |
| Prairie Mountain | $16,999 | $40,373 | $9,048 | $-726 - $21,572 |
| Public Trustee/In CFS care | $8,044 | $6,349 | $8,044 | $3,555 - $12,533 |
| **Overall** | **$16,486** | **$33,336** | **$9,417** | **$2,806 - $21,572** |

[1] The period for which costs were added was 180 days before to 730 days after diagnosis for invasive cancer; [2] Standard deviation; [3] Q1 and Q3 are the first and third quartile respectively

#### Net cost by year of diagnosis

Supplementary Table 55 **Distribution of net direct medical cost[1] of anogenital warts (for patients compared to their matches) in 2017 Canadian dollars among Manitoba males by year of diagnosis (1997-2015)**

| Year of diagnosis | Average | SD[2] | Median | Q1 - Q3[3] |
|---|---|---|---|---|
| 1997 | $588 | $10,700 | $232 | $-21 - $1,052 |
| 1998 | $1,005 | $4,622 | $205 | $-88 - $1,160 |
| 1999 | $653 | $4,085 | $173 | $-100 - $797 |
| 2000 | $1,200 | $17,041 | $238 | $-61 - $1,019 |
| 2001 | $728 | $5,503 | $197 | $-94 - $732 |
| 2002 | $312 | $4,756 | $176 | $-138 - $695 |
| 2003 | $2,044 | $26,449 | $202 | $-105 - $759 |
| 2004 | $928 | $7,921 | $264 | $-28 - $878 |
| 2005 | $1,163 | $17,280 | $203 | $-112 - $702 |
| 2006 | $217 | $7,553 | $183 | $-169 - $660 |
| 2007 | $948 | $8,499 | $234 | $-93 - $821 |
| 2008 | $1,329 | $9,361 | $242 | $-137 - $835 |
| 2009 | $817 | $4,946 | $278 | $-85 - $918 |
| 2010 | $633 | $4,565 | $342 | $-88 - $1,052 |
| 2011 | $1,338 | $13,543 | $345 | $-11 - $1,011 |
| 2012 | $1,067 | $6,414 | $315 | $-104 - $975 |
| 2013 | $743 | $8,297 | $347 | $-75 - $1,085 |
| 2014 | $1,025 | $8,102 | $391 | $-30 - $1,196 |
| 2015 | $1,292 | $10,625 | $327 | $-89 - $1,258 |
| **Overall** | **$958** | **$10,883** | **$256** | **$-86 - $941** |

[1] The period for which costs were added was 0 to 365 days after diagnosis for AGW; [2] Standard deviation; [3] Q1 and Q3 are the first and third quartile respectively

Supplementary Table 56 **Distribution of net direct medical cost[1] of HPV-related carcinoma *in situ* (for patients compared to their matches) in 2017 Canadian dollars among Manitoba males by year of diagnosis (1997-2015)**

| Year of diagnosis | Average | SD[2] | Median | Q1 - Q3[3] |
|---|---|---|---|---|
| 1997 | $1,791 | $1,019 | $1,791 | $1,071 - $2,511 |
| 1998 | $3,550 | $4,317 | $3,203 | $-584 - $8,030 |
| 1999 | $2,295 | $2,002 | $2,295 | $879 - $3,711 |
| 2000 | $3,911 | $4,858 | $2,168 | $799 - $7,022 |
| 2001 | $1,914 | $2,919 | $697 | $-200 - $5,244 |
| 2002 | $-4,986 | $16,571 | $923 | $-1,051 - $3,554 |
| 2003 | $2,513 | $4,400 | $2,513 | $-598 - $5,624 |
| 2004 | $2,107 | $4,745 | $2,097 | $-134 - $4,085 |
| 2005 | $789 | $4,295 | $525 | $-2,092 - $3,671 |
| 2006 | $-80 | $5,389 | $2,096 | $-6,217 - $3,880 |
| 2007 | $3,571 | $3,601 | $3,014 | $678 - $6,463 |
| 2008 | $14,547 | $13,651 | $14,547 | $4,894 - $24,199 |
| 2009 | $3,964 | $22,050 | $5,560 | $1,025 - $14,854 |
| 2010 | $-14,148 | $29,770 | $101 | $-29,492 - $1,197 |
| 2011 | $205 | $4,717 | $439 | $-84 - $1,025 |
| 2012 | $1,929 | $2,436 | $1,218 | $-72 - $4,641 |
| 2013 | $4,063 | $15,969 | $1,192 | $-2,765 - $3,131 |
| 2014 | $2,898 | $3,575 | $1,019 | $-226 - $6,799 |
| 2015 | $3,610 | $4,200 | $2,248 | $429 - $4,433 |
| **Overall** | **$1,466** | **$11,483** | **$1,253** | **$-45 - $4,638** |

[1] The period for which costs were added was 0 to 365 days after diagnosis for carcinoma *in situ*; [2] Standard deviation; [3] Q1 and Q3 are the first and third quartile respectively

Supplementary Table 57 **Distribution of net direct medical cost[1] of HPV-related invasive cancer (for patients compared to their matches) in 2017 Canadian dollars among Manitoba males by year of diagnosis (1997-2015)**

| Year of diagnosis | Average | SD[2] | Median | Q1 - Q3[3] |
|---|---|---|---|---|
| 1997 | $24,270 | $37,416 | $19,242 | $1,750 - $34,934 |
| 1998 | $28,495 | $49,487 | $21,016 | $5,484 - $36,891 |
| 1999 | $33,631 | $62,066 | $15,162 | $1,308 - $54,032 |
| 2000 | $25,505 | $47,256 | $21,636 | $3,669 - $42,209 |
| 2001 | $48,384 | $208,928 | $17,944 | $3,231 - $42,871 |
| 2002 | $26,482 | $34,182 | $22,024 | $6,444 - $50,453 |
| 2003 | $18,177 | $88,144 | $20,556 | $6,274 - $43,817 |
| 2004 | $38,038 | $64,373 | $21,292 | $4,736 - $45,574 |
| 2005 | $36,816 | $47,412 | $20,066 | $7,130 - $58,227 |
| 2006 | $25,963 | $42,424 | $12,700 | $165 - $33,308 |
| 2007 | $38,350 | $74,064 | $17,718 | $4,096 - $47,807 |
| 2008 | $39,127 | $80,999 | $16,002 | $4,107 - $61,977 |
| 2009 | $32,683 | $60,373 | $18,937 | $4,254 - $44,128 |
| 2010 | $38,490 | $99,302 | $19,048 | $5,045 - $43,131 |
| 2011 | $27,433 | $36,223 | $14,385 | $4,902 - $42,561 |
| 2012 | $32,273 | $58,507 | $11,996 | $7,046 - $30,771 |
| 2013 | $24,051 | $37,912 | $17,283 | $5,705 - $43,135 |
| 2014 | $24,516 | $39,853 | $14,452 | $4,259 - $47,549 |
| 2015 | $19,897 | $31,470 | $12,295 | $3,950 - $31,498 |
| **Overall** | **$30,382** | **$72,834** | **$16,411** | **$4,527 - $43,189** |

[1] The period for which costs were added was 180 days before to 730 days after diagnosis for invasive cancer; [2] Standard deviation; [3] Q1 and Q3 are the first and third quartile respectively

Supplementary Table 58 **Distribution of net direct medical cost[1] of invasive cancer of the anus (for patients compared to their matches) in 2017 Canadian dollars among Manitoba males by year of diagnosis (1997-2015)**

| Year of diagnosis | Average | SD[2] | Median | Q1 - Q3[3] |
|---|---|---|---|---|
| 1997 | S | S | S | S |
| 1998 | $19,304 | $17,978 | $14,946 | $3,906 - $39,061 |
| 1999 | $77,158 | $62,950 | $79,233 | $13,197 - $139,045 |
| 2000 | S | S | S | S |
| 2002 | $50,274 | $58,751 | $50,274 | $8,731 - $91,817 |
| 2003 | S | S | S | S |
| 2004 | $55,135 | $67,206 | $44,567 | $11,136 - $49,172 |
| 2005 | $63,491 | $75,351 | $63,491 | $10,210 - $116,772 |
| 2006 | $16,888 | $15,179 | $13,985 | $3,370 - $33,308 |
| 2007 | $68,051 | $86,985 | $22,064 | $3,887 - $162,416 |
| 2008 | $77,599 | $81,087 | $25,363 | $21,683 - $146,443 |
| 2009 | $63,569 | $56,562 | $62,118 | $15,454 - $111,684 |
| 2010 | $33,631 | $15,111 | $30,410 | $21,421 - $45,841 |
| 2011 | $2,545 | $9,669 | $1,543 | $-5,670 - $10,760 |
| 2012 | $32,871 | $28,105 | $32,871 | $12,998 - $52,745 |
| 2013 | $63,459 | $35,796 | $65,033 | $28,606 - $105,631 |
| 2014 | $8,800 | $95,505 | $70,718 | $-36,877 - $74,348 |
| 2015 | $19,846 | $30,742 | $9,710 | $2,925 - $18,377 |
| **Overall** | **$43,629** | **$56,467** | **$23,712** | **$10,213 - $72,533** |

[1] The period for which costs were added was 180 days before to 730 days after diagnosis for invasive cancer; [2] Standard deviation; [3] Q1 and Q3 are the first and third quartile respectively; S = Suppressed

Supplementary Table 59 **Distribution of net direct medical cost[1] of invasive cancer of the oral cavity (for patients compared to their matches) in 2017 Canadian dollars among Manitoba males by year of diagnosis (1997-2015)**

| Year of diagnosis | Average | SD[2] | Median | Q1 - Q3[3] |
|---|---|---|---|---|
| 1997 | $21,847 | $33,640 | $20,146 | $539 - $34,934 |
| 1998 | $24,334 | $37,284 | $20,339 | $4,918 - $26,062 |
| 1999 | $31,933 | $79,155 | $15,470 | $335 - $47,945 |
| 2000 | $14,989 | $33,380 | $6,392 | $-580 - $18,573 |
| 2001 | $31,458 | $37,492 | $22,937 | $3,112 - $47,548 |
| 2002 | $25,684 | $42,109 | $27,341 | $3,108 - $52,256 |
| 2003 | $28,914 | $35,140 | $31,332 | $16,367 - $43,866 |
| 2004 | $73,368 | $84,663 | $30,992 | $17,534 - $85,703 |
| 2005 | $52,886 | $40,064 | $49,382 | $16,501 - $78,667 |
| 2006 | $39,716 | $39,040 | $28,578 | $9,818 - $70,880 |
| 2007 | $51,328 | $74,801 | $26,554 | $8,826 - $72,532 |
| 2008 | $78,373 | $111,950 | $61,977 | $16,930 - $93,706 |
| 2009 | $30,532 | $53,812 | $20,174 | $2,147 - $48,896 |
| 2010 | $59,460 | $140,853 | $29,892 | $8,027 - $64,587 |
| 2011 | $46,828 | $44,642 | $42,561 | $14,433 - $70,784 |
| 2012 | $45,102 | $82,013 | $15,167 | $7,248 - $43,695 |
| 2013 | $36,700 | $33,765 | $35,299 | $8,088 - $47,160 |
| 2014 | $34,341 | $34,677 | $19,282 | $10,206 - $47,549 |
| 2015 | $27,949 | $37,108 | $29,790 | $9,247 - $45,632 |
| **Overall** | **$40,669** | **$67,962** | **$27,175** | **$7,307 - $57,202** |

[1] The period for which costs were added was 180 days before to 730 days after diagnosis for invasive cancer; [2] Standard deviation; [3] Q1 and Q3 are the first and third quartile respectively

Supplementary Table 60 **Distribution of net direct medical cost[1] of invasive cancer of the oropharynx (for patients compared to their matches) in 2017 Canadian dollars among Manitoba males by year of diagnosis (1997-2015)**

| Year of diagnosis | Average | SD[2] | Median | Q1 - Q3[3] |
|---|---|---|---|---|
| 1997 | $15,635 | $25,111 | $15,236 | $5,979 - $26,010 |
| 1998 | $34,634 | $66,401 | $28,616 | $9,672 - $53,995 |
| 1999 | $35,369 | $45,167 | $27,997 | $1,182 - $69,854 |
| 2000 | $42,443 | $56,695 | $41,904 | $22,992 - $56,940 |
| 2001 | $77,767 | $310,734 | $21,286 | $2,889 - $43,330 |
| 2002 | $21,400 | $24,544 | $20,030 | $6,444 - $34,012 |
| 2003 | $11,998 | $124,497 | $19,261 | $5,596 - $47,405 |
| 2004 | $10,917 | $28,731 | $14,415 | $-2,114 - $21,970 |
| 2005 | $21,771 | $51,238 | $16,843 | $4,645 - $25,971 |
| 2006 | $17,778 | $47,418 | $6,291 | $-2,372 - $17,476 |
| 2007 | $28,568 | $78,370 | $12,892 | $3,053 - $32,445 |
| 2008 | $9,251 | $34,842 | $10,553 | $-4,552 - $22,945 |
| 2009 | $30,418 | $68,177 | $10,917 | $2,028 - $42,474 |
| 2010 | $20,232 | $28,951 | $11,153 | $4,694 - $28,693 |
| 2011 | $18,433 | $27,674 | $7,806 | $4,587 - $24,514 |
| 2012 | $23,389 | $31,277 | $11,593 | $4,344 - $29,873 |
| 2013 | $13,805 | $36,676 | $10,402 | $709 - $27,071 |
| 2014 | $17,402 | $23,568 | $11,673 | $2,954 - $20,254 |
| 2015 | $15,671 | $19,315 | $9,123 | $4,740 - $18,613 |
| **Overall** | **$23,530** | **$84,366** | **$12,551** | **$3,683 - $29,873** |

[1] The period for which costs were added was 180 days before to 730 days after diagnosis for invasive cancer; [2] Standard deviation; [3] Q1 and Q3 are the first and third quartile respectively

Supplementary Table 61 **Distribution of net direct medical cost[1] of invasive cancer of the penis (for patients compared to their matches) in 2017 Canadian dollars among Manitoba males by year of diagnosis (1997-2015)**

| Year of diagnosis | Average | SD[2] | Median | Q1 - Q3[3] |
|---|---|---|---|---|
| 1997 | $110,095 | $68,650 | $110,095 | $61,552 - $158,637 |
| 1998 | S | S | S | S |
| 1999 | $16,355 | $28,533 | $8,161 | $1,308 - $14,854 |
| 2000 | $7,300 | $45,010 | $18,573 | $2,685 - $24,807 |
| 2001 | $10,409 | $28,597 | $7,120 | $4,537 - $27,283 |
| 2002 | $44,463 | $36,508 | $45,229 | $12,920 - $76,006 |
| 2003 | $10,948 | $28,026 | $-92 | $-5,465 - $12,533 |
| 2004 | $9,305 | $12,899 | $4,736 | $3,097 - $5,043 |
| 2005 | $7,862 | $11,432 | $7,467 | $-3,367 - $19,487 |
| 2006 | $4,253 | $6,779 | $3,269 | $-1,981 - $11,469 |
| 2007 | $10,473 | $9,764 | $11,904 | $4,937 - $17,718 |
| 2008 | $3,751 | $6,881 | $4,340 | $-2,146 - $5,616 |
| 2009 | $20,798 | $279 | $20,798 | $20,601 - $20,996 |
| 2010 | $-6,669 | $32,583 | $-6,669 | $-29,709 - $16,370 |
| 2011 | $16,962 | $13,615 | $16,264 | $12,134 - $16,718 |
| 2012 | $13,895 | $10,260 | $9,820 | $9,411 - $15,435 |
| 2013 | $6,205 | $23,502 | $9,417 | $3,555 - $19,514 |
| 2014 | $37,072 | $45,472 | $26,507 | $4,353 - $73,214 |
| 2015 | $14,037 | $39,968 | $7,224 | $-5,487 - $19,021 |
| **Overall** | **$16,486** | **$33,336** | **$9,417** | **$2,806 - $21,572** |

[1] The period for which costs were added was 180 days before to 730 days after diagnosis for invasive cancer; [2] Standard deviation; [3] Q1 and Q3 are the first and third quartile respectively; S = Suppressed

#### Healthcare utilization

Supplementary Table 62 **Average number of healthcare encounters[1] (per year) of HPV-related disease patients and their matches among Manitoba males (1997-2015)**

| **HPV-related disease** | **Patients** | | | | **Matches** | | | |
|---|---|---|---|---|---|---|---|---|
| | **Clinician visits** | **Hospital admissions** | **ER visits[2]** | **Prescriptions** | **Clinician visits** | **Hospital admissions** | **ER visits[2]** | **Prescriptions** |
| Anogenital warts | 8.9 | 0.2 | 0.3 | 8.9 | 4.3 | 0.1 | 0.2 | 6.3 |
| All HPV-related carcinoma *in situ* | 17.9 | 1.1 | 0.5 | 29.2 | 9.5 | 0.3 | 0.4 | 20.2 |
| All HPV-related invasive cancers | 19.9 | 1.3 | 0.8 | 34.8 | 9.0 | 0.3 | 0.3 | 21.4 |
| Invasive cancer of the anus | 24.7 | 1.9 | 2.2 | 94.6 | 8.1 | 0.2 | 0.2 | 20.0 |
| Invasive cancer of the oral cavity | 19.8 | 1.3 | 0.7 | 32.5 | 9.1 | 0.3 | 0.3 | 20.7 |
| Invasive cancer of the oropharynx | 20.0 | 1.3 | 0.6 | 27.0 | 8.7 | 0.3 | 0.3 | 20.0 |
| Invasive cancer of the penis | 17.6 | 1.4 | 0.6 | 36.9 | 10.2 | 0.3 | 0.4 | 29.3 |
| **Total** | **10.5** | **0.4** | **0.4** | **12.6** | **4.9** | **0.1** | **0.2** | **8.4** |

[1] The periods for which encounters were averaged (as number per year) were 0 to 365 days after diagnosis for AGW and carcinoma *in situ* and 180 days before to 730 days after diagnosis for invasive cancer;
[2] ER visits are for the Winnipeg Regional Health Authority only

Supplementary Table 63 **Excess number of healthcare encounters[1] (per year) due to HPV-related diseases (for patients compared to their matches) among Manitoba males (1997-2015)**

| HPV-related disease | Clinician visits | Hospital admissions | ER visits[2] | Prescriptions |
|---|---|---|---|---|
| Anogenital warts | 67,637 | 2,218 | 1,118 | 35,618 |
| All HPV-related carcinoma *in situ* | 733 | 66 | -3 | 811 |
| All HPV-related invasive cancers | 11,665 | 1,174 | 319 | 13,811 |
| Invasive cancer of the anus | 1,167 | 126 | 107 | 5,747 |
| Invasive cancer of the oral cavity | 4,330 | 425 | 88 | 5,107 |
| Invasive cancer of the oropharynx | 5,224 | 478 | 113 | 2,089 |
| Invasive cancer of the penis | 943 | 145 | 11 | 868 |
| **Total** | **80,034** | **3,458** | **1,434** | **50,239** |

[1] The periods for which encounters were averaged (as number per year) were 0 to 365 days after diagnosis for AGW and carcinoma *in situ* and 180 days before to 730 days after diagnosis for invasive cancer;
[2] ER visits are for the Winnipeg Regional Health Authority only

Supplementary Table 64 **Distribution of excess number of healthcare encounters[1] (per year) due to HPV-related diseases (for patients compared to their matches) among Manitoba males (1997-2015)**

| HPV-related disease | Average | SD[2] | Median | Q1 - Q3[3] |
|---|---|---|---|---|
| **Excess clinician visits** | | | | |
| Anogenital warts | 4.6 | 9.7 | 3.0 | -0.3 - 7.7 |
| All HPV-related carcinoma *in situ* | 8.6 | 11.1 | 4.8 | 1.2 - 15.8 |
| All HPV-related invasive cancers | 10.2 | 15.4 | 8.8 | 3.0 - 16.7 |
| Invasive cancer of the anus | 16.2 | 20.6 | 14.4 | 6.9 - 23.6 |
| Invasive cancer of the oral cavity | 10.1 | 16.2 | 8.1 | 2.1 - 15.5 |
| Invasive cancer of the oropharynx | 10.2 | 13.8 | 9.3 | 3.7 - 17.5 |
| Invasive cancer of the penis | 6.8 | 14.7 | 6.5 | 0.7 - 12.9 |
| **Excess hospital admissions** | | | | |
| Anogenital warts | 0.2 | 0.7 | 0.0 | 0.0 - 0.0 |
| All HPV-related carcinoma *in situ* | 0.8 | 1.0 | 0.7 | 0.0 - 1.3 |
| All HPV-related invasive cancers | 1.0 | 1.2 | 0.8 | 0.4 - 1.6 |
| Invasive cancer of the anus | 1.8 | 1.6 | 1.4 | 0.7 - 2.4 |
| Invasive cancer of the oral cavity | 1.0 | 1.1 | 0.8 | 0.4 - 1.6 |
| Invasive cancer of the oropharynx | 0.9 | 1.2 | 0.7 | 0.3 - 1.4 |
| Invasive cancer of the penis | 1.0 | 1.0 | 0.8 | 0.4 - 1.6 |
| **Excess ER visits[4]** | | | | |
| Anogenital warts | 0.1 | 1.2 | 0.0 | -0.3 - 0.0 |
| All HPV-related carcinoma *in situ* | -0.0 | 1.3 | 0.0 | -0.3 - 0.0 |
| All HPV-related invasive cancers | 0.5 | 2.1 | 0.0 | -0.1 - 0.5 |
| Invasive cancer of the anus | 2.2 | 6.2 | 0.4 | 0.0 - 1.5 |
| Invasive cancer of the oral cavity | 0.3 | 1.4 | 0.0 | -0.1 - 0.5 |
| Invasive cancer of the oropharynx | 0.4 | 1.2 | 0.0 | -0.1 - 0.4 |
| Invasive cancer of the penis | 0.2 | 0.8 | 0.0 | -0.1 - 0.4 |
| **Excess prescriptions** | | | | |
| Anogenital warts | 2.4 | 25.9 | 0.0 | -2.3 - 4.0 |
| All HPV-related carcinoma *in situ* | 9.6 | 49.6 | 1.8 | -10.0 - 14.2 |
| All HPV-related invasive cancers | 12.2 | 91.5 | 6.0 | -7.9 - 24.6 |
| Invasive cancer of the anus | 79.8 | 266.5 | 9.2 | -1.5 - 46.8 |
| Invasive cancer of the oral cavity | 11.9 | 67.3 | 6.8 | -7.9 - 25.0 |
| Invasive cancer of the oropharynx | 4.1 | 58.5 | 4.9 | -7.1 - 20.9 |
| Invasive cancer of the penis | 6.3 | 50.6 | 1.2 | -15.1 - 28.0 |

[1] The periods for which encounters were averaged (as number per year) were 0 to 365 days after diagnosis for AGW and carcinoma *in situ* and 180 days before to 730 days after diagnosis for invasive cancer; [2] Standard deviation; [3] Q1 and Q3 are the first and third quartile respectively; [4] ER visits are for the Winnipeg Regional Health Authority only

#### Excess healthcare utilization by age

Supplementary Table 65 **Distribution of excess number of clinician visits[1] (per year) due to anogenital warts (for patients compared to their matches) among Manitoba males by age group (1997-2015).**

| Age (years) | Average | SD[2] | Median | Q1 - Q3[3] |
|---|---|---|---|---|
| 9 - 17 | 3.5 | 5.8 | 2.3 | 0.0 - 6.0 |
| 18 - 30 | 4.2 | 7.4 | 3.0 | 0.3 - 6.7 |
| 31 - 50 | 4.7 | 10.5 | 2.7 | -0.7 - 8.3 |
| 51 - 64 | 6.2 | 14.9 | 4.0 | -1.0 - 11.0 |
| ≥65 | 6.7 | 18.0 | 5.3 | -1.3 - 14.0 |
| **Overall** | **4.6** | **9.7** | **3.0** | **-0.3 - 7.7** |

[1] The period for which encounters were averaged (as number per year) was 0 to 365 days after diagnosis for AGW; [2] Standard deviation; [3] Q1 and Q3 are the first and third quartile respectively

Supplementary Table 66 **Distribution of excess number of hospital admissions[1] (per year) due to anogenital warts (for patients compared to their matches) among Manitoba males by age group (1997-2015).**

| Age (years) | Average | SD[2] | Median | Q1 - Q3[3] |
|---|---|---|---|---|
| 9 - 17 | 0.2 | 0.6 | 0.0 | 0.0 - 0.0 |
| 18 - 30 | 0.1 | 0.5 | 0.0 | 0.0 - 0.0 |
| 31 - 50 | 0.2 | 0.7 | 0.0 | 0.0 - 0.0 |
| 51 - 64 | 0.3 | 1.1 | 0.0 | 0.0 - 0.7 |
| ≥65 | 0.6 | 1.3 | 0.0 | 0.0 - 1.0 |
| **Overall** | **0.2** | **0.7** | **0.0** | **0.0 - 0.0** |

[1] The period for which encounters were averaged (as number per year) was 0 to 365 days after diagnosis for AGW; [2] Standard deviation; [3] Q1 and Q3 are the first and third quartile respectively

Supplementary Table 67 **Distribution of excess number of ER visits[1,2] (per year) due to anogenital warts (for patients compared to their matches) among Manitoba males by age group (1997-2015).**

| Age (years) | Average | SD[3] | Median | Q1 - Q3[4] |
|---|---|---|---|---|
| 9 - 17 | 0.1 | 1.0 | 0.0 | -0.3 - 0.0 |
| 18 - 30 | 0.1 | 1.0 | 0.0 | -0.3 - 0.0 |
| 31 - 50 | 0.1 | 1.3 | 0.0 | -0.3 - 0.0 |

| | | | | |
|---|---|---|---|---|
| 51 - 64 | 0.1 | 1.1 | 0.0 | -0.3 - 0.0 |
| ≥65 | 0.2 | 3.1 | 0.0 | -0.3 - 0.0 |
| **Overall** | **0.1** | **1.2** | **0.0** | **-0.3 - 0.0** |

[1] ER visits are for the Winnipeg Regional Health Authority only; [2] The period for which encounters were averaged (as number per year) was 0 to 365 days after diagnosis for AGW; [3] Standard deviation; [4] Q1 and Q3 are the first and third quartile respectively

Supplementary Table 68 **Distribution of excess number of prescriptions[1] (per year) due to anogenital warts (for patients compared to their matches) among Manitoba males by age group (1997-2015).**

| Age (years) | Average | SD[2] | Median | Q1 - Q3[3] |
|---|---|---|---|---|
| 9 - 17 | 0.9 | 9.5 | 0.0 | -1.7 - 2.7 |
| 18 - 30 | 1.0 | 14.0 | 0.0 | -1.7 - 3.0 |
| 31 - 50 | 3.4 | 29.9 | 0.0 | -3.3 - 5.3 |
| 51 - 64 | 7.1 | 44.0 | 1.0 | -8.7 - 15.0 |
| ≥65 | 4.5 | 61.4 | 2.0 | -12.3 - 25.7 |
| **Overall** | **2.4** | **25.9** | **0.0** | **-2.3 - 4.0** |

[1] The period for which encounters were averaged (as number per year) was 0 to 365 days after diagnosis for AGW; [2] Standard deviation; [3] Q1 and Q3 are the first and third quartile respectively

Supplementary Table 69 **Distribution of excess number of clinician visits[1] (per year) due to HPV-related carcinoma *in situ* (for patients compared to their matches) among Manitoba males by age group (1997-2015).**

| Age (years) | Average | SD[2] | Median | Q1 - Q3[3] |
|---|---|---|---|---|
| 18 - 30 | 5.3 | 7.9 | 4.3 | 1.7 - 9.0 |
| 31 - 50 | 8.8 | 11.8 | 4.3 | 1.3 - 14.5 |
| 51 - 64 | 5.2 | 7.7 | 4.0 | -0.8 - 11.5 |
| ≥65 | 12.6 | 13.2 | 14.0 | 3.3 - 20.7 |
| **Overall** | **8.6** | **11.1** | **4.8** | **1.2 - 15.8** |

[1] The period for which encounters were averaged (as number per year) was 0 to 365 days after diagnosis for carcinoma *in situ*; [2] Standard deviation; [3] Q1 and Q3 are the first and third quartile respectively

Supplementary Table 70 **Distribution of excess number of hospital admissions[1] (per year) due to HPV-related carcinoma *in situ* (for patients compared to their matches) among Manitoba males by age group (1997-2015).**

| Age (years) | Average | SD[2] | Median | Q1 - Q3[3] |
|---|---|---|---|---|

| | | | | |
|---|---|---|---|---|
| 18 - 30 | 0.3 | 0.9 | 0.0 | 0.0 - 1.0 |
| 31 - 50 | 0.6 | 0.9 | 0.0 | 0.0 - 1.5 |
| 51 - 64 | 0.8 | 0.8 | 0.7 | 0.3 - 1.0 |
| ≥65 | 0.9 | 1.3 | 1.0 | -0.3 - 1.7 |
| **Overall** | **0.8** | **1.0** | **0.7** | **0.0 - 1.3** |

[1] The period for which encounters were averaged (as number per year) was 0 to 365 days after diagnosis for carcinoma *in situ*; [2] Standard deviation; [3] Q1 and Q3 are the first and third quartile respectively

Supplementary Table 71 **Distribution of excess number of ER visits[1,2] (per year) due to HPV-related carcinoma *in situ* (for patients compared to their matches) among Manitoba males by age group (1997-2015).**

| Age (years) | Average | SD[3] | Median | Q1 - Q3[4] |
|---|---|---|---|---|
| 18 - 30 | 0.1 | 2.3 | -0.7 | -0.7 - -0.3 |
| 31 - 50 | 0.3 | 0.9 | 0.0 | 0.0 - 0.7 |
| 51 - 64 | -0.2 | 0.5 | 0.0 | -0.3 - 0.0 |
| ≥65 | -0.1 | 1.8 | 0.0 | -0.8 - 0.8 |
| **Overall** | **-0.0** | **1.3** | **0.0** | **-0.3 - 0.0** |

[1] ER visits are for the Winnipeg Regional Health Authority only; [2] The period for which encounters were averaged (as number per year) was 0 to 365 days after diagnosis for carcinoma *in situ*; [3] Standard deviation; [4] Q1 and Q3 are the first and third quartile respectively

Supplementary Table 72 **Distribution of excess number of prescriptions[1] (per year) due to HPV-related carcinoma *in situ* (for patients compared to their matches) among Manitoba males by age group (1997-2015).**

| Age (years) | Average | SD[2] | Median | Q1 - Q3[3] |
|---|---|---|---|---|
| 18 - 30 | -7.0 | 25.1 | 0.7 | -1.0 - 4.3 |
| 31 - 50 | 21.8 | 41.7 | 5.2 | 0.2 - 23.7 |
| 51 - 64 | 9.2 | 39.9 | -0.5 | -10.0 - 7.0 |
| ≥65 | 7.2 | 64.6 | 4.3 | -23.0 - 27.7 |
| **Overall** | **9.6** | **49.6** | **1.8** | **-10.0 - 14.2** |

[1] The period for which encounters were averaged (as number per year) was 0 to 365 days after diagnosis for carcinoma *in situ*; [2] Standard deviation; [3] Q1 and Q3 are the first and third quartile respectively

Supplementary Table 73 **Distribution of excess number of clinician visits[1] (per year) due to HPV-related invasive cancer (for patients compared to their matches) among Manitoba males by age group (1997-2015).**

| Age (years) | Average | SD[2] | Median | Q1 - Q3[3] |
|---|---|---|---|---|
| 18 - 30 | 12.6 | 8.0 | 6.9 | 6.9 - 18.5 |
| 31 - 50 | 12.6 | 16.9 | 9.5 | 4.4 - 17.5 |
| 51 - 64 | 10.5 | 12.8 | 9.2 | 3.6 - 16.6 |
| ≥65 | 8.7 | 17.2 | 7.9 | 1.5 - 15.6 |
| **Overall** | **10.2** | **15.4** | **8.8** | **3.0 - 16.7** |

[1] The periods for which encounters were averaged (as number per year) was 180 days before to 730 days after diagnosis for invasive cancer; [2] Standard deviation; [3] Q1 and Q3 are the first and third quartile respectively

Supplementary Table 74 **Distribution of excess number of hospital admissions[1] (per year) due to HPV-related invasive cancer (for patients compared to their matches) among Manitoba males by age group (1997-2015).**

| Age (years) | Average | SD[2] | Median | Q1 - Q3[3] |
|---|---|---|---|---|
| 18 - 30 | 1.1 | 0.6 | 1.1 | 0.8 - 1.6 |
| 31 - 50 | 1.1 | 1.2 | 0.8 | 0.4 - 1.5 |
| 51 - 64 | 1.0 | 1.0 | 0.8 | 0.4 - 1.5 |
| ≥65 | 1.0 | 1.3 | 0.8 | 0.2 - 1.6 |
| **Overall** | **1.0** | **1.2** | **0.8** | **0.4 - 1.6** |

[1] The periods for which encounters were averaged (as number per year) was 180 days before to 730 days after diagnosis for invasive cancer; [2] Standard deviation; [3] Q1 and Q3 are the first and third quartile respectively

Supplementary Table 75 **Distribution of excess number of ER visits[1,2] (per year) due to HPV-related invasive cancer (for patients compared to their matches) among Manitoba males by age group (1997-2015).**

| Age (years) | Average | SD[3] | Median | Q1 - Q3[4] |
|---|---|---|---|---|
| 18 - 30 | 1.2 | 1.3 | 1.2 | 0.3 - 2.1 |
| 31 - 50 | 0.7 | 3.9 | 0.0 | -0.1 - 0.4 |
| 51 - 64 | 0.4 | 1.4 | 0.1 | -0.1 - 0.5 |
| ≥65 | 0.4 | 1.7 | 0.0 | -0.1 - 0.8 |
| **Overall** | **0.5** | **2.1** | **0.0** | **-0.1 - 0.5** |

[1] ER visits are for the Winnipeg Regional Health Authority only; [2] The periods for which encounters were averaged (as number per year) was 180 days before to 730 days after diagnosis for invasive cancer; [3] Standard deviation; [4] Q1 and Q3 are the first and third quartile respectively

Supplementary Table 76 **Distribution of excess number of prescriptions[1] (per year) due to HPV-related invasive cancer (for patients compared to their matches) among Manitoba males by age group (1997-2015).**

| Age (years) | Average | SD[2] | Median | Q1 - Q3[3] |
|---|---|---|---|---|
| 18 - 30 | 13.4 | 11.2 | 16.9 | 1.6 - 21.2 |
| 31 - 50 | 16.2 | 33.7 | 6.2 | -0.4 - 23.5 |
| 51 - 64 | 17.9 | 112.0 | 6.3 | -5.9 - 22.7 |
| ≥65 | 4.1 | 84.2 | 3.6 | -15.6 - 26.5 |
| **Overall** | **12.2** | **91.5** | **6.0** | **-7.9 - 24.6** |

[1] The periods for which encounters were averaged (as number per year) was 180 days before to 730 days after diagnosis for invasive cancer; [2] Standard deviation; [3] Q1 and Q3 are the first and third quartile respectively

Supplementary Table 77 **Distribution of excess number of clinician visits[1] (per year) due to invasive cancer of the anus (for patients compared to their matches) among Manitoba males by age group (1997-2015).**

| Age (years) | Average | SD[2] | Median | Q1 - Q3[3] |
|---|---|---|---|---|
| 31 - 50 | 27.1 | 25.7 | 16.1 | 11.7 - 30.7 |
| 51 - 64 | 13.0 | 12.0 | 10.3 | 5.0 - 18.9 |
| ≥65 | 14.8 | 26.4 | 13.2 | 10.3 - 26.0 |
| **Overall** | **16.2** | **20.6** | **14.4** | **6.9 - 23.6** |

[1] The periods for which encounters were averaged (as number per year) was 180 days before to 730 days after diagnosis for invasive cancer; [2] Standard deviation; [3] Q1 and Q3 are the first and third quartile respectively

Supplementary Table 78 **Distribution of excess number of hospital admissions[1] (per year) due to invasive cancer of the anus (for patients compared to their matches) among Manitoba males by age group (1997-2015).**

| Age (years) | Average | SD[2] | Median | Q1 - Q3[3] |
|---|---|---|---|---|
| 31 - 50 | 2.5 | 2.6 | 1.8 | 0.8 - 2.9 |
| 51 - 64 | 1.5 | 1.3 | 1.3 | 0.5 - 2.4 |
| ≥65 | 1.6 | 1.0 | 1.5 | 0.8 - 2.2 |
| **Overall** | **1.8** | **1.6** | **1.4** | **0.7 - 2.4** |

[1] The periods for which encounters were averaged (as number per year) was 180 days before to 730 days after diagnosis for invasive cancer; [2] Standard deviation; [3] Q1 and Q3 are the first and third quartile respectively

Supplementary Table 79 **Distribution of excess number of ER visits[1,2] (per year) due to invasive cancer of the anus (for patients compared to their matches) among Manitoba males by age group (1997-2015).**

| Age (years) | Average | SD[3] | Median | Q1 - Q3[4] |
|---|---|---|---|---|
| 31 - 50 | 6.1 | 12.5 | 1.5 | 0.0 - 4.0 |
| 51 - 64 | 1.3 | 3.2 | 0.5 | 0.0 - 1.2 |
| ≥65 | 1.2 | 2.7 | 0.4 | 0.1 - 1.2 |
| **Overall** | **2.2** | **6.2** | **0.4** | **0.0 - 1.5** |

[1] ER visits are for the Winnipeg Regional Health Authority only; [2] The periods for which encounters were averaged (as number per year) was 180 days before to 730 days after diagnosis for invasive cancer; [3] Standard deviation; [4] Q1 and Q3 are the first and third quartile respectively

Supplementary Table 80 **Distribution of excess number of prescriptions[1] (per year) due to invasive cancer of the anus (for patients compared to their matches) among Manitoba males by age group (1997-2015).**

| Age (years) | Average | SD[2] | Median | Q1 - Q3[3] |
|---|---|---|---|---|
| 31 - 50 | 56.1 | 58.1 | 38.9 | 8.4 - 94.2 |
| 51 - 64 | 122.7 | 367.8 | 6.5 | -8.1 - 61.6 |
| ≥65 | 25.4 | 79.0 | 8.1 | -0.8 - 27.0 |
| **Overall** | **79.8** | **266.5** | **9.2** | **-1.5 - 46.8** |

[1] The periods for which encounters were averaged (as number per year) was 180 days before to 730 days after diagnosis for invasive cancer; [2] Standard deviation; [3] Q1 and Q3 are the first and third quartile respectively

Supplementary Table 81 **Distribution of excess number of clinician visits[1] (per year) due to invasive cancer of the oral cavity (for patients compared to their matches) among Manitoba males by age group (1997-2015).**

| Age (years) | Average | SD[2] | Median | Q1 - Q3[3] |
|---|---|---|---|---|
| 18 - 30 | 15.2 | 11.9 | 15.2 | 6.8 - 23.6 |
| 31 - 50 | 11.9 | 22.3 | 8.3 | 3.0 - 12.7 |
| 51 - 64 | 9.6 | 13.8 | 8.6 | 2.4 - 16.4 |
| ≥65 | 9.8 | 15.7 | 7.8 | 1.2 - 15.4 |
| **Overall** | **10.1** | **16.2** | **8.1** | **2.1 - 15.5** |

[1] The periods for which encounters were averaged (as number per year) was 180 days before to 730 days after diagnosis for invasive cancer; [2] Standard deviation; [3] Q1 and Q3 are the first and third quartile respectively

Supplementary Table 82 **Distribution of excess number of hospital admissions[1] (per year) due to invasive cancer of the oral cavity (for patients compared to their matches) among Manitoba males by age group (1997-2015).**

| Age (years) | Average | SD[2] | Median | Q1 - Q3[3] |
|---|---|---|---|---|
| 18 - 30 | 1.1 | 0.9 | 1.1 | 0.4 - 1.7 |
| 31 - 50 | 0.9 | 0.9 | 0.7 | 0.4 - 1.3 |
| 51 - 64 | 0.9 | 0.9 | 0.8 | 0.4 - 1.6 |
| ≥65 | 1.1 | 1.4 | 0.8 | 0.3 - 1.6 |
| **Overall** | **1.0** | **1.1** | **0.8** | **0.4 - 1.6** |

[1] The periods for which encounters were averaged (as number per year) was 180 days before to 730 days after diagnosis for invasive cancer; [2] Standard deviation; [3] Q1 and Q3 are the first and third quartile respectively

Supplementary Table 83 **Distribution of excess number of ER visits[1,2] (per year) due to invasive cancer of the oral cavity (for patients compared to their matches) among Manitoba males by age group (1997-2015).**

| Age (years) | Average | SD[3] | Median | Q1 - Q3[4] |
|---|---|---|---|---|
| 18 - 30 | 2.1 | | 2.1 | 2.1 - 2.1 |
| 31 - 50 | 0.2 | 0.8 | 0.0 | -0.1 - 0.6 |
| 51 - 64 | 0.2 | 1.0 | 0.0 | -0.1 - 0.4 |
| ≥65 | 0.5 | 1.9 | 0.0 | -0.2 - 0.7 |
| **Overall** | **0.3** | **1.4** | **0.0** | **-0.1 - 0.5** |

[1] ER visits are for the Winnipeg Regional Health Authority only; [2] The periods for which encounters were averaged (as number per year) was 180 days before to 730 days after diagnosis for invasive cancer; [3] Standard deviation; [4] Q1 and Q3 are the first and third quartile respectively

Supplementary Table 84 **Distribution of excess number of prescriptions[1] (per year) due to invasive cancer of the oral cavity (for patients compared to their matches) among Manitoba males by age group (1997-2015).**

| Age (years) | Average | SD[2] | Median | Q1 - Q3[3] |
|---|---|---|---|---|
| 18 - 30 | 11.3 | 14.0 | 11.3 | 1.5 - 21.2 |
| 31 - 50 | 11.7 | 35.5 | 1.7 | -3.4 - 21.9 |
| 51 - 64 | 10.1 | 44.5 | 9.7 | -2.3 - 24.0 |
| ≥65 | 13.6 | 90.8 | 7.4 | -13.3 - 28.4 |
| **Overall** | **11.9** | **67.3** | **6.8** | **-7.9 - 25.0** |

[1] The periods for which encounters were averaged (as number per year) was 180 days before to 730 days after diagnosis for invasive cancer; [2] Standard deviation; [3] Q1 and Q3 are the first and third quartile respectively

Supplementary Table 85 **Distribution of excess number of clinician visits[1] (per year) due to invasive cancer of the oropharynx (for patients compared to their matches) among Manitoba males by age group (1997-2015).**

| Age (years) | Average | SD[2] | Median | Q1 - Q3[3] |
|---|---|---|---|---|
| 31 - 50 | 11.8 | 9.6 | 10.3 | 5.7 - 17.9 |
| 51 - 64 | 11.4 | 11.8 | 9.5 | 4.5 - 16.8 |
| ≥65 | 7.5 | 17.9 | 7.6 | 2.3 - 16.9 |
| **Overall** | **10.2** | **13.8** | **9.3** | **3.7 - 17.5** |

[1] The periods for which encounters were averaged (as number per year) was 180 days before to 730 days after diagnosis for invasive cancer; [2] Standard deviation; [3] Q1 and Q3 are the first and third quartile respectively

Supplementary Table 86 **Distribution of excess number of hospital admissions[1] (per year) due to invasive cancer of the oropharynx (for patients compared to their matches) among Manitoba males by age group (1997-2015).**

| Age (years) | Average | SD[2] | Median | Q1 - Q3[3] |
|---|---|---|---|---|
| 31 - 50 | 1.1 | 1.0 | 0.8 | 0.4 - 1.3 |
| 51 - 64 | 0.9 | 1.1 | 0.7 | 0.3 - 1.3 |
| ≥65 | 0.9 | 1.4 | 0.5 | 0.0 - 1.6 |
| **Overall** | **0.9** | **1.2** | **0.7** | **0.3 - 1.4** |

[1] The periods for which encounters were averaged (as number per year) was 180 days before to 730 days after diagnosis for invasive cancer; [2] Standard deviation; [3] Q1 and Q3 are the first and third quartile respectively

Supplementary Table 87 **Distribution of excess number of ER visits[1,2] (per year) due to invasive cancer of the oropharynx (for patients compared to their matches) among Manitoba males by age group (1997-2015).**

| Age (years) | Average | SD[3] | Median | Q1 - Q3[4] |
|---|---|---|---|---|
| 31 - 50 | 0.2 | 0.8 | 0.0 | -0.1 - 0.4 |
| 51 - 64 | 0.4 | 1.2 | 0.1 | 0.0 - 0.4 |
| ≥65 | 0.3 | 1.5 | 0.0 | -0.2 - 0.8 |
| **Overall** | **0.4** | **1.2** | **0.0** | **-0.1 - 0.4** |

[1] ER visits are for the Winnipeg Regional Health Authority only; [2] The periods for which encounters were averaged (as number per year) was 180 days before to 730 days after diagnosis for invasive cancer; [3] Standard deviation; [4] Q1 and Q3 are the first and third quartile respectively

Supplementary Table 88 **Distribution of excess number of prescriptions[1] (per year) due to invasive cancer of the oropharynx (for patients compared to their matches) among Manitoba males by age group (1997-2015).**

| Age (years) | Average | SD[2] | Median | Q1 - Q3[3] |
|---|---|---|---|---|
| 31 - 50 | 15.0 | 25.6 | 7.3 | 0.9 - 22.5 |
| 51 - 64 | 8.6 | 40.9 | 4.9 | -5.9 - 20.7 |
| ≥65 | -9.9 | 87.6 | 1.6 | -22.1 - 21.2 |
| **Overall** | **4.1** | **58.5** | **4.9** | **-7.1 - 20.9** |

[1] The periods for which encounters were averaged (as number per year) was 180 days before to 730 days after diagnosis for invasive cancer; [2] Standard deviation; [3] Q1 and Q3 are the first and third quartile respectively

Supplementary Table 89 **Distribution of excess number of clinician visits[1] (per year) due to invasive cancer of the penis (for patients compared to their matches) among Manitoba males by age group (1997-2015).**

| Age (years) | Average | SD[2] | Median | Q1 - Q3[3] |
|---|---|---|---|---|
| 18 - 30 | 10.8 | 6.7 | 6.9 | 6.9 - 18.5 |
| 31 - 50 | 8.3 | 10.9 | 5.7 | 2.4 - 16.3 |
| 51 - 64 | 5.2 | 14.5 | 6.4 | 0.7 - 11.5 |
| ≥65 | 7.0 | 15.7 | 6.5 | -0.3 - 13.2 |
| **Overall** | **6.8** | **14.7** | **6.5** | **0.7 - 12.9** |

[1] The periods for which encounters were averaged (as number per year) was 180 days before to 730 days after diagnosis for invasive cancer; [2] Standard deviation; [3] Q1 and Q3 are the first and third quartile respectively

Supplementary Table 90 **Distribution of excess number of hospital admissions[1] (per year) due to invasive cancer of the penis (for patients compared to their matches) among Manitoba males by age group (1997-2015).**

| Age (years) | Average | SD[2] | Median | Q1 - Q3[3] |
|---|---|---|---|---|
| 18 - 30 | 1.2 | 0.4 | 1.1 | 0.8 - 1.6 |
| 31 - 50 | 1.1 | 1.1 | 0.7 | 0.4 - 1.3 |
| 51 - 64 | 1.0 | 1.0 | 0.7 | 0.4 - 1.2 |
| ≥65 | 1.1 | 1.0 | 0.8 | 0.4 - 1.6 |
| **Overall** | **1.0** | **1.0** | **0.8** | **0.4 - 1.6** |

[1] The periods for which encounters were averaged (as number per year) was 180 days before to 730 days after diagnosis for invasive cancer; [2] Standard deviation; [3] Q1 and Q3 are the first and third quartile respectively

Supplementary Table 91 **Distribution of excess number of ER visits[1,2] (per year) due to invasive cancer of the penis (for patients compared to their matches) among Manitoba males by age group (1997-2015).**

| Age (years) | Average | SD[3] | Median | Q1 - Q3[4] |
|---|---|---|---|---|
| 18 - 30 | 0.3 | | 0.3 | 0.3 - 0.3 |
| 31 - 50 | 0.1 | 0.2 | 0.0 | 0.0 - 0.3 |
| 51 - 64 | 0.1 | 0.8 | 0.3 | -0.1 - 0.5 |
| ≥65 | 0.2 | 0.9 | 0.0 | -0.4 - 0.5 |
| **Overall** | **0.2** | **0.8** | **0.0** | **-0.1 - 0.4** |

[1] ER visits are for the Winnipeg Regional Health Authority only; [2] The periods for which encounters were averaged (as number per year) was 180 days before to 730 days after diagnosis for invasive cancer; [3] Standard deviation; [4] Q1 and Q3 are the first and third quartile respectively

Supplementary Table 92 **Distribution of excess number of prescriptions[1] (per year) due to invasive cancer of the penis (for patients compared to their matches) among Manitoba males by age group (1997-2015).**

| Age (years) | Average | SD[2] | Median | Q1 - Q3[3] |
|---|---|---|---|---|
| 18 - 30 | 14.7 | 12.2 | 16.9 | 1.6 - 25.6 |
| 31 - 50 | 8.4 | 21.2 | 3.9 | -2.8 - 17.6 |
| 51 - 64 | 10.6 | 41.3 | -2.7 | -11.2 - 28.0 |
| ≥65 | 4.1 | 57.8 | 1.3 | -26.1 - 31.9 |
| **Overall** | **6.3** | **50.6** | **1.2** | **-15.1 - 28.0** |

[1] The periods for which encounters were averaged (as number per year) was 180 days before to 730 days after diagnosis for invasive cancer; [2] Standard deviation; [3] Q1 and Q3 are the first and third quartile respectively

#### Excess healthcare utilization by regional health authority and Winnipeg region of residence

Supplementary Table 93 **Distribution of excess number of clinician visits[1] (per year) due to anogenital warts (for patients compared to their matches) among Manitoba males by regional health authority and Winnipeg region of residence (1997-2015).**

| Region | Average | SD[2] | Median | Q1 - Q3[3] |
|---|---|---|---|---|
| Winnipeg | 4.7 | 10.2 | 3.0 | -0.3 - 8.0 |
| Northern suburbs | 4.6 | 10.7 | 3.0 | -0.3 - 7.7 |
| Inner city | 5.2 | 11.0 | 3.0 | -0.3 - 9.0 |
| Southern suburbs | 4.6 | 9.6 | 3.0 | -0.3 - 8.0 |
| Interlake-Eastern | 4.2 | 9.9 | 2.7 | 0.0 - 7.0 |
| Northern | 4.0 | 9.1 | 2.3 | 0.0 - 6.0 |
| Southern | 4.0 | 7.0 | 3.0 | 0.0 - 7.0 |
| Prairie Mountain | 4.4 | 7.7 | 3.0 | 0.0 - 7.3 |
| Public Trustee/In CFS care | 5.4 | 12.6 | 3.2 | -1.7 - 8.0 |
| **Overall** | **4.6** | **9.7** | **3.0** | **-0.3 - 7.7** |

[1] The period for which encounters were averaged (as number per year) was 0 to 365 days after diagnosis for AGW; [2] Standard deviation; [3] Q1 and Q3 are the first and third quartile respectively

Supplementary Table 94 **Distribution of excess number of hospital admissions[1] (per year) due to anogenital warts (for patients compared to their matches) among Manitoba males by regional health authority and Winnipeg region of residence (1997-2015).**

| Region | Average | SD[2] | Median | Q1 - Q3[3] |
|---|---|---|---|---|
| Winnipeg | 0.1 | 0.7 | 0.0 | 0.0 - 0.0 |
| Northern suburbs | 0.2 | 0.6 | 0.0 | 0.0 - 0.0 |
| Inner city | 0.2 | 1.0 | 0.0 | 0.0 - 0.0 |
| Southern suburbs | 0.1 | 0.5 | 0.0 | 0.0 - 0.0 |
| Interlake-Eastern | 0.2 | 0.8 | 0.0 | 0.0 - 0.0 |
| Northern | 0.4 | 1.0 | 0.0 | 0.0 - 1.0 |
| Southern | 0.1 | 0.6 | 0.0 | 0.0 - 0.0 |
| Prairie Mountain | 0.1 | 0.6 | 0.0 | 0.0 - 0.0 |
| Public Trustee/In CFS care | 0.5 | 1.0 | 0.0 | 0.0 - 1.0 |
| **Overall** | **0.2** | **0.7** | **0.0** | **0.0 - 0.0** |

[1] The period for which encounters were averaged (as number per year) was 0 to 365 days after diagnosis for AGW; [2] Standard deviation; [3] Q1 and Q3 are the first and third quartile respectively

Supplementary Table 95 **Distribution of excess number of ER visits[1,2] (per year) due to anogenital warts (for patients compared to their matches) among Manitoba males by regional health authority and Winnipeg region of residence (1997-2015).**

| Region | Average | SD[3] | Median | Q1 - Q3[4] |
|---|---|---|---|---|
| Winnipeg | 0.1 | 1.2 | 0.0 | -0.3 - 0.0 |
| Northern suburbs | 0.2 | 1.0 | 0.0 | -0.3 - 0.0 |
| Inner city | 0.1 | 2.2 | 0.0 | -0.3 - 0.0 |
| Southern suburbs | 0.1 | 0.8 | 0.0 | 0.0 - 0.0 |
| **Overall** | **0.1** | **1.2** | **0.0** | **-0.3 - 0.0** |

[1] ER visits are for the Winnipeg Regional Health Authority only; [2] The period for which encounters were averaged (as number per year) was 0 to 365 days after diagnosis for AGW; [3] Standard deviation; [4] Q1 and Q3 are the first and third quartile respectively

Supplementary Table 96 **Distribution of excess number of prescriptions[1] (per year) due to anogenital warts (for patients compared to their matches) among Manitoba males by regional health authority and Winnipeg region of residence (1997-2015).**

| Region | Average | SD[2] | Median | Q1 - Q3[3] |
|---|---|---|---|---|
| Winnipeg | 2.3 | 27.0 | 0.0 | -2.3 - 4.0 |
| Northern suburbs | 2.9 | 28.0 | 0.0 | -2.0 - 4.0 |
| Inner city | 3.6 | 35.9 | 0.0 | -3.7 - 5.0 |
| Southern suburbs | 1.6 | 22.2 | 0.0 | -2.3 - 4.0 |
| Interlake-Eastern | 2.7 | 19.9 | 0.3 | -2.3 - 4.0 |
| Northern | 4.4 | 17.4 | 0.7 | -1.7 - 5.7 |
| Southern | 2.3 | 14.6 | 0.3 | -1.7 - 4.0 |
| Prairie Mountain | 2.4 | 30.1 | 0.0 | -2.3 - 4.0 |
| Public Trustee/In CFS care | 3.1 | 48.2 | -0.6 | -9.0 - 6.7 |
| **Overall** | **2.4** | **25.9** | **0.0** | **-2.3 - 4.0** |

[1] The period for which encounters were averaged (as number per year) was 0 to 365 days after diagnosis for AGW; [2] Standard deviation; [3] Q1 and Q3 are the first and third quartile respectively

Supplementary Table 97 **Distribution of excess number of clinician visits[1] (per year) due to HPV-related carcinoma *in situ* (for patients compared to their matches) among Manitoba males by regional health authority and Winnipeg region of residence (1997-2015).**

| Region | Average | SD[2] | Median | Q1 - Q3[3] |
|---|---|---|---|---|
| Winnipeg | 9.0 | 11.6 | 5.7 | 1.0 - 16.0 |
| Northern suburbs | 11.2 | 12.5 | 11.0 | 1.7 - 17.7 |
| Inner city | 13.6 | 15.5 | 9.3 | 2.5 - 25.0 |
| Southern suburbs | 5.9 | 8.4 | 4.0 | 0.7 - 12.0 |
| Interlake-Eastern | 8.7 | 8.7 | 8.0 | 2.8 - 16.7 |
| Northern | 8.8 | 11.2 | 6.3 | 2.0 - 9.7 |
| Southern | -0.1 | 6.0 | 3.0 | -7.0 - 3.7 |
| Prairie Mountain | 5.4 | 10.9 | 3.5 | -1.8 - 12.7 |
| Public Trustee/In CFS care | 21.0 | | 21.0 | 21.0 - 21.0 |
| **Overall** | **8.6** | **11.1** | **4.8** | **1.2 - 15.8** |

[1] The period for which encounters were averaged (as number per year) was 0 to 365 days after diagnosis for carcinoma *in situ*; [2] Standard deviation; [3] Q1 and Q3 are the first and third quartile respectively

Supplementary Table 98 **Distribution of excess number of hospital admissions[1] (per year) due to HPV-related carcinoma *in situ* (for patients compared to their matches) among Manitoba males by regional health authority and Winnipeg region of residence (1997-2015).**

| Region | Average | SD[2] | Median | Q1 - Q3[3] |
|---|---|---|---|---|
| Winnipeg | 0.8 | 1.0 | 1.0 | 0.0 - 1.3 |
| Northern suburbs | 0.9 | 0.7 | 1.0 | 0.0 - 1.3 |
| Inner city | 0.5 | 1.0 | 0.5 | -0.2 - 1.0 |
| Southern suburbs | 0.9 | 1.1 | 1.0 | 0.0 - 1.7 |
| Interlake-Eastern | 0.7 | 1.3 | 0.3 | -0.3 - 1.7 |
| Northern | 0.8 | 0.9 | 0.7 | 0.0 - 1.7 |
| Southern | 0.3 | 1.2 | 1.0 | -1.0 - 1.0 |
| Prairie Mountain | 0.5 | 1.9 | 1.0 | -1.0 - 2.0 |
| Public Trustee/In CFS care | 0.0 | | 0.0 | 0.0 - 0.0 |
| **Overall** | **0.8** | **1.0** | **0.7** | **0.0 - 1.3** |

[1] The period for which encounters were averaged (as number per year) was 0 to 365 days after diagnosis for carcinoma *in situ*; [2] Standard deviation; [3] Q1 and Q3 are the first and third quartile respectively

Supplementary Table 99 **Distribution of excess number of ER visits[1,2] (per year) due to HPV-related carcinoma *in situ* (for patients compared to their matches) among Manitoba males by regional health authority and Winnipeg region of residence (1997-2015).**

| Region | Average | SD[3] | Median | Q1 - Q3[4] |
|---|---|---|---|---|
| Winnipeg | -0.0 | 1.3 | 0.0 | -0.3 - 0.0 |
| Northern suburbs | 0.3 | 1.4 | 0.0 | -0.3 - 0.0 |
| Inner city | -0.4 | 2.1 | 0.0 | -1.3 - 0.5 |
| Southern suburbs | -0.1 | 0.8 | 0.0 | -0.3 - 0.0 |
| **Overall** | **-0.0** | **1.3** | **0.0** | **-0.3 - 0.0** |

[1] ER visits are for the Winnipeg Regional Health Authority only; [2] The period for which encounters were averaged (as number per year) was 0 to 365 days after diagnosis for carcinoma *in situ*; [3] Standard deviation; [4] Q1 and Q3 are the first and third quartile respectively

Supplementary Table 100 **Distribution of excess number of prescriptions[1] (per year) due to HPV-related carcinoma *in situ* (for patients compared to their matches) among Manitoba males by regional health authority and Winnipeg region of residence (1997-2015).**

| Region | Average | SD[2] | Median | Q1 - Q3[3] |
|---|---|---|---|---|
| Winnipeg | 10.5 | 52.4 | 1.2 | -10.7 - 16.7 |
| Northern suburbs | 5.0 | 19.9 | 0.7 | -1.7 - 10.7 |
| Inner city | 37.5 | 103.0 | 44.5 | -23.8 - 130.0 |
| Southern suburbs | 2.6 | 28.2 | 1.0 | -10.7 - 6.7 |
| Interlake-Eastern | 12.6 | 66.7 | 3.2 | -23.5 - 22.7 |
| Northern | 9.1 | 29.9 | 1.0 | -5.7 - 7.3 |
| Southern | -7.1 | 17.6 | -13.0 | -21.0 - 12.7 |
| Prairie Mountain | 6.0 | 11.2 | 5.0 | -1.2 - 13.2 |
| Public Trustee/In CFS care | -0.7 | | -0.7 | -0.7 - -0.7 |
| **Overall** | **9.6** | **49.6** | **1.8** | **-10.0 - 14.2** |

[1] The period for which encounters were averaged (as number per year) was 0 to 365 days after diagnosis for carcinoma *in situ*; [2] Standard deviation; [3] Q1 and Q3 are the first and third quartile respectively

Supplementary Table 101 **Distribution of excess number of clinician visits[1] (per year) due to HPV-related invasive cancer (for patients compared to their matches) among Manitoba males by regional health authority and Winnipeg region of residence (1997-2015).**

| Region | Average | SD[2] | Median | Q1 - Q3[3] |
|---|---|---|---|---|
| Winnipeg | 9.9 | 16.4 | 8.7 | 2.5 - 16.9 |
| Northern suburbs | 8.5 | 11.7 | 7.7 | 2.7 - 14.5 |
| Inner city | 12.4 | 24.1 | 9.5 | 3.1 - 21.0 |
| Southern suburbs | 9.6 | 14.5 | 8.5 | 2.2 - 16.8 |
| Interlake-Eastern | 9.9 | 10.4 | 8.5 | 2.7 - 15.6 |
| Northern | 10.5 | 16.9 | 7.6 | 0.1 - 14.6 |
| Southern | 11.9 | 14.8 | 10.4 | 6.0 - 18.2 |
| Prairie Mountain | 10.5 | 14.6 | 8.4 | 3.3 - 16.3 |
| Public Trustee/In CFS care | 8.9 | 15.7 | 9.0 | 6.4 - 11.5 |
| **Overall** | **10.2** | **15.4** | **8.8** | **3.0 - 16.7** |

[1] The periods for which encounters were averaged (as number per year) was 180 days before to 730 days after diagnosis for invasive cancer; [2] Standard deviation; [3] Q1 and Q3 are the first and third quartile respectively

Supplementary Table 102 **Distribution of excess number of hospital admissions[1] (per year) due to HPV-related invasive cancer (for patients compared to their matches) among Manitoba males by regional health authority and Winnipeg region of residence (1997-2015).**

| Region | Average | SD[2] | Median | Q1 - Q3[3] |
|---|---|---|---|---|
| Winnipeg | 1.0 | 1.3 | 0.8 | 0.3 - 1.6 |
| Northern suburbs | 1.0 | 1.5 | 0.7 | 0.3 - 1.5 |
| Inner city | 1.3 | 1.4 | 0.9 | 0.5 - 1.9 |
| Southern suburbs | 0.9 | 1.0 | 0.7 | 0.3 - 1.3 |
| Interlake-Eastern | 1.0 | 1.1 | 0.7 | 0.3 - 1.6 |
| Northern | 1.2 | 1.2 | 0.8 | 0.4 - 1.6 |
| Southern | 1.1 | 1.1 | 0.8 | 0.4 - 1.6 |
| Prairie Mountain | 1.1 | 1.0 | 0.8 | 0.4 - 1.6 |
| Public Trustee/In CFS care | 0.2 | 0.6 | 0.3 | -0.4 - 0.6 |
| **Overall** | **1.0** | **1.2** | **0.8** | **0.4 - 1.6** |

[1] The periods for which encounters were averaged (as number per year) was 180 days before to 730 days after diagnosis for invasive cancer; [2] Standard deviation; [3] Q1 and Q3 are the first and third quartile respectively

Supplementary Table 103 **Distribution of excess number of ER visits[1,2] (per year) due to HPV-related invasive cancer (for patients compared to their matches) among Manitoba males by regional health authority and Winnipeg region of residence (1997-2015).**

| Region | Average | SD[3] | Median | Q1 - Q3[4] |
|---|---|---|---|---|
| Winnipeg | 0.5 | 2.1 | 0.0 | -0.1 - 0.5 |
| Northern suburbs | 0.3 | 1.1 | 0.0 | -0.1 - 0.5 |
| Inner city | 1.0 | 4.2 | 0.0 | -0.1 - 0.8 |
| Southern suburbs | 0.3 | 1.1 | 0.0 | -0.1 - 0.6 |
| **Overall** | **0.5** | **2.1** | **0.0** | **-0.1 - 0.5** |

[1] ER visits are for the Winnipeg Regional Health Authority only; [2] The periods for which encounters were averaged (as number per year) was 180 days before to 730 days after diagnosis for invasive cancer; [3] Standard deviation; [4] Q1 and Q3 are the first and third quartile respectively

Supplementary Table 104 **Distribution of excess number of prescriptions[1] (per year) due to HPV-related invasive cancer (for patients compared to their matches) among Manitoba males by regional health authority and Winnipeg region of residence (1997-2015).**

| Region | Average | SD[2] | Median | Q1 - Q3[3] |
|---|---|---|---|---|
| Winnipeg | 11.2 | 88.9 | 5.1 | -7.9 - 22.3 |
| Northern suburbs | -3.5 | 74.2 | 4.1 | -9.7 - 18.2 |
| Inner city | 36.5 | 161.7 | 9.1 | -11.4 - 42.7 |
| Southern suburbs | 8.8 | 40.3 | 4.3 | -5.3 - 19.5 |
| Interlake-Eastern | 8.5 | 33.9 | 3.9 | -8.9 - 21.9 |
| Northern | 5.4 | 38.4 | 1.6 | -9.3 - 25.4 |
| Southern | 14.6 | 40.2 | 10.9 | -3.3 - 24.3 |
| Prairie Mountain | 18.0 | 148.7 | 6.7 | -15.2 - 29.6 |
| Public Trustee/In CFS care | 48.3 | 66.2 | 52.5 | 17.7 - 69.2 |
| **Overall** | **12.2** | **91.5** | **6.0** | **-7.9 - 24.6** |

[1] The periods for which encounters were averaged (as number per year) was 180 days before to 730 days after diagnosis for invasive cancer; [2] Standard deviation; [3] Q1 and Q3 are the first and third quartile respectively

Supplementary Table 105 **Distribution of excess number of clinician visits[1] (per year) due to invasive cancer of the anus (for patients compared to their matches) among Manitoba males by regional health authority and Winnipeg region of residence (1997-2015).**

| Region | Average | SD[2] | Median | Q1 - Q3[3] |
|---|---|---|---|---|
| Winnipeg | 14.7 | 22.6 | 10.9 | 5.5 - 19.1 |
| Northern suburbs | 13.3 | 14.6 | 10.9 | 10.3 - 16.7 |
| Inner city | 26.5 | 24.7 | 17.5 | 9.3 - 32.5 |
| Southern suburbs | 7.9 | 22.2 | 9.7 | 4.8 - 17.2 |
| Interlake-Eastern | 23.0 | 15.8 | 17.7 | 8.5 - 31.2 |
| Northern | 8.5 | 12.3 | 8.5 | -0.2 - 17.2 |
| Southern | 21.8 | 5.9 | 21.7 | 16.0 - 27.7 |
| Prairie Mountain | 21.0 | 14.7 | 15.7 | 12.7 - 28.4 |
| **Overall** | **16.2** | **20.6** | **14.4** | **6.9 - 23.6** |

[1] The periods for which encounters were averaged (as number per year) was 180 days before to 730 days after diagnosis for invasive cancer; [2] Standard deviation; [3] Q1 and Q3 are the first and third quartile respectively

Supplementary Table 106 **Distribution of excess number of hospital admissions[1] (per year) due to invasive cancer of the anus (for patients compared to their matches) among Manitoba males by regional health authority and Winnipeg region of residence (1997-2015).**

| Region | Average | SD[2] | Median | Q1 - Q3[3] |
|---|---|---|---|---|
| Winnipeg | 1.8 | 1.7 | 1.3 | 0.7 - 2.4 |
| Northern suburbs | 1.3 | 1.0 | 0.9 | 0.4 - 2.1 |
| Inner city | 2.8 | 2.4 | 2.0 | 1.1 - 3.2 |
| Southern suburbs | 1.4 | 1.0 | 1.3 | 0.7 - 2.4 |
| Interlake-Eastern | 1.6 | 1.3 | 1.6 | 0.7 - 2.4 |
| Northern | 1.0 | 0.0 | 1.0 | 1.0 - 1.1 |
| Southern | 2.5 | 1.8 | 2.9 | 0.5 - 4.1 |
| Prairie Mountain | 1.6 | 1.1 | 2.0 | 0.8 - 2.4 |
| **Overall** | **1.8** | **1.6** | **1.4** | **0.7 - 2.4** |

[1] The periods for which encounters were averaged (as number per year) was 180 days before to 730 days after diagnosis for invasive cancer; [2] Standard deviation; [3] Q1 and Q3 are the first and third quartile respectively

Supplementary Table 107 **Distribution of excess number of ER visits[1,2] (per year) due to invasive cancer of the anus (for patients compared to their matches) among Manitoba males by regional health authority and Winnipeg region of residence (1997-2015).**

| Region | Average | SD[3] | Median | Q1 - Q3[4] |
|---|---|---|---|---|
| Winnipeg | 2.2 | 6.2 | 0.4 | 0.0 - 1.5 |
| Northern suburbs | 0.5 | 0.6 | 0.4 | 0.0 - 1.2 |
| Inner city | 5.8 | 10.6 | 1.2 | 0.1 - 9.1 |
| Southern suburbs | 0.7 | 1.0 | 0.3 | 0.0 - 1.6 |
| **Overall** | **2.2** | **6.2** | **0.4** | **0.0 - 1.5** |

[1] ER visits are for the Winnipeg Regional Health Authority only; [2] The periods for which encounters were averaged (as number per year) was 180 days before to 730 days after diagnosis for invasive cancer; [3] Standard deviation; [4] Q1 and Q3 are the first and third quartile respectively

Supplementary Table 108 **Distribution of excess number of prescriptions[1] (per year) due to invasive cancer of the anus (for patients compared to their matches) among Manitoba males by regional health authority and Winnipeg region of residence (1997-2015).**

| Region | Average | SD[2] | Median | Q1 - Q3[3] |
|---|---|---|---|---|
| Winnipeg | 74.0 | 258.2 | 8.8 | -1.6 - 65.5 |
| Northern suburbs | 25.4 | 112.3 | 6.0 | -13.7 - 9.6 |
| Inner city | 173.3 | 451.3 | 39.0 | 9.2 - 115.6 |
| Southern suburbs | 35.0 | 67.2 | 5.1 | -2.4 - 37.9 |
| Interlake-Eastern | 32.1 | 67.2 | 7.5 | -10.9 - 47.6 |
| Northern | 11.4 | 21.8 | 11.4 | -4.0 - 26.8 |
| Southern | 34.2 | 23.1 | 33.2 | 11.6 - 57.7 |
| Prairie Mountain | 256.4 | 545.5 | 21.6 | 7.1 - 27.2 |
| **Overall** | **79.8** | **266.5** | **9.2** | **-1.5 - 46.8** |

[1] The periods for which encounters were averaged (as number per year) was 180 days before to 730 days after diagnosis for invasive cancer; [2] Standard deviation; [3] Q1 and Q3 are the first and third quartile respectively

Supplementary Table 109 **Distribution of excess number of clinician visits[1] (per year) due to invasive cancer of the oral cavity (for patients compared to their matches) among Manitoba males by regional health authority and Winnipeg region of residence (1997-2015).**

| Region | Average | SD[2] | Median | Q1 - Q3[3] |
|---|---|---|---|---|
| Winnipeg | 10.2 | 17.7 | 7.9 | 1.7 - 16.3 |
| Northern suburbs | 7.9 | 13.0 | 6.1 | 0.7 - 13.3 |
| Inner city | 10.5 | 29.1 | 6.1 | 0.9 - 15.5 |
| Southern suburbs | 11.2 | 13.5 | 8.9 | 2.2 - 18.4 |
| Interlake-Eastern | 7.0 | 9.3 | 6.8 | 0.1 - 12.5 |
| Northern | 7.0 | 13.4 | 5.7 | 0.1 - 10.8 |
| Southern | 12.0 | 9.0 | 10.5 | 6.0 - 17.3 |
| Prairie Mountain | 12.4 | 17.6 | 10.2 | 2.5 - 16.3 |
| Public Trustee/In CFS care | 3.8 | 12.5 | 9.0 | 6.4 - 10.6 |
| **Overall** | **10.1** | **16.2** | **8.1** | **2.1 - 15.5** |

[1] The periods for which encounters were averaged (as number per year) was 180 days before to 730 days after diagnosis for invasive cancer; [2] Standard deviation; [3] Q1 and Q3 are the first and third quartile respectively

Supplementary Table 110 **Distribution of excess number of hospital admissions[1] (per year) due to invasive cancer of the oral cavity (for patients compared to their matches) among Manitoba males by regional health authority and Winnipeg region of residence (1997-2015).**

| Region | Average | SD[2] | Median | Q1 - Q3[3] |
|---|---|---|---|---|
| Winnipeg | 1.0 | 1.2 | 0.8 | 0.3 - 1.4 |
| Northern suburbs | 1.0 | 1.8 | 0.5 | 0.3 - 1.3 |
| Inner city | 1.1 | 0.8 | 0.9 | 0.7 - 1.6 |
| Southern suburbs | 0.9 | 1.1 | 0.7 | 0.3 - 1.4 |
| Interlake-Eastern | 1.0 | 0.9 | 0.8 | 0.4 - 1.6 |
| Northern | 1.1 | 0.8 | 1.2 | 0.4 - 1.7 |
| Southern | 1.1 | 0.8 | 1.0 | 0.5 - 1.9 |
| Prairie Mountain | 1.1 | 1.0 | 0.8 | 0.4 - 2.0 |
| Public Trustee/In CFS care | 0.2 | 0.7 | 0.3 | -0.4 - 0.6 |
| **Overall** | **1.0** | **1.1** | **0.8** | **0.4 - 1.6** |

[1] The periods for which encounters were averaged (as number per year) was 180 days before to 730 days after diagnosis for invasive cancer; [2] Standard deviation; [3] Q1 and Q3 are the first and third quartile respectively

Supplementary Table 111 **Distribution of excess number of ER visits[1,2] (per year) due to invasive cancer of the oral cavity (for patients compared to their matches) among Manitoba males by regional health authority and Winnipeg region of residence (1997-2015).**

| Region | Average | SD[3] | Median | Q1 - Q3[4] |
|---|---|---|---|---|
| Winnipeg | 0.3 | 1.4 | 0.0 | -0.1 - 0.5 |
| Northern suburbs | 0.2 | 0.9 | 0.0 | -0.1 - 0.4 |
| Inner city | 0.6 | 2.4 | 0.0 | -0.1 - 0.8 |
| Southern suburbs | 0.3 | 1.1 | 0.0 | -0.1 - 0.7 |
| **Overall** | **0.3** | **1.4** | **0.0** | **-0.1 - 0.5** |

[1] ER visits are for the Winnipeg Regional Health Authority only; [2] The periods for which encounters were averaged (as number per year) was 180 days before to 730 days after diagnosis for invasive cancer; [3] Standard deviation; [4] Q1 and Q3 are the first and third quartile respectively

Supplementary Table 112 **Distribution of excess number of prescriptions[1] (per year) due to invasive cancer of the oral cavity (for patients compared to their matches) among Manitoba males by regional health authority and Winnipeg region of residence (1997-2015).**

| Region | Average | SD[2] | Median | Q1 - Q3[3] |
|---|---|---|---|---|
| Winnipeg | 8.7 | 60.0 | 9.6 | -8.0 - 25.6 |
| Northern suburbs | -7.4 | 79.8 | 5.9 | -10.4 - 20.0 |
| Inner city | 21.7 | 62.2 | 11.5 | -26.4 - 64.9 |
| Southern suburbs | 11.5 | 45.1 | 10.1 | -2.9 - 24.6 |
| Interlake-Eastern | 1.3 | 29.0 | -0.5 | -14.3 - 6.8 |
| Northern | -3.0 | 40.8 | 0.5 | -8.9 - 12.2 |
| Southern | 10.5 | 23.8 | 9.9 | -4.0 - 23.2 |
| Prairie Mountain | 41.0 | 127.6 | 7.2 | -2.5 - 40.3 |
| Public Trustee/In CFS care | 42.1 | 80.0 | 45.1 | 17.7 - 52.5 |
| **Overall** | **11.9** | **67.3** | **6.8** | **-7.9 - 25.0** |

[1] The periods for which encounters were averaged (as number per year) was 180 days before to 730 days after diagnosis for invasive cancer; [2] Standard deviation; [3] Q1 and Q3 are the first and third quartile respectively

Supplementary Table 113 **Distribution of excess number of clinician visits[1] (per year) due to invasive cancer of the oropharynx (for patients compared to their matches) among Manitoba males by regional health authority and Winnipeg region of residence (1997-2015).**

| Region | Average | SD[2] | Median | Q1 - Q3[3] |
|---|---|---|---|---|
| Winnipeg | 9.8 | 13.9 | 10.0 | 3.5 - 17.6 |
| Northern suburbs | 9.0 | 10.5 | 9.6 | 4.7 - 16.0 |
| Inner city | 12.7 | 19.9 | 16.1 | 5.3 - 26.8 |
| Southern suburbs | 9.2 | 13.1 | 8.5 | 2.5 - 15.7 |
| Interlake-Eastern | 10.4 | 8.8 | 9.3 | 3.1 - 16.6 |
| Northern | 14.6 | 20.1 | 10.0 | 5.2 - 15.7 |
| Southern | 9.7 | 19.6 | 9.0 | 5.4 - 15.9 |
| Prairie Mountain | 11.5 | 9.7 | 9.2 | 5.7 - 17.7 |
| **Overall** | **10.2** | **13.8** | **9.3** | **3.7 - 17.5** |

[1] The periods for which encounters were averaged (as number per year) was 180 days before to 730 days after diagnosis for invasive cancer; [2] Standard deviation; [3] Q1 and Q3 are the first and third quartile respectively

Supplementary Table 114 **Distribution of excess number of hospital admissions[1] (per year) due to invasive cancer of the oropharynx (for patients compared to their matches) among Manitoba males by regional health authority and Winnipeg region of residence (1997-2015).**

| Region | Average | SD[2] | Median | Q1 - Q3[3] |
|---|---|---|---|---|
| Winnipeg | 0.9 | 1.2 | 0.7 | 0.3 - 1.3 |
| Northern suburbs | 1.1 | 1.4 | 0.5 | 0.3 - 1.5 |
| Inner city | 1.0 | 1.3 | 0.9 | 0.4 - 1.9 |
| Southern suburbs | 0.8 | 1.0 | 0.7 | 0.3 - 1.2 |
| Interlake-Eastern | 0.8 | 1.3 | 0.7 | 0.1 - 1.5 |
| Northern | 1.2 | 1.3 | 0.8 | 0.3 - 1.2 |
| Southern | 0.7 | 1.0 | 0.6 | 0.2 - 1.2 |
| Prairie Mountain | 1.2 | 1.0 | 1.0 | 0.4 - 1.6 |
| **Overall** | **0.9** | **1.2** | **0.7** | **0.3 - 1.4** |

[1] The periods for which encounters were averaged (as number per year) was 180 days before to 730 days after diagnosis for invasive cancer; [2] Standard deviation; [3] Q1 and Q3 are the first and third quartile respectively

Supplementary Table 115 **Distribution of excess number of ER visits[1,2] (per year) due to invasive cancer of the oropharynx (for patients compared to their matches) among Manitoba males by regional health authority and Winnipeg region of residence (1997-2015).**

| Region | Average | SD[3] | Median | Q1 - Q3[4] |
|---|---|---|---|---|
| Winnipeg | 0.4 | 1.2 | 0.0 | -0.1 - 0.4 |
| Northern suburbs | 0.4 | 1.4 | 0.0 | -0.1 - 0.5 |
| Inner city | 0.4 | 1.5 | 0.0 | -0.1 - 0.4 |
| Southern suburbs | 0.3 | 1.0 | 0.0 | -0.1 - 0.5 |
| **Overall** | **0.4** | **1.2** | **0.0** | **-0.1 - 0.4** |

[1] ER visits are for the Winnipeg Regional Health Authority only; [2] The periods for which encounters were averaged (as number per year) was 180 days before to 730 days after diagnosis for invasive cancer; [3] Standard deviation; [4] Q1 and Q3 are the first and third quartile respectively

Supplementary Table 116 **Distribution of excess number of prescriptions[1] (per year) due to invasive cancer of the oropharynx (for patients compared to their matches) among Manitoba males by regional health authority and Winnipeg region of residence (1997-2015).**

| Region | Average | SD[2] | Median | Q1 - Q3[3] |
|---|---|---|---|---|
| Winnipeg | 5.4 | 47.6 | 4.1 | -6.7 - 17.6 |
| Northern suburbs | -3.6 | 67.4 | 3.5 | -8.4 - 21.3 |
| Inner city | 23.2 | 57.5 | 8.3 | -4.0 - 31.5 |
| Southern suburbs | 4.2 | 23.8 | 2.8 | -6.5 - 13.1 |
| Interlake-Eastern | 11.5 | 32.0 | 12.5 | -6.7 - 25.9 |
| Northern | 15.7 | 38.8 | 10.1 | -9.3 - 34.9 |
| Southern | 9.7 | 40.5 | 5.3 | -3.7 - 19.6 |
| Prairie Mountain | -17.2 | 116.0 | 3.7 | -25.9 - 24.0 |
| **Overall** | **4.1** | **58.5** | **4.9** | **-7.1 - 20.9** |

[1] The periods for which encounters were averaged (as number per year) was 180 days before to 730 days after diagnosis for invasive cancer; [2] Standard deviation; [3] Q1 and Q3 are the first and third quartile respectively

Supplementary Table 117 **Distribution of excess number of clinician visits[1] (per year) due to invasive cancer of the penis (for patients compared to their matches) among Manitoba males by regional health authority and Winnipeg region of residence (1997-2015).**

| Region | Average | SD[2] | Median | Q1 - Q3[3] |
|---|---|---|---|---|
| Winnipeg | 4.9 | 14.4 | 5.6 | -2.4 - 12.0 |
| Northern suburbs | 5.0 | 9.4 | 5.6 | 2.3 - 10.1 |
| Inner city | 4.3 | 9.8 | 5.5 | -2.4 - 8.7 |
| Southern suburbs | 5.1 | 19.3 | 5.0 | -3.1 - 15.1 |
| Interlake-Eastern | 9.6 | 11.7 | 7.6 | 0.4 - 13.4 |
| Northern | 5.0 | 12.7 | 7.8 | -4.1 - 14.1 |
| Southern | 15.5 | 14.7 | 13.6 | 4.4 - 32.2 |
| Prairie Mountain | 5.4 | 15.4 | 6.5 | 1.3 - 12.9 |
| Public Trustee/In CFS care | 21.7 | 19.6 | 21.7 | 7.9 - 35.6 |
| **Overall** | **6.8** | **14.7** | **6.5** | **0.7 - 12.9** |

[1] The periods for which encounters were averaged (as number per year) was 180 days before to 730 days after diagnosis for invasive cancer; [2] Standard deviation; [3] Q1 and Q3 are the first and third quartile respectively

Supplementary Table 118 **Distribution of excess number of hospital admissions[1] (per year) due to invasive cancer of the penis (for patients compared to their matches) among Manitoba males by regional health authority and Winnipeg region of residence (1997-2015).**

| Region | Average | SD[2] | Median | Q1 - Q3[3] |
|---|---|---|---|---|
| Winnipeg | 1.1 | 0.9 | 0.8 | 0.4 - 1.6 |
| Northern suburbs | 1.1 | 0.9 | 0.9 | 0.4 - 1.6 |
| Inner city | 1.3 | 0.8 | 1.2 | 0.8 - 1.7 |
| Southern suburbs | 1.0 | 1.1 | 0.7 | 0.4 - 1.2 |
| Interlake-Eastern | 1.2 | 0.9 | 0.9 | 0.6 - 1.5 |
| Northern | 1.3 | 2.1 | 0.5 | 0.1 - 2.5 |
| Southern | 1.6 | 1.1 | 1.6 | 0.7 - 2.0 |
| Prairie Mountain | 0.8 | 0.9 | 0.7 | 0.3 - 1.5 |
| Public Trustee/In CFS care | 0.3 | 0.0 | 0.3 | 0.3 - 0.3 |
| **Overall** | **1.0** | **1.0** | **0.8** | **0.4 - 1.6** |

[1] The periods for which encounters were averaged (as number per year) was 180 days before to 730 days after diagnosis for invasive cancer; [2] Standard deviation; [3] Q1 and Q3 are the first and third quartile respectively

Supplementary Table 119 **Distribution of excess number of ER visits[1,2] (per year) due to invasive cancer of the penis (for patients compared to their matches) among Manitoba males by regional health authority and Winnipeg region of residence (1997-2015).**

| Region | Average | SD[3] | Median | Q1 - Q3[4] |
|---|---|---|---|---|
| Winnipeg | 0.2 | 0.8 | 0.0 | -0.1 - 0.4 |
| Northern suburbs | 0.3 | 0.7 | 0.0 | 0.0 - 0.3 |
| Inner city | 0.0 | 0.8 | 0.1 | -0.5 - 0.4 |
| Southern suburbs | 0.2 | 0.9 | 0.1 | -0.1 - 0.5 |
| **Overall** | **0.2** | **0.8** | **0.0** | **-0.1 - 0.4** |

[1] ER visits are for the Winnipeg Regional Health Authority only; [2] The periods for which encounters were averaged (as number per year) was 180 days before to 730 days after diagnosis for invasive cancer; [3] Standard deviation; [4] Q1 and Q3 are the first and third quartile respectively

Supplementary Table 120 **Distribution of excess number of prescriptions[1] (per year) due to invasive cancer of the penis (for patients compared to their matches) among Manitoba males by regional health authority and Winnipeg region of residence (1997-2015).**

| Region | Average | SD[2] | Median | Q1 - Q3[3] |
|---|---|---|---|---|
| Winnipeg | -3.2 | 49.3 | -2.6 | -14.1 - 16.1 |
| Northern suburbs | -9.1 | 50.4 | 1.6 | -15.1 - 8.9 |
| Inner city | -0.6 | 39.4 | -2.6 | -12.8 - 4.2 |
| Southern suburbs | -1.2 | 55.7 | -4.5 | -26.1 - 17.6 |
| Interlake-Eastern | 4.3 | 24.5 | 0.2 | -10.1 - 19.2 |
| Northern | -16.5 | 27.8 | -7.2 | -34.4 - 1.3 |
| Southern | 36.5 | 70.5 | 22.3 | -3.0 - 46.7 |
| Prairie Mountain | 10.1 | 50.7 | 6.3 | -28.5 - 47.5 |
| Public Trustee/In CFS care | 64.0 | 7.4 | 64.0 | 58.7 - 69.2 |
| **Overall** | **6.3** | **50.6** | **1.2** | **-15.1 - 28.0** |

[1] The periods for which encounters were averaged (as number per year) was 180 days before to 730 days after diagnosis for invasive cancer; [2] Standard deviation; [3] Q1 and Q3 are the first and third quartile respectively

#### Excess healthcare utilization by year of diagnosis

Supplementary Table 121 **Distribution of excess number of clinician visits[1] (per year) due to anogenital warts (for patients compared to their matches) among Manitoba males by year of diagnosis (1997-2015).**

| Year of diagnosis | Average | SD[2] | Median | Q1 - Q3[3] |
|---|---|---|---|---|
| 1997 | 4.8 | 9.1 | 3.0 | -0.3 - 8.0 |
| 1998 | 3.9 | 9.5 | 2.7 | -0.7 - 7.3 |
| 1999 | 3.7 | 8.7 | 2.7 | -1.0 - 6.7 |
| 2000 | 4.4 | 8.6 | 3.0 | 0.0 - 8.0 |
| 2001 | 4.6 | 11.0 | 3.0 | -0.7 - 6.7 |
| 2002 | 3.7 | 9.0 | 2.3 | -0.7 - 7.0 |
| 2003 | 4.2 | 9.0 | 2.7 | -0.3 - 7.0 |
| 2004 | 4.6 | 9.0 | 3.3 | 0.3 - 7.3 |
| 2005 | 4.0 | 9.4 | 2.3 | -0.3 - 6.7 |
| 2006 | 4.0 | 8.1 | 2.7 | -0.3 - 7.0 |
| 2007 | 5.3 | 10.3 | 3.3 | 0.0 - 8.3 |
| 2008 | 4.7 | 8.9 | 3.3 | 0.3 - 8.0 |
| 2009 | 4.5 | 10.7 | 2.7 | -0.3 - 7.0 |
| 2010 | 4.7 | 8.7 | 3.0 | -0.3 - 8.3 |
| 2011 | 5.2 | 10.7 | 3.3 | 0.0 - 8.3 |
| 2012 | 4.8 | 10.3 | 3.0 | -0.3 - 7.7 |
| 2013 | 4.8 | 11.1 | 3.0 | 0.0 - 7.3 |
| 2014 | 5.2 | 9.6 | 3.7 | 0.3 - 8.3 |
| 2015 | 5.0 | 11.1 | 3.3 | -0.3 - 8.7 |
| **Overall** | **4.6** | **9.7** | **3.0** | **-0.3 - 7.7** |

[1] The period for which encounters were averaged (as number per year) was 0 to 365 days after diagnosis for AGW; [2] Standard deviation; [3] Q1 and Q3 are the first and third quartile respectively

Supplementary Table 122 **Distribution of excess number of hospital admissions[1] (per year) due to anogenital warts (for patients compared to their matches) among Manitoba males by year of diagnosis (1997-2015).**

| Year of diagnosis | Average | SD[2] | Median | Q1 - Q3[3] |
|---|---|---|---|---|
| 1997 | 0.3 | 0.7 | 0.0 | 0.0 - 0.7 |
| 1998 | 0.3 | 0.8 | 0.0 | 0.0 - 0.7 |
| 1999 | 0.2 | 0.6 | 0.0 | 0.0 - 0.0 |
| 2000 | 0.3 | 0.8 | 0.0 | 0.0 - 0.7 |
| 2001 | 0.3 | 1.2 | 0.0 | 0.0 - 0.0 |
| 2002 | 0.1 | 0.6 | 0.0 | 0.0 - 0.0 |
| 2003 | 0.2 | 0.6 | 0.0 | 0.0 - 0.0 |
| 2004 | 0.2 | 0.7 | 0.0 | 0.0 - 0.0 |
| 2005 | 0.1 | 0.6 | 0.0 | 0.0 - 0.0 |
| 2006 | 0.1 | 0.6 | 0.0 | 0.0 - 0.0 |
| 2007 | 0.1 | 0.7 | 0.0 | 0.0 - 0.0 |
| 2008 | 0.2 | 1.0 | 0.0 | 0.0 - 0.0 |
| 2009 | 0.1 | 0.6 | 0.0 | 0.0 - 0.0 |
| 2010 | 0.1 | 0.5 | 0.0 | 0.0 - 0.0 |
| 2011 | 0.1 | 0.5 | 0.0 | 0.0 - 0.0 |
| 2012 | 0.1 | 0.5 | 0.0 | 0.0 - 0.0 |
| 2013 | 0.1 | 0.5 | 0.0 | 0.0 - 0.0 |
| 2014 | 0.1 | 0.6 | 0.0 | 0.0 - 0.0 |
| 2015 | 0.1 | 0.6 | 0.0 | 0.0 - 0.0 |
| **Overall** | **0.2** | **0.7** | **0.0** | **0.0 - 0.0** |

[1] The period for which encounters were averaged (as number per year) was 0 to 365 days after diagnosis for AGW; [2] Standard deviation; [3] Q1 and Q3 are the first and third quartile respectively

Supplementary Table 123 **Distribution of excess number of ER visits[1,2] (per year) due to anogenital warts (for patients compared to their matches) among Manitoba males by year of diagnosis (1997-2015).**

| Year of diagnosis | Average | SD[3] | Median | Q1 - Q3[4] |
|---|---|---|---|---|
| 1997 | 0.0 | 0.0 | 0.0 | 0.0 - 0.0 |
| 1998 | 0.0 | 0.3 | 0.0 | 0.0 - 0.0 |
| 1999 | 0.0 | 0.9 | 0.0 | -0.3 - 0.0 |
| 2000 | 0.2 | 1.1 | 0.0 | -0.3 - 0.0 |
| 2001 | 0.1 | 1.1 | 0.0 | -0.3 - 0.0 |
| 2002 | 0.1 | 1.1 | 0.0 | -0.3 - 0.0 |
| 2003 | 0.1 | 1.0 | 0.0 | -0.3 - 0.0 |
| 2004 | 0.2 | 2.7 | 0.0 | -0.3 - 0.0 |
| 2005 | 0.2 | 1.3 | 0.0 | -0.3 - 0.0 |
| 2006 | 0.1 | 1.0 | 0.0 | -0.3 - 0.0 |
| 2007 | 0.1 | 1.1 | 0.0 | -0.3 - 0.0 |
| 2008 | 0.1 | 1.3 | 0.0 | -0.3 - 0.0 |
| 2009 | 0.0 | 2.1 | 0.0 | -0.3 - 0.0 |
| 2010 | 0.1 | 1.0 | 0.0 | -0.3 - 0.0 |
| 2011 | 0.1 | 0.9 | 0.0 | -0.3 - 0.0 |
| 2012 | 0.1 | 1.0 | 0.0 | -0.3 - 0.0 |
| 2013 | 0.1 | 1.4 | 0.0 | -0.3 - 0.0 |
| 2014 | 0.2 | 1.0 | 0.0 | 0.0 - 0.0 |
| 2015 | 0.0 | 0.1 | 0.0 | 0.0 - 0.0 |
| **Overall** | **0.1** | **1.2** | **0.0** | **-0.3 - 0.0** |

[1] ER visits are for the Winnipeg Regional Health Authority only; [2] The period for which encounters were averaged (as number per year) was 0 to 365 days after diagnosis for AGW; [3] Standard deviation; [4] Q1 and Q3 are the first and third quartile respectively

Supplementary Table 124 **Distribution of excess number of prescriptions[1] (per year) due to anogenital warts (for patients compared to their matches) among Manitoba males by year of diagnosis (1997-2015).**

| Year of diagnosis | Average | SD[2] | Median | Q1 - Q3[3] |
|---|---|---|---|---|
| 1997 | 3.2 | 16.1 | 0.3 | -1.3 - 3.7 |
| 1998 | 3.0 | 17.1 | 0.3 | -1.7 - 4.0 |
| 1999 | 0.9 | 12.3 | 0.0 | -2.3 - 2.7 |
| 2000 | 2.7 | 20.1 | 0.0 | -2.0 - 4.0 |
| 2001 | 2.9 | 17.0 | 0.0 | -2.0 - 3.0 |
| 2002 | 0.8 | 19.2 | 0.0 | -2.3 - 3.0 |
| 2003 | 3.3 | 23.9 | 0.0 | -2.0 - 3.7 |
| 2004 | 2.6 | 21.4 | 0.3 | -2.0 - 4.0 |
| 2005 | 0.7 | 17.3 | 0.0 | -2.7 - 3.7 |
| 2006 | 0.7 | 20.9 | 0.0 | -2.7 - 4.0 |
| 2007 | 2.7 | 23.3 | 0.3 | -2.0 - 4.0 |
| 2008 | 1.6 | 31.9 | 0.3 | -2.3 - 5.0 |
| 2009 | 2.8 | 35.4 | 0.0 | -2.7 - 4.3 |
| 2010 | 2.8 | 22.9 | 0.3 | -2.7 - 4.3 |
| 2011 | 3.3 | 26.9 | 0.3 | -2.3 - 5.0 |
| 2012 | 2.5 | 32.0 | 0.0 | -2.7 - 4.0 |
| 2013 | 1.3 | 30.5 | 0.3 | -2.7 - 5.0 |
| 2014 | 3.4 | 35.2 | 0.0 | -2.7 - 5.0 |
| 2015 | 3.9 | 35.0 | 0.3 | -3.3 - 5.3 |
| **Overall** | **2.4** | **25.9** | **0.0** | **-2.3 - 4.0** |

[1] The period for which encounters were averaged (as number per year) was 0 to 365 days after diagnosis for AGW; [2] Standard deviation; [3] Q1 and Q3 are the first and third quartile respectively

Supplementary Table 125 **Distribution of excess number of clinician visits[1] (per year) due to HPV-related carcinoma *in situ* (for patients compared to their matches) among Manitoba males by year of diagnosis (1997-2015).**

| Year of diagnosis | Average | SD[2] | Median | Q1 - Q3[3] |
|---|---|---|---|---|
| 1997 | 3.8 | 1.6 | 3.8 | 2.7 - 5.0 |
| 1998 | 12.2 | 8.2 | 13.0 | 3.7 - 20.0 |
| 1999 | 5.2 | 1.6 | 5.2 | 4.0 - 6.3 |
| 2000 | 13.8 | 8.0 | 15.3 | 7.2 - 20.3 |
| 2001 | 8.4 | 18.7 | -2.0 | -2.7 - 30.0 |
| 2002 | 18.8 | 8.5 | 17.5 | 14.0 - 26.3 |
| 2003 | 8.2 | 8.7 | 8.2 | 2.0 - 14.3 |
| 2004 | 4.0 | 8.8 | 3.3 | 0.7 - 9.0 |
| 2005 | 6.7 | 14.0 | 0.3 | -1.0 - 14.3 |
| 2006 | 3.9 | 13.7 | -2.7 | -5.3 - 19.7 |
| 2007 | 1.3 | 4.2 | 2.5 | -1.7 - 4.2 |
| 2008 | 16.7 | 0.0 | 16.7 | 16.7 - 16.7 |
| 2009 | 16.7 | 13.5 | 11.0 | 9.7 - 17.7 |
| 2010 | 6.3 | 17.1 | -0.2 | -5.2 - 17.7 |
| 2011 | 12.3 | 19.8 | 4.0 | 2.0 - 15.7 |
| 2012 | 7.9 | 12.2 | 6.7 | -3.7 - 20.7 |
| 2013 | 7.5 | 9.5 | 7.5 | 0.0 - 17.3 |
| 2014 | 5.2 | 7.5 | 4.7 | 1.0 - 10.7 |
| 2015 | 4.1 | 4.2 | 4.0 | 2.0 - 4.0 |
| **Overall** | **8.6** | **11.1** | **4.8** | **1.2 - 15.8** |

[1] The period for which encounters were averaged (as number per year) was 0 to 365 days after diagnosis for carcinoma *in situ*; [2] Standard deviation; [3] Q1 and Q3 are the first and third quartile respectively

Supplementary Table 126 **Distribution of excess number of hospital admissions[1] (per year) due to HPV-related carcinoma *in situ* (for patients compared to their matches) among Manitoba males by year of diagnosis (1997-2015).**

| Year of diagnosis | Average | SD[2] | Median | Q1 - Q3[3] |
|---|---|---|---|---|
| 1997 | 1.3 | 0.9 | 1.3 | 0.7 - 2.0 |
| 1998 | 1.0 | 0.0 | 1.0 | 1.0 - 1.0 |
| 1999 | 1.2 | 1.2 | 1.2 | 0.3 - 2.0 |
| 2000 | 1.0 | 0.8 | 1.0 | 0.5 - 1.5 |
| 2001 | 1.0 | 0.0 | 1.0 | 1.0 - 1.0 |
| 2002 | 0.8 | 0.8 | 0.8 | 0.0 - 1.7 |
| 2003 | 0.8 | 1.2 | 0.8 | 0.0 - 1.7 |
| 2004 | 0.7 | 1.1 | 1.0 | 0.0 - 1.7 |
| 2005 | 0.3 | 0.9 | 0.2 | -0.5 - 1.0 |
| 2006 | 0.9 | 1.2 | 1.0 | -0.3 - 2.0 |
| 2007 | 0.3 | 0.9 | 0.0 | -0.2 - 0.8 |
| 2008 | 2.5 | 0.7 | 2.5 | 2.0 - 3.0 |
| 2009 | 1.2 | 0.8 | 1.3 | 0.7 - 2.0 |
| 2010 | -0.4 | 1.3 | -0.3 | -1.3 - 0.5 |
| 2011 | 0.8 | 1.3 | 0.3 | 0.0 - 0.7 |
| 2012 | 0.7 | 0.6 | 1.0 | 0.0 - 1.0 |
| 2013 | 0.8 | 1.7 | 0.5 | -0.3 - 1.0 |
| 2014 | 0.6 | 1.4 | 0.0 | -0.3 - 2.0 |
| 2015 | 0.4 | 0.4 | 0.3 | 0.0 - 0.7 |
| **Overall** | **0.8** | **1.0** | **0.7** | **0.0 - 1.3** |

[1] The period for which encounters were averaged (as number per year) was 0 to 365 days after diagnosis for carcinoma *in situ*; [2] Standard deviation; [3] Q1 and Q3 are the first and third quartile respectively

Supplementary Table 127 **Distribution of excess number of ER visits[1,2] (per year) due to HPV-related carcinoma *in situ* (for patients compared to their matches) among Manitoba males by year of diagnosis (1997-2015).**

| Year of diagnosis | Average | SD[3] | Median | Q1 - Q3[4] |
|---|---|---|---|---|
| 1997 | 0.0 | | 0.0 | 0.0 - 0.0 |
| 1998 | 0.0 | 0.0 | 0.0 | 0.0 - 0.0 |
| 1999 | 1.0 | | 1.0 | 1.0 - 1.0 |
| 2000 | 1.3 | 2.3 | 0.0 | 0.0 - 4.0 |
| 2001 | 0.4 | 0.5 | 0.3 | 0.0 - 1.0 |
| 2002 | -0.1 | 0.2 | 0.0 | -0.2 - 0.0 |
| 2003 | 1.7 | | 1.7 | 1.7 - 1.7 |
| 2004 | -0.1 | 1.8 | 0.0 | -0.7 - 0.0 |
| 2005 | -0.4 | 0.2 | -0.3 | -0.7 - -0.3 |
| 2006 | -0.7 | 0.9 | -0.3 | -1.7 - 0.0 |
| 2007 | -0.1 | 0.2 | 0.0 | -0.3 - 0.0 |
| 2008 | 1.3 | 2.8 | 1.3 | -0.7 - 3.3 |
| 2009 | -0.1 | 1.4 | -0.7 | -1.0 - 0.8 |
| 2010 | -0.2 | 0.4 | 0.0 | -0.7 - 0.0 |
| 2011 | -1.7 | 2.9 | -0.3 | -5.0 - 0.3 |
| 2012 | 0.0 | 0.6 | -0.3 | -0.3 - 0.7 |
| 2013 | -0.8 | 1.5 | -1.0 | -2.0 - 0.5 |
| 2014 | 0.0 | 1.2 | -0.3 | -0.7 - 0.0 |
| 2015 | 0.0 | 0.0 | 0.0 | 0.0 - 0.0 |
| **Overall** | **-0.0** | **1.3** | **0.0** | **-0.3 - 0.0** |

[1] ER visits are for the Winnipeg Regional Health Authority only; [2] The period for which encounters were averaged (as number per year) was 0 to 365 days after diagnosis for carcinoma *in situ*; [3] Standard deviation; [4] Q1 and Q3 are the first and third quartile respectively

Supplementary Table 128 **Distribution of excess number of prescriptions[1] (per year) due to HPV-related carcinoma *in situ* (for patients compared to their matches) among Manitoba males by year of diagnosis (1997-2015).**

| Year of diagnosis | Average | SD[2] | Median | Q1 - Q3[3] |
|---|---|---|---|---|
| 1997 | 4.8 | 0.7 | 4.8 | 4.3 - 5.3 |
| 1998 | 36.4 | 47.1 | 41.7 | -13.0 - 80.7 |
| 1999 | 1.8 | 1.6 | 1.8 | 0.7 - 3.0 |
| 2000 | 8.6 | 8.7 | 6.3 | 2.3 - 14.8 |
| 2001 | 23.9 | 57.7 | -1.7 | -16.7 - 90.0 |
| 2002 | 11.6 | 31.2 | -0.7 | -10.7 - 27.7 |
| 2003 | 43.0 | 70.2 | 43.0 | -6.7 - 92.7 |
| 2004 | 11.0 | 57.5 | -1.0 | -21.0 - 5.7 |
| 2005 | -3.6 | 20.3 | -3.2 | -20.8 - 13.7 |
| 2006 | -2.2 | 12.2 | 4.3 | -16.3 - 5.3 |
| 2007 | 18.5 | 34.8 | 7.0 | -1.3 - 38.3 |
| 2008 | 76.7 | 96.6 | 76.7 | 8.3 - 145.0 |
| 2009 | -3.2 | 40.4 | 7.3 | -39.0 - 25.7 |
| 2010 | -48.3 | 98.6 | -1.3 | -99.7 - 3.2 |
| 2011 | 30.2 | 71.5 | 8.3 | 2.3 - 29.7 |
| 2012 | -21.4 | 18.3 | -31.3 | -32.7 - -0.3 |
| 2013 | 18.5 | 69.8 | 8.0 | -10.7 - 22.0 |
| 2014 | -1.0 | 14.9 | -2.7 | -15.3 - 15.7 |
| 2015 | 20.4 | 53.0 | -0.7 | -5.7 - -0.3 |
| **Overall** | **9.6** | **49.6** | **1.8** | **-10.0 - 14.2** |

[1] The period for which encounters were averaged (as number per year) was 0 to 365 days after diagnosis for carcinoma *in situ*; [2] Standard deviation; [3] Q1 and Q3 are the first and third quartile respectively

Supplementary Table 129 **Distribution of excess number of clinician visits[1] (per year) due to HPV-related invasive cancer (for patients compared to their matches) among Manitoba males by year of diagnosis (1997-2015).**

| Year of diagnosis | Average | SD[2] | Median | Q1 - Q3[3] |
|---|---|---|---|---|
| 1997 | 7.5 | 10.5 | 6.8 | 1.3 - 10.6 |
| 1998 | 9.0 | 16.4 | 7.7 | 1.6 - 16.8 |
| 1999 | 8.9 | 12.1 | 7.3 | 0.7 - 14.0 |
| 2000 | 8.1 | 10.7 | 6.7 | 2.9 - 15.1 |
| 2001 | 8.5 | 15.1 | 10.0 | 2.3 - 19.3 |
| 2002 | 13.9 | 11.7 | 10.1 | 6.2 - 18.3 |
| 2003 | 11.2 | 12.6 | 9.9 | 4.5 - 16.6 |
| 2004 | 13.3 | 18.0 | 10.7 | 5.9 - 22.7 |
| 2005 | 11.5 | 14.2 | 10.4 | 5.9 - 18.7 |
| 2006 | 11.8 | 14.7 | 9.3 | 3.1 - 18.1 |
| 2007 | 9.2 | 13.9 | 7.7 | 3.1 - 15.7 |
| 2008 | 12.0 | 17.3 | 9.1 | 2.9 - 17.1 |
| 2009 | 10.7 | 13.9 | 8.5 | 5.2 - 13.8 |
| 2010 | 9.5 | 10.6 | 8.7 | 1.7 - 14.6 |
| 2011 | 14.5 | 23.3 | 10.9 | 4.4 - 17.9 |
| 2012 | 12.5 | 12.6 | 8.0 | 2.9 - 18.4 |
| 2013 | 9.6 | 16.7 | 9.0 | 2.3 - 17.2 |
| 2014 | 8.7 | 17.1 | 8.2 | 0.5 - 16.8 |
| 2015 | 4.7 | 16.0 | 6.1 | -0.3 - 11.5 |
| **Overall** | **10.2** | **15.4** | **8.8** | **3.0 - 16.7** |

[1] The periods for which encounters were averaged (as number per year) was 180 days before to 730 days after diagnosis for invasive cancer; [2] Standard deviation; [3] Q1 and Q3 are the first and third quartile respectively

Supplementary Table 130 **Distribution of excess number of hospital admissions[1] (per year) due to HPV-related invasive cancer (for patients compared to their matches) among Manitoba males by year of diagnosis (1997-2015).**

| Year of diagnosis | Average | SD[2] | Median | Q1 - Q3[3] |
|---|---|---|---|---|
| 1997 | 1.1 | 1.1 | 0.8 | 0.4 - 1.7 |
| 1998 | 1.4 | 2.2 | 1.0 | 0.1 - 2.3 |
| 1999 | 1.2 | 1.0 | 1.0 | 0.4 - 1.9 |
| 2000 | 1.0 | 1.3 | 0.7 | 0.3 - 1.4 |
| 2001 | 1.2 | 1.1 | 0.9 | 0.5 - 1.6 |
| 2002 | 1.6 | 1.3 | 1.2 | 0.8 - 2.1 |
| 2003 | 0.9 | 1.2 | 0.7 | 0.3 - 1.3 |
| 2004 | 1.2 | 1.4 | 0.9 | 0.3 - 1.7 |
| 2005 | 1.1 | 1.0 | 0.8 | 0.4 - 1.9 |
| 2006 | 0.9 | 1.0 | 0.8 | 0.3 - 1.6 |
| 2007 | 0.9 | 1.3 | 0.8 | 0.4 - 1.5 |
| 2008 | 1.2 | 1.2 | 0.8 | 0.4 - 2.0 |
| 2009 | 1.1 | 1.4 | 0.8 | 0.4 - 1.3 |
| 2010 | 0.9 | 0.8 | 0.8 | 0.4 - 1.6 |
| 2011 | 1.0 | 0.9 | 0.7 | 0.4 - 1.5 |
| 2012 | 1.0 | 1.1 | 0.8 | 0.3 - 1.5 |
| 2013 | 0.8 | 1.1 | 0.8 | 0.3 - 1.2 |
| 2014 | 0.9 | 1.0 | 0.5 | 0.1 - 1.3 |
| 2015 | 0.7 | 1.1 | 0.4 | 0.1 - 1.0 |
| **Overall** | **1.0** | **1.2** | **0.8** | **0.4 - 1.6** |

[1] The periods for which encounters were averaged (as number per year) was 180 days before to 730 days after diagnosis for invasive cancer; [2] Standard deviation; [3] Q1 and Q3 are the first and third quartile respectively

Supplementary Table 131 **Distribution of excess number of ER visits[1,2] (per year) due to HPV-related invasive cancer (for patients compared to their matches) among Manitoba males by year of diagnosis (1997-2015).**

| Year of diagnosis | Average | SD[3] | Median | Q1 - Q3[4] |
|---|---|---|---|---|
| 1997 | 0.0 | 0.1 | 0.0 | 0.0 - 0.0 |
| 1998 | 0.3 | 1.1 | 0.0 | -0.1 - 0.3 |
| 1999 | 0.7 | 2.2 | 0.0 | -0.1 - 0.8 |
| 2000 | 0.0 | 0.9 | -0.1 | -0.4 - 0.3 |
| 2001 | 0.3 | 0.9 | 0.0 | -0.1 - 1.0 |
| 2002 | 0.8 | 1.7 | 0.4 | -0.1 - 1.0 |
| 2003 | 0.3 | 0.7 | 0.1 | -0.1 - 0.4 |
| 2004 | 1.7 | 6.8 | 0.3 | -0.1 - 1.0 |
| 2005 | 0.3 | 0.9 | 0.1 | 0.0 - 1.1 |
| 2006 | 0.4 | 1.1 | 0.1 | -0.3 - 0.5 |
| 2007 | 0.1 | 1.1 | 0.0 | -0.3 - 0.6 |
| 2008 | 0.1 | 1.0 | 0.3 | -0.4 - 0.5 |
| 2009 | 1.0 | 1.9 | 0.4 | 0.0 - 1.2 |
| 2010 | 0.3 | 1.1 | 0.1 | -0.1 - 0.5 |
| 2011 | 0.7 | 1.5 | 0.3 | 0.0 - 0.7 |
| 2012 | 0.7 | 1.3 | 0.1 | -0.1 - 1.2 |
| 2013 | 1.2 | 3.8 | 0.1 | -0.3 - 0.8 |
| 2014 | 0.1 | 0.6 | 0.0 | -0.1 - 0.4 |
| 2015 | -0.0 | 0.3 | 0.0 | 0.0 - 0.0 |
| **Overall** | **0.5** | **2.1** | **0.0** | **-0.1 - 0.5** |

[1] ER visits are for the Winnipeg Regional Health Authority only; [2] The periods for which encounters were averaged (as number per year) was 180 days before to 730 days after diagnosis for invasive cancer; [3] Standard deviation; [4] Q1 and Q3 are the first and third quartile respectively

Supplementary Table 132 **Distribution of excess number of prescriptions[1] (per year) due to HPV-related invasive cancer (for patients compared to their matches) among Manitoba males by year of diagnosis (1997-2015).**

| Year of diagnosis | Average | SD[2] | Median | Q1 - Q3[3] |
|---|---|---|---|---|
| 1997 | 11.2 | 29.2 | 6.9 | 0.2 - 16.7 |
| 1998 | 12.8 | 22.7 | 9.1 | 1.2 - 23.2 |
| 1999 | 15.0 | 39.4 | 7.3 | -2.8 - 27.9 |
| 2000 | 13.0 | 27.5 | 7.2 | -2.7 - 24.2 |
| 2001 | 13.3 | 32.7 | 11.6 | -0.7 - 22.8 |
| 2002 | 20.2 | 38.1 | 6.7 | -0.5 - 47.6 |
| 2003 | 12.6 | 23.9 | 7.7 | -4.8 - 26.7 |
| 2004 | 13.8 | 45.4 | 9.3 | -11.5 - 31.9 |
| 2005 | 5.5 | 30.6 | 6.0 | -10.4 - 28.9 |
| 2006 | 4.5 | 30.9 | 2.3 | -7.5 - 21.2 |
| 2007 | -13.4 | 91.1 | -2.8 | -27.1 - 13.5 |
| 2008 | 9.4 | 87.7 | 0.3 | -18.9 - 28.3 |
| 2009 | 1.9 | 71.4 | 0.8 | -13.7 - 16.9 |
| 2010 | 10.0 | 44.6 | 3.3 | -7.9 - 16.8 |
| 2011 | 22.3 | 100.7 | 4.3 | -6.8 - 33.1 |
| 2012 | 9.6 | 116.4 | 17.6 | -0.9 - 40.1 |
| 2013 | 33.9 | 221.9 | 9.3 | -3.9 - 25.6 |
| 2014 | 32.0 | 164.1 | 7.0 | -10.1 - 22.7 |
| 2015 | -0.2 | 49.3 | -0.2 | -12.9 - 12.9 |
| **Overall** | **12.2** | **91.5** | **6.0** | **-7.9 - 24.6** |

[1] The periods for which encounters were averaged (as number per year) was 180 days before to 730 days after diagnosis for invasive cancer; [2] Standard deviation; [3] Q1 and Q3 are the first and third quartile respectively

Supplementary Table 133 **Distribution of excess number of clinician visits[1] (per year) due to invasive cancer of the anus (for patients compared to their matches) among Manitoba males by year of diagnosis (1997-2015).**

| Year of diagnosis | Average | SD[2] | Median | Q1 - Q3[3] |
|---|---|---|---|---|
| 1997 | 7.1 | | 7.1 | 7.1 - 7.1 |
| 1998 | 12.6 | 7.5 | 14.8 | 4.3 - 18.7 |
| 1999 | 14.7 | 15.3 | 14.0 | -0.2 - 30.3 |
| 2000 | 9.6 | | 9.6 | 9.6 - 9.6 |
| 2002 | 11.9 | 8.1 | 11.9 | 6.2 - 17.7 |
| 2003 | 10.5 | | 10.5 | 10.5 - 10.5 |
| 2004 | 32.7 | 32.9 | 25.5 | 19.1 - 29.9 |
| 2005 | 23.6 | 11.1 | 23.6 | 15.7 - 31.5 |
| 2006 | 10.2 | 1.0 | 10.0 | 9.3 - 11.3 |
| 2007 | 21.4 | 19.5 | 13.5 | 6.7 - 38.8 |
| 2008 | 21.4 | 16.3 | 15.1 | 9.5 - 34.4 |
| 2009 | 25.8 | 29.2 | 14.9 | 7.6 - 44.0 |
| 2010 | 16.5 | 12.5 | 13.0 | 8.3 - 24.7 |
| 2011 | 8.9 | 7.9 | 10.9 | 4.3 - 13.5 |
| 2012 | 0.9 | 2.5 | 0.9 | -0.8 - 2.7 |
| 2013 | 22.9 | 7.3 | 18.0 | 17.2 - 31.2 |
| 2014 | 1.3 | 50.6 | 21.7 | -12.8 - 28.4 |
| 2015 | 9.1 | 4.1 | 8.1 | 5.2 - 12.7 |
| **Overall** | **16.2** | **20.6** | **14.4** | **6.9 - 23.6** |

[1] The periods for which encounters were averaged (as number per year) was 180 days before to 730 days after diagnosis for invasive cancer; [2] Standard deviation; [3] Q1 and Q3 are the first and third quartile respectively

Supplementary Table 134 **Distribution of excess number of hospital admissions[1] (per year) due to invasive cancer of the anus (for patients compared to their matches) among Manitoba males by year of diagnosis (1997-2015).**

| Year of diagnosis | Average | SD[2] | Median | Q1 - Q3[3] |
|---|---|---|---|---|
| 1997 | 2.0 | | 2.0 | 2.0 - 2.0 |
| 1998 | 1.9 | 1.7 | 2.4 | 0.0 - 3.2 |
| 1999 | 1.3 | 0.4 | 1.2 | 1.0 - 1.8 |
| 2000 | 1.6 | | 1.6 | 1.6 - 1.6 |
| 2002 | 2.6 | 0.4 | 2.6 | 2.4 - 2.9 |
| 2003 | 1.6 | | 1.6 | 1.6 - 1.6 |
| 2004 | 2.6 | 2.8 | 1.6 | 1.3 - 2.4 |
| 2005 | 1.4 | 0.8 | 1.4 | 0.8 - 2.0 |
| 2006 | 0.9 | 0.1 | 0.9 | 0.8 - 1.1 |
| 2007 | 1.8 | 1.7 | 1.6 | 0.4 - 3.2 |
| 2008 | 1.8 | 1.0 | 2.3 | 0.8 - 2.5 |
| 2009 | 3.2 | 3.4 | 2.0 | 1.1 - 5.4 |
| 2010 | 1.5 | 0.9 | 1.4 | 0.7 - 2.2 |
| 2011 | 0.7 | 0.7 | 0.6 | 0.3 - 1.1 |
| 2012 | 0.6 | 0.5 | 0.6 | 0.3 - 0.9 |
| 2013 | 2.1 | 1.4 | 1.5 | 1.1 - 3.6 |
| 2014 | 1.7 | 1.0 | 2.0 | 1.3 - 2.1 |
| 2015 | 1.3 | 2.0 | 0.7 | -0.1 - 1.7 |
| **Overall** | **1.8** | **1.6** | **1.4** | **0.7 - 2.4** |

[1] The periods for which encounters were averaged (as number per year) was 180 days before to 730 days after diagnosis for invasive cancer; [2] Standard deviation; [3] Q1 and Q3 are the first and third quartile respectively

Supplementary Table 135 **Distribution of excess number of ER visits[1,2] (per year) due to invasive cancer of the anus (for patients compared to their matches) among Manitoba males by year of diagnosis (1997-2015).**

| Year of diagnosis | Average | SD[3] | Median | Q1 - Q3[4] |
|---|---|---|---|---|
| 1997 | 0.3 | | 0.3 | 0.3 - 0.3 |
| 1998 | 1.2 | 1.2 | 1.2 | 0.0 - 2.4 |
| 1999 | 10.2 | | 10.2 | 10.2 - 10.2 |
| 2000 | -0.3 | | -0.3 | -0.3 - -0.3 |
| 2002 | 0.0 | | 0.0 | 0.0 - 0.0 |
| 2003 | 0.8 | | 0.8 | 0.8 - 0.8 |
| 2004 | 10.3 | 18.7 | 1.4 | 0.7 - 20.0 |
| 2005 | 1.5 | | 1.5 | 1.5 - 1.5 |
| 2006 | 0.0 | 0.5 | 0.3 | -0.5 - 0.4 |
| 2007 | 0.7 | 1.1 | 0.1 | 0.1 - 1.2 |
| 2008 | 0.6 | 1.0 | 0.6 | -0.1 - 1.2 |
| 2009 | 3.1 | 4.1 | 1.5 | 0.7 - 5.5 |
| 2010 | 0.6 | 0.5 | 0.4 | 0.3 - 1.2 |
| 2011 | 0.9 | 1.5 | 0.4 | -0.3 - 2.7 |
| 2012 | 0.0 | 0.8 | 0.0 | -0.5 - 0.5 |
| 2013 | 5.4 | 6.9 | 2.6 | 1.0 - 9.8 |
| 2014 | 0.3 | 0.9 | 0.0 | -0.4 - 1.2 |
| 2015 | 0.0 | 0.0 | 0.0 | 0.0 - 0.0 |
| **Overall** | **2.2** | **6.2** | **0.4** | **0.0 - 1.5** |

[1] ER visits are for the Winnipeg Regional Health Authority only; [2] The periods for which encounters were averaged (as number per year) was 180 days before to 730 days after diagnosis for invasive cancer; [3] Standard deviation; [4] Q1 and Q3 are the first and third quartile respectively

Supplementary Table 136 **Distribution of excess number of prescriptions[1] (per year) due to invasive cancer of the anus (for patients compared to their matches) among Manitoba males by year of diagnosis (1997-2015).**

| Year of diagnosis | Average | SD[2] | Median | Q1 - Q3[3] |
|---|---|---|---|---|
| 1997 | 2.7 | | 2.7 | 2.7 - 2.7 |
| 1998 | 8.8 | 12.7 | 5.1 | -1.6 - 22.9 |
| 1999 | 78.5 | 85.0 | 32.1 | 26.8 - 176.7 |
| 2000 | 65.5 | | 65.5 | 65.5 - 65.5 |
| 2002 | 37.4 | 14.5 | 37.4 | 27.1 - 47.6 |
| 2003 | 7.2 | | 7.2 | 7.2 - 7.2 |
| 2004 | 77.6 | 76.8 | 66.1 | 18.0 - 130.8 |
| 2005 | 20.0 | 36.8 | 20.0 | -6.0 - 46.0 |
| 2006 | 4.1 | 25.9 | -0.3 | -19.3 - 31.9 |
| 2007 | 29.9 | 83.4 | -8.1 | -23.1 - 72.8 |
| 2008 | 29.8 | 45.9 | 9.6 | 5.1 - 27.2 |
| 2009 | 16.4 | 66.2 | -14.2 | -18.1 - 50.9 |
| 2010 | 70.5 | 98.4 | 29.7 | 13.7 - 127.3 |
| 2011 | 13.8 | 30.8 | 4.9 | -6.9 - 34.5 |
| 2012 | 100.0 | 128.4 | 100.0 | 9.2 - 190.8 |
| 2013 | 257.4 | 647.2 | 9.2 | 3.2 - 33.2 |
| 2014 | 304.1 | 544.9 | 11.6 | 8.8 - 353.4 |
| 2015 | 0.3 | 9.5 | 5.1 | -10.9 - 7.5 |
| **Overall** | **79.8** | **266.5** | **9.2** | **-1.5 - 46.8** |

[1] The periods for which encounters were averaged (as number per year) was 180 days before to 730 days after diagnosis for invasive cancer; [2] Standard deviation; [3] Q1 and Q3 are the first and third quartile respectively

Supplementary Table 137 **Distribution of excess number of clinician visits[1] (per year) due to invasive cancer of the oral cavity (for patients compared to their matches) among Manitoba males by year of diagnosis (1997-2015).**

| Year of diagnosis | Average | SD[2] | Median | Q1 - Q3[3] |
|---|---|---|---|---|
| 1997 | 5.4 | 11.0 | 6.8 | -0.7 - 8.9 |
| 1998 | 8.6 | 12.7 | 3.1 | 0.3 - 10.8 |
| 1999 | 7.9 | 9.9 | 6.3 | 1.1 - 13.8 |
| 2000 | 5.2 | 9.1 | 3.5 | -0.4 - 6.9 |
| 2001 | 6.9 | 12.0 | 4.0 | -1.5 - 17.0 |
| 2002 | 11.9 | 7.8 | 10.8 | 6.3 - 16.8 |
| 2003 | 12.5 | 10.8 | 11.5 | 4.3 - 21.1 |
| 2004 | 13.6 | 9.2 | 10.7 | 7.9 - 19.2 |
| 2005 | 11.0 | 17.5 | 9.5 | 6.4 - 18.3 |
| 2006 | 10.7 | 14.7 | 8.7 | 2.7 - 22.9 |
| 2007 | 7.6 | 17.2 | 8.0 | -2.5 - 16.3 |
| 2008 | 14.5 | 22.6 | 8.4 | 2.5 - 16.3 |
| 2009 | 8.1 | 13.2 | 8.0 | 5.3 - 11.5 |
| 2010 | 9.3 | 12.0 | 8.5 | 0.3 - 15.2 |
| 2011 | 16.1 | 34.1 | 12.2 | 3.1 - 17.6 |
| 2012 | 15.1 | 14.5 | 10.1 | 6.1 - 20.5 |
| 2013 | 12.8 | 14.4 | 9.2 | 2.9 - 15.3 |
| 2014 | 7.2 | 14.0 | 2.7 | -0.8 - 8.1 |
| 2015 | 5.5 | 17.0 | 7.9 | -0.8 - 13.3 |
| **Overall** | **10.1** | **16.2** | **8.1** | **2.1 - 15.5** |

[1] The periods for which encounters were averaged (as number per year) was 180 days before to 730 days after diagnosis for invasive cancer; [2] Standard deviation; [3] Q1 and Q3 are the first and third quartile respectively

Supplementary Table 138 **Distribution of excess number of hospital admissions[1] (per year) due to invasive cancer of the oral cavity (for patients compared to their matches) among Manitoba males by year of diagnosis (1997-2015).**

| Year of diagnosis | Average | SD[2] | Median | Q1 - Q3[3] |
|---|---|---|---|---|
| 1997 | 0.8 | 0.9 | 0.8 | 0.5 - 1.6 |
| 1998 | 1.4 | 2.8 | 0.8 | 0.1 - 1.5 |
| 1999 | 1.1 | 1.0 | 0.9 | 0.4 - 2.0 |
| 2000 | 0.4 | 0.9 | 0.3 | 0.1 - 0.6 |
| 2001 | 1.2 | 1.0 | 0.9 | 0.4 - 1.6 |
| 2002 | 1.3 | 0.7 | 1.1 | 0.7 - 1.7 |
| 2003 | 1.2 | 0.8 | 1.1 | 0.5 - 1.5 |
| 2004 | 1.3 | 1.4 | 1.0 | 0.4 - 2.0 |
| 2005 | 1.2 | 1.2 | 0.8 | 0.4 - 2.2 |
| 2006 | 1.1 | 0.9 | 0.9 | 0.4 - 1.6 |
| 2007 | 0.6 | 1.3 | 0.7 | 0.3 - 1.4 |
| 2008 | 1.2 | 1.0 | 0.9 | 0.4 - 2.1 |
| 2009 | 0.6 | 0.8 | 0.5 | 0.3 - 1.1 |
| 2010 | 0.9 | 0.8 | 0.8 | 0.4 - 1.6 |
| 2011 | 0.9 | 0.8 | 0.8 | 0.4 - 1.4 |
| 2012 | 1.1 | 1.2 | 0.8 | 0.4 - 1.2 |
| 2013 | 0.8 | 0.7 | 0.8 | 0.3 - 1.3 |
| 2014 | 1.0 | 0.8 | 0.8 | 0.4 - 1.3 |
| 2015 | 0.7 | 0.8 | 0.4 | 0.3 - 0.8 |
| **Overall** | **1.0** | **1.1** | **0.8** | **0.4 - 1.6** |

[1] The periods for which encounters were averaged (as number per year) was 180 days before to 730 days after diagnosis for invasive cancer; [2] Standard deviation; [3] Q1 and Q3 are the first and third quartile respectively

Supplementary Table 139 **Distribution of excess number of ER visits[1,2] (per year) due to invasive cancer of the oral cavity (for patients compared to their matches) among Manitoba males by year of diagnosis (1997-2015).**

| Year of diagnosis | Average | SD[3] | Median | Q1 - Q3[4] |
|---|---|---|---|---|
| 1997 | 0.0 | 0.1 | 0.0 | 0.0 - 0.0 |
| 1998 | 0.4 | 1.5 | 0.0 | -0.1 - 0.2 |
| 1999 | 0.4 | 1.3 | 0.0 | -0.1 - 0.8 |
| 2000 | -0.1 | 0.8 | -0.1 | -0.5 - -0.1 |
| 2001 | 0.5 | 1.1 | 0.3 | -0.1 - 1.1 |
| 2002 | 0.1 | 0.8 | 0.3 | -0.6 - 0.8 |
| 2003 | 0.1 | 0.4 | 0.1 | -0.1 - 0.3 |
| 2004 | 0.8 | 2.1 | 0.3 | 0.0 - 1.0 |
| 2005 | 0.5 | 0.7 | 0.3 | 0.0 - 1.2 |
| 2006 | 0.3 | 0.8 | 0.1 | -0.1 - 0.5 |
| 2007 | -0.0 | 0.5 | -0.1 | -0.4 - 0.1 |
| 2008 | -0.1 | 0.8 | 0.3 | -0.3 - 0.4 |
| 2009 | 0.4 | 1.0 | 0.4 | -0.4 - 1.1 |
| 2010 | 0.5 | 1.2 | 0.0 | 0.0 - 0.4 |
| 2011 | 0.3 | 1.1 | 0.3 | -0.1 - 0.7 |
| 2012 | 0.5 | 1.2 | 0.0 | -0.1 - 1.2 |
| 2013 | 1.5 | 4.7 | 0.0 | -0.5 - 1.1 |
| 2014 | -0.0 | 0.9 | 0.0 | -0.1 - 0.4 |
| 2015 | -0.1 | 0.5 | 0.0 | 0.0 - 0.0 |
| **Overall** | **0.3** | **1.4** | **0.0** | **-0.1 - 0.5** |

[1] ER visits are for the Winnipeg Regional Health Authority only; [2] The periods for which encounters were averaged (as number per year) was 180 days before to 730 days after diagnosis for invasive cancer; [3] Standard deviation; [4] Q1 and Q3 are the first and third quartile respectively

Supplementary Table 140 **Distribution of excess number of prescriptions[1] (per year) due to invasive cancer of the oral cavity (for patients compared to their matches) among Manitoba males by year of diagnosis (1997-2015).**

| Year of diagnosis | Average | SD[2] | Median | Q1 - Q3[3] |
|---|---|---|---|---|
| 1997 | 10.1 | 39.9 | 4.7 | -8.9 - 15.2 |
| 1998 | 10.7 | 26.4 | 3.2 | -4.1 - 15.6 |
| 1999 | 14.8 | 30.6 | 6.7 | -4.9 - 27.9 |
| 2000 | 17.9 | 35.4 | 5.2 | -2.7 - 26.3 |
| 2001 | 22.3 | 31.3 | 14.9 | -0.7 - 29.6 |
| 2002 | 13.9 | 33.9 | 4.2 | -11.7 - 33.5 |
| 2003 | 12.4 | 22.8 | 9.9 | -3.9 - 27.5 |
| 2004 | 12.6 | 31.3 | 10.1 | -8.5 - 31.9 |
| 2005 | 1.1 | 29.5 | 3.1 | -16.7 - 22.3 |
| 2006 | 9.0 | 26.7 | 8.0 | -6.7 - 21.2 |
| 2007 | -33.0 | 135.9 | -0.8 | -21.2 - 7.1 |
| 2008 | 41.8 | 100.7 | 12.1 | -14.0 - 57.1 |
| 2009 | 21.7 | 70.1 | 9.7 | -12.9 - 21.9 |
| 2010 | 2.2 | 36.5 | -0.4 | -7.9 - 12.8 |
| 2011 | 30.3 | 155.6 | 11.9 | -1.7 - 33.1 |
| 2012 | 21.9 | 31.8 | 20.6 | 2.8 - 33.4 |
| 2013 | 17.6 | 36.6 | 13.6 | 0.1 - 29.1 |
| 2014 | 2.0 | 56.3 | 2.4 | -38.0 - 16.5 |
| 2015 | -4.4 | 63.3 | 11.5 | -19.2 - 24.4 |
| **Overall** | **11.9** | **67.3** | **6.8** | **-7.9 - 25.0** |

[1] The periods for which encounters were averaged (as number per year) was 180 days before to 730 days after diagnosis for invasive cancer; [2] Standard deviation; [3] Q1 and Q3 are the first and third quartile respectively

Supplementary Table 141 **Distribution of excess number of clinician visits[1] (per year) due to invasive cancer of the oropharynx (for patients compared to their matches) among Manitoba males by year of diagnosis (1997-2015).**

| Year of diagnosis | Average | SD[2] | Median | Q1 - Q3[3] |
|---|---|---|---|---|
| 1997 | 6.9 | 7.4 | 6.1 | 1.6 - 10.5 |
| 1998 | 8.5 | 21.9 | 11.8 | 4.6 - 21.2 |
| 1999 | 13.7 | 14.5 | 9.9 | 4.3 - 21.0 |
| 2000 | 12.6 | 11.0 | 14.4 | 10.3 - 18.1 |
| 2001 | 9.3 | 19.0 | 11.8 | 6.0 - 20.8 |
| 2002 | 15.1 | 14.5 | 10.0 | 6.8 - 20.9 |
| 2003 | 10.2 | 12.8 | 9.9 | 6.3 - 15.4 |
| 2004 | 10.0 | 18.7 | 10.1 | 5.8 - 23.6 |
| 2005 | 12.8 | 9.2 | 11.9 | 6.4 - 19.1 |
| 2006 | 11.9 | 16.4 | 7.0 | 3.2 - 17.9 |
| 2007 | 9.1 | 9.6 | 11.2 | 3.7 - 13.3 |
| 2008 | 11.0 | 14.1 | 11.5 | 3.6 - 18.5 |
| 2009 | 11.0 | 9.6 | 9.5 | 5.2 - 22.3 |
| 2010 | 9.6 | 8.2 | 8.8 | 4.5 - 13.6 |
| 2011 | 13.5 | 15.0 | 9.3 | 4.8 - 17.9 |
| 2012 | 11.5 | 11.1 | 7.3 | 2.3 - 19.7 |
| 2013 | 7.1 | 18.0 | 5.7 | 1.5 - 13.5 |
| 2014 | 9.0 | 10.8 | 8.3 | 3.6 - 14.8 |
| 2015 | 5.1 | 11.8 | 5.2 | 0.1 - 10.7 |
| **Overall** | **10.2** | **13.8** | **9.3** | **3.7 - 17.5** |

[1] The periods for which encounters were averaged (as number per year) was 180 days before to 730 days after diagnosis for invasive cancer; [2] Standard deviation; [3] Q1 and Q3 are the first and third quartile respectively

Supplementary Table 142 **Distribution of excess number of hospital admissions[1] (per year) due to invasive cancer of the oropharynx (for patients compared to their matches) among Manitoba males by year of diagnosis (1997-2015).**

| Year of diagnosis | Average | SD[2] | Median | Q1 - Q3[3] |
|---|---|---|---|---|
| 1997 | 1.0 | 1.1 | 0.7 | 0.4 - 1.2 |
| 1998 | 1.4 | 1.4 | 1.3 | 0.8 - 2.3 |
| 1999 | 1.4 | 1.2 | 1.4 | 0.3 - 2.0 |
| 2000 | 1.4 | 1.7 | 0.8 | 0.5 - 1.6 |
| 2001 | 1.2 | 1.4 | 1.2 | 0.4 - 2.4 |
| 2002 | 1.7 | 1.6 | 1.2 | 0.8 - 2.1 |
| 2003 | 0.7 | 1.3 | 0.6 | 0.3 - 1.2 |
| 2004 | 0.8 | 0.9 | 0.6 | 0.2 - 1.4 |
| 2005 | 1.0 | 0.9 | 0.8 | 0.4 - 1.7 |
| 2006 | 0.8 | 1.1 | 0.4 | 0.1 - 1.3 |
| 2007 | 0.9 | 1.2 | 0.8 | 0.4 - 1.4 |
| 2008 | 1.2 | 1.6 | 0.8 | 0.1 - 1.7 |
| 2009 | 1.0 | 0.9 | 0.8 | 0.5 - 1.2 |
| 2010 | 0.8 | 0.9 | 0.7 | 0.4 - 1.2 |
| 2011 | 1.0 | 1.0 | 0.4 | 0.4 - 1.5 |
| 2012 | 0.9 | 1.1 | 0.7 | 0.3 - 1.7 |
| 2013 | 0.7 | 1.1 | 0.4 | 0.0 - 1.2 |
| 2014 | 0.4 | 0.6 | 0.4 | -0.1 - 0.4 |
| 2015 | 0.5 | 1.0 | 0.4 | 0.0 - 0.8 |
| **Overall** | **0.9** | **1.2** | **0.7** | **0.3 - 1.4** |

[1] The periods for which encounters were averaged (as number per year) was 180 days before to 730 days after diagnosis for invasive cancer; [2] Standard deviation; [3] Q1 and Q3 are the first and third quartile respectively

Supplementary Table 143 **Distribution of excess number of ER visits[1,2] (per year) due to invasive cancer of the oropharynx (for patients compared to their matches) among Manitoba males by year of diagnosis (1997-2015).**

| Year of diagnosis | Average | SD[3] | Median | Q1 - Q3[4] |
|---|---|---|---|---|
| 1997 | -0.1 | 0.1 | 0.0 | -0.1 - 0.0 |
| 1998 | 0.1 | 0.2 | 0.0 | 0.0 - 0.1 |
| 1999 | 0.1 | 0.5 | 0.0 | 0.0 - 0.4 |
| 2000 | 0.2 | 1.2 | 0.3 | -0.7 - 0.3 |
| 2001 | 0.2 | 0.8 | 0.0 | -0.4 - 0.7 |
| 2002 | 1.4 | 2.1 | 0.8 | 0.4 - 1.7 |
| 2003 | 0.4 | 1.0 | 0.1 | -0.1 - 0.5 |
| 2004 | 0.3 | 1.7 | 0.3 | -0.4 - 0.4 |
| 2005 | -0.0 | 1.0 | 0.0 | 0.0 - 0.5 |
| 2006 | 0.5 | 1.6 | 0.0 | -0.4 - 0.5 |
| 2007 | 0.2 | 1.4 | 0.1 | -0.1 - 1.3 |
| 2008 | 0.3 | 1.0 | 0.3 | -0.4 - 0.5 |
| 2009 | 0.8 | 1.5 | 0.3 | 0.0 - 0.8 |
| 2010 | 0.2 | 1.1 | 0.1 | -0.3 - 0.8 |
| 2011 | 0.8 | 1.7 | 0.2 | 0.0 - 0.5 |
| 2012 | 1.2 | 1.6 | 0.4 | 0.0 - 2.8 |
| 2013 | -0.1 | 0.5 | -0.1 | -0.4 - 0.1 |
| 2014 | 0.1 | 0.6 | 0.0 | -0.1 - 0.4 |
| 2015 | 0.0 | 0.1 | 0.0 | 0.0 - 0.0 |
| **Overall** | **0.4** | **1.2** | **0.0** | **-0.1 - 0.4** |

[1] ER visits are for the Winnipeg Regional Health Authority only; [2] The periods for which encounters were averaged (as number per year) was 180 days before to 730 days after diagnosis for invasive cancer; [3] Standard deviation; [4] Q1 and Q3 are the first and third quartile respectively

Supplementary Table 144 **Distribution of excess number of prescriptions[1] (per year) due to invasive cancer of the oropharynx (for patients compared to their matches) among Manitoba males by year of diagnosis (1997-2015).**

| Year of diagnosis | Average | SD[2] | Median | Q1 - Q3[3] |
|---|---|---|---|---|
| 1997 | 10.0 | 11.2 | 9.9 | 0.9 - 12.5 |
| 1998 | 14.6 | 20.1 | 19.9 | 7.6 - 27.8 |
| 1999 | 11.9 | 37.2 | 14.8 | 1.6 - 29.2 |
| 2000 | 8.8 | 14.6 | 7.2 | -4.8 - 16.1 |
| 2001 | 9.3 | 36.1 | 11.1 | 0.5 - 16.1 |
| 2002 | 21.9 | 43.4 | 6.7 | 2.9 - 30.4 |
| 2003 | 8.8 | 23.3 | 3.2 | -9.2 - 16.5 |
| 2004 | 1.3 | 38.8 | 4.9 | -10.3 - 19.4 |
| 2005 | 8.5 | 32.5 | 13.2 | 3.3 - 30.9 |
| 2006 | 4.3 | 30.3 | 2.4 | -7.1 - 21.0 |
| 2007 | 0.5 | 42.6 | 0.4 | -20.5 - 15.5 |
| 2008 | -21.1 | 90.3 | -7.2 | -20.5 - 11.9 |
| 2009 | -17.0 | 72.7 | 0.4 | -12.5 - 13.0 |
| 2010 | 7.6 | 33.0 | 3.2 | -9.2 - 14.1 |
| 2011 | 17.0 | 57.0 | 0.7 | -6.8 - 27.1 |
| 2012 | -12.8 | 165.8 | 15.3 | -0.9 - 45.1 |
| 2013 | -4.5 | 49.4 | 1.1 | -12.3 - 18.5 |
| 2014 | 14.2 | 56.1 | 7.6 | -8.4 - 21.7 |
| 2015 | 0.4 | 21.5 | -0.4 | -7.6 - 8.3 |
| **Overall** | **4.1** | **58.5** | **4.9** | **-7.1 - 20.9** |

[1] The periods for which encounters were averaged (as number per year) was 180 days before to 730 days after diagnosis for invasive cancer; [2] Standard deviation; [3] Q1 and Q3 are the first and third quartile respectively

Supplementary Table 145 **Distribution of excess number of clinician visits[1] (per year) due to invasive cancer of the penis (for patients compared to their matches) among Manitoba males by year of diagnosis (1997-2015).**

| Year of diagnosis | Average | SD[2] | Median | Q1 - Q3[3] |
|---|---|---|---|---|
| 1997 | 29.6 | 3.9 | 29.6 | 26.8 - 32.4 |
| 1998 | 12.0 | | 12.0 | 12.0 - 12.0 |
| 1999 | 0.1 | 7.2 | 0.7 | -9.3 - 6.8 |
| 2000 | 3.2 | 11.2 | 5.1 | 2.9 - 8.7 |
| 2001 | 9.9 | 10.4 | 6.9 | 5.1 - 18.5 |
| 2002 | 16.3 | 12.0 | 13.7 | 7.6 - 25.0 |
| 2003 | 11.4 | 18.4 | 4.7 | 1.3 - 32.2 |
| 2004 | 5.9 | 6.8 | 5.9 | 3.2 - 6.3 |
| 2005 | -1.3 | 14.2 | 2.5 | -17.0 - 10.6 |
| 2006 | 19.3 | 7.8 | 18.3 | 12.1 - 27.6 |
| 2007 | 2.5 | 4.9 | 4.2 | -2.5 - 6.0 |
| 2008 | 5.0 | 7.3 | 5.7 | -2.1 - 12.8 |
| 2009 | -0.3 | 19.2 | -0.3 | -13.9 - 13.2 |
| 2010 | -3.3 | 7.2 | -3.3 | -8.4 - 1.7 |
| 2011 | 18.0 | 20.8 | 7.1 | 4.4 - 23.7 |
| 2012 | 9.3 | 10.8 | 8.0 | 3.7 - 13.5 |
| 2013 | 1.9 | 16.1 | 6.9 | -0.3 - 9.3 |
| 2014 | 12.0 | 12.4 | 12.7 | 6.5 - 16.8 |
| 2015 | 0.9 | 23.2 | 3.5 | -6.7 - 11.0 |
| **Overall** | **6.8** | **14.7** | **6.5** | **0.7 - 12.9** |

[1] The periods for which encounters were averaged (as number per year) was 180 days before to 730 days after diagnosis for invasive cancer; [2] Standard deviation; [3] Q1 and Q3 are the first and third quartile respectively

Supplementary Table 146 **Distribution of excess number of hospital admissions[1] (per year) due to invasive cancer of the penis (for patients compared to their matches) among Manitoba males by year of diagnosis (1997-2015).**

| Year of diagnosis | Average | SD[2] | Median | Q1 - Q3[3] |
|---|---|---|---|---|
| 1997 | 3.1 | 0.3 | 3.1 | 2.9 - 3.3 |
| 1998 | 0.8 | | 0.8 | 0.8 - 0.8 |
| 1999 | 0.9 | 0.6 | 0.7 | 0.4 - 1.6 |
| 2000 | 1.4 | 1.0 | 1.2 | 0.7 - 1.7 |
| 2001 | 1.0 | 0.5 | 0.8 | 0.7 - 1.2 |
| 2002 | 2.1 | 1.0 | 2.3 | 1.3 - 2.9 |
| 2003 | 0.7 | 1.6 | 0.0 | 0.0 - 0.3 |
| 2004 | 1.2 | 1.0 | 0.9 | 0.5 - 1.2 |
| 2005 | 0.1 | 1.3 | 0.8 | -1.4 - 0.9 |
| 2006 | 0.5 | 0.6 | 0.1 | 0.1 - 1.2 |
| 2007 | 0.8 | 0.5 | 0.7 | 0.5 - 1.1 |
| 2008 | 0.8 | 0.5 | 0.7 | 0.5 - 0.8 |
| 2009 | 2.0 | 0.6 | 2.0 | 1.6 - 2.4 |
| 2010 | 0.9 | 0.8 | 0.9 | 0.4 - 1.5 |
| 2011 | 1.2 | 0.8 | 0.8 | 0.7 - 1.6 |
| 2012 | 0.7 | 0.8 | 0.4 | 0.4 - 1.5 |
| 2013 | 0.5 | 0.6 | 0.7 | 0.3 - 0.8 |
| 2014 | 1.7 | 1.3 | 1.3 | 0.8 - 2.7 |
| 2015 | 0.9 | 1.1 | 0.5 | 0.0 - 1.9 |
| **Overall** | **1.0** | **1.0** | **0.8** | **0.4 - 1.6** |

[1] The periods for which encounters were averaged (as number per year) was 180 days before to 730 days after diagnosis for invasive cancer; [2] Standard deviation; [3] Q1 and Q3 are the first and third quartile respectively

Supplementary Table 147 **Distribution of excess number of ER visits[1,2] (per year) due to invasive cancer of the penis (for patients compared to their matches) among Manitoba males by year of diagnosis (1997-2015).**

| Year of diagnosis | Average | SD[3] | Median | Q1 - Q3[4] |
|---|---|---|---|---|
| 1997 | 0.0 | | 0.0 | 0.0 - 0.0 |
| 1998 | 0.3 | | 0.3 | 0.3 - 0.3 |
| 1999 | 0.4 | 1.0 | 0.1 | -0.3 - 1.0 |
| 2000 | 0.0 | 0.3 | 0.1 | -0.2 - 0.3 |
| 2001 | 0.2 | 0.2 | 0.3 | 0.0 - 0.3 |
| 2002 | 0.7 | 1.8 | 0.7 | -0.5 - 2.0 |
| 2003 | | | | |
| 2004 | 0.3 | 0.4 | 0.4 | -0.1 - 0.5 |
| 2005 | -0.6 | | -0.6 | -0.6 - -0.6 |
| 2006 | 0.3 | 0.3 | 0.3 | 0.1 - 0.5 |
| 2007 | -0.5 | 0.4 | -0.7 | -0.8 - -0.0 |
| 2008 | -0.3 | 1.1 | 0.0 | -0.8 - 0.5 |
| 2009 | 1.4 | | 1.4 | 1.4 - 1.4 |
| 2010 | -0.9 | | -0.9 | -0.9 - -0.9 |
| 2011 | 1.9 | 2.7 | 1.9 | 0.0 - 3.8 |
| 2012 | 0.0 | | 0.0 | 0.0 - 0.0 |
| 2013 | 0.2 | 0.6 | 0.4 | -0.1 - 0.7 |
| 2014 | 0.4 | 0.4 | 0.3 | 0.0 - 0.7 |
| 2015 | -0.0 | 0.1 | 0.0 | 0.0 - 0.0 |
| **Overall** | **0.2** | **0.8** | **0.0** | **-0.1 - 0.4** |

[1] ER visits are for the Winnipeg Regional Health Authority only; [2] The periods for which encounters were averaged (as number per year) was 180 days before to 730 days after diagnosis for invasive cancer; [3] Standard deviation; [4] Q1 and Q3 are the first and third quartile respectively

Supplementary Table 148 **invasive cancer of the penis Distribution of excess number of prescriptions[1] (per year) due to invasive cancer of the penis (for patients compared to their matches) among Manitoba males by year of diagnosis (1997-2015).**

| Year of diagnosis | Average | SD[2] | Median | Q1 - Q3[3] |
|---|---|---|---|---|
| 1997 | 33.9 | 23.1 | 33.9 | 17.6 - 50.2 |
| 1998 | 35.6 | | 35.6 | 35.6 - 35.6 |
| 1999 | -5.9 | 7.9 | -2.8 | -12.8 - 1.7 |
| 2000 | 4.7 | 26.1 | 3.9 | -2.7 - 28.4 |
| 2001 | 5.4 | 24.9 | 1.6 | -6.5 - 24.8 |
| 2002 | 27.8 | 37.9 | 30.3 | -4.3 - 60.0 |
| 2003 | 26.7 | 29.4 | 27.7 | -4.1 - 46.7 |
| 2004 | 3.9 | 27.8 | -11.5 | -13.6 - 10.2 |
| 2005 | 5.2 | 36.6 | -14.1 | -17.7 - 47.5 |
| 2006 | -24.3 | 63.3 | 1.2 | -96.4 - 22.3 |
| 2007 | -56.1 | 66.2 | -28.7 | -99.1 - -10.4 |
| 2008 | -3.7 | 42.6 | -8.0 | -32.1 - 19.2 |
| 2009 | 30.5 | 46.4 | 30.5 | -2.4 - 63.3 |
| 2010 | 23.5 | 83.4 | 23.5 | -35.5 - 82.5 |
| 2011 | 28.9 | 39.1 | 32.0 | 4.0 - 44.6 |
| 2012 | 19.7 | 85.1 | -19.6 | -29.2 - 107.5 |
| 2013 | 21.5 | 75.6 | 12.5 | -3.9 - 25.6 |
| 2014 | 6.7 | 38.1 | 6.6 | -19.6 - 26.4 |
| 2015 | 6.2 | 69.9 | -9.1 | -13.4 - 8.7 |
| **Overall** | **6.3** | **50.6** | **1.2** | **-15.1 - 28.0** |

[1] The periods for which encounters were averaged (as number per year) was 180 days before to 730 days after diagnosis for invasive cancer; [2] Standard deviation; [3] Q1 and Q3 are the first and third quartile respectively

#### Attributable excess healthcare utilization

Supplementary Table 149 **Excess number of healthcare encounters[1] (per year) due to HPV-related diseases (for patients compared to their matches) among Manitoba males according to different estimates of the attributable fraction (1997-2015)**

| HPV-related disease | Total healthcare encounters | Volesky et al.[3] | | Saraiya et al.[2] | |
|---|---|---|---|---|---|
| | | AF[2] | Attributable visits | AF[2] | Attributable visits |
| **Excess clinician visits** | | | | | |
| Anogenital warts | 67,637 | 100% | 67,637 | 100% | 67,637 |
| All HPV-related carcinoma *in situ* | 733 | [3] | 250 | [4] | 424 |
| All HPV-related invasive cancers | 11,665 | | 4,876 | | 6,823 |
| Invasive cancer of the anus | 1,167 | 88% | 1,027 | 89% | 1,039 |
| Invasive cancer of the oral cavity | 4,330 | 8% | 346 | 33% | 1,429 |
| Invasive cancer of the oropharynx | 5,224 | 60% | 3,134 | 72% | 3,761 |
| Invasive cancer of the penis | 943 | 39% | 368 | 63% | 594 |
| **Total** | **80,034** | | **72,763** | | **74,885** |
| **Excess hospital admissions** | | | | | |
| Anogenital warts | 2,218 | 100% | 2,218 | 100% | 2,218 |
| All HPV-related carcinoma *in situ* | 66 | [3] | 21 | [4] | 36 |
| All HPV-related invasive cancers | 1,174 | | 489 | | 688 |
| Invasive cancer of the anus | 126 | 88% | 111 | 89% | 113 |
| Invasive cancer of the oral cavity | 425 | 8% | 34 | 33% | 140 |
| Invasive cancer of the oropharynx | 478 | 60% | 287 | 72% | 344 |
| Invasive cancer of the penis | 145 | 39% | 56 | 63% | 91 |
| **Total** | **3,458** | | **2,728** | | **2,943** |
| **Excess ER visits[5]** | | | | | |
| Anogenital warts | 1,118 | 100% | 1,118 | 100% | 1,118 |
| All HPV-related carcinoma *in situ* | -3 | [3] | -4 | [4] | -4 |
| All HPV-related invasive cancers | 319 | | 174 | | 213 |

| | | | | | |
|---|---|---|---|---|---|
| Invasive cancer of the anus | 107 | 88% | 95 | 89% | 96 |
| Invasive cancer of the oral cavity | 88 | 8% | 7 | 33% | 29 |
| Invasive cancer of the oropharynx | 113 | 60% | 68 | 72% | 81 |
| Invasive cancer of the penis | 11 | 39% | 4 | 63% | 7 |
| **Total** | **1,434** | | **1,287** | | **1,327** |
| **Excess prescriptions** | | | | | |
| Anogenital warts | 35,618 | 100% | 35,618 | 100% | 35,618 |
| All HPV-related carcinoma *in situ* | 811 | [3] | 297 | [4] | 479 |
| All HPV-related invasive cancers | 13,811 | | 7,057 | | 8,851 |
| Invasive cancer of the anus | 5,747 | 88% | 5,057 | 89% | 5,115 |
| Invasive cancer of the oral cavity | 5,107 | 8% | 409 | 33% | 1,685 |
| Invasive cancer of the oropharynx | 2,089 | 60% | 1,253 | 72% | 1,504 |
| Invasive cancer of the penis | 868 | 39% | 338 | 63% | 547 |
| **Total** | **50,239** | | **42,973** | | **44,947** |

[1] The periods for which encounters were averaged (as number per year) were 0 to 365 days after diagnosis for AGW and carcinoma *in situ* and 180 days before to 730 days after diagnosis for invasive cancer; [2] AF = Fraction of disease attributable to HPV; [3] Carcinoma *in situ* AF: 88% for anus, 8% for oral cavity, 60% for oropharynx, and 39% for penis; [4] Carcinoma *in situ* AF: 89% for anus, 33% for oral cavity, 72% for oropharynx, and 63% for penis; [5] ER visits are for the Winnipeg Regional Health Authority only

Supplementary Table 150 **Excess number of clinician visits[1] (per year) due to HPV-related diseases (for patients compared to their matches) among Manitoba males by year of diagnosis according to different estimates of the attributable fraction**

| Year | Volesky et al.[3] | | Saraiya et al.[2] | |
|---|---|---|---|---|
| | Total | Per case | Total | Per case |
| 1997 | 3,870 | 4.73 | 3,942 | 4.81 |
| 1998 | 2,710 | 3.87 | 2,798 | 4.00 |
| 1999 | 2,339 | 3.69 | 2,406 | 3.79 |
| 2000 | 2,935 | 4.32 | 3,020 | 4.44 |
| 2001 | 2,945 | 4.48 | 3,043 | 4.62 |
| 2002 | 2,931 | 3.90 | 3,080 | 4.10 |
| 2003 | 3,233 | 4.20 | 3,337 | 4.33 |
| 2004 | 3,843 | 4.68 | 3,959 | 4.82 |
| 2005 | 3,425 | 4.00 | 3,519 | 4.11 |
| 2006 | 3,382 | 4.06 | 3,497 | 4.19 |
| 2007 | 4,185 | 5.21 | 4,275 | 5.32 |
| 2008 | 4,239 | 4.72 | 4,381 | 4.87 |
| 2009 | 4,096 | 4.50 | 4,217 | 4.63 |
| 2010 | 4,613 | 4.66 | 4,725 | 4.78 |
| 2011 | 5,220 | 5.20 | 5,426 | 5.41 |
| 2012 | 4,758 | 4.76 | 4,909 | 4.91 |
| 2013 | 4,323 | 4.80 | 4,424 | 4.91 |
| 2014 | 4,968 | 5.04 | 5,090 | 5.16 |
| 2015 | 4,749 | 4.73 | 4,838 | 4.82 |
| **Total** | **72,763** | **4.54** | **74,885** | **4.67** |

[1] The periods for which encounters were averaged (as number per year) were 0 to 365 days after diagnosis for AGW and carcinoma *in situ* and 180 days before to 730 days after diagnosis for invasive cancer

Supplementary Table 151 **Excess number of hospital admissions[1] (per year) due to HPV-related diseases (for patients compared to their matches) among Manitoba males by year of diagnosis according to different estimates of the attributable fraction**

| Year | Volesky et al.[3] | | Saraiya et al.[2] | |
|---|---|---|---|---|
| | Total | Per case | Total | Per case |
| 1997 | 220 | 0.27 | 230 | 0.28 |
| 1998 | 217 | 0.31 | 230 | 0.33 |
| 1999 | 144 | 0.23 | 154 | 0.24 |
| 2000 | 190 | 0.28 | 199 | 0.29 |
| 2001 | 177 | 0.27 | 189 | 0.29 |
| 2002 | 125 | 0.17 | 140 | 0.19 |
| 2003 | 154 | 0.20 | 163 | 0.21 |
| 2004 | 150 | 0.18 | 162 | 0.20 |
| 2005 | 133 | 0.16 | 143 | 0.17 |
| 2006 | 90 | 0.11 | 100 | 0.12 |
| 2007 | 115 | 0.14 | 125 | 0.16 |
| 2008 | 178 | 0.20 | 192 | 0.21 |
| 2009 | 122 | 0.13 | 133 | 0.15 |
| 2010 | 114 | 0.12 | 125 | 0.13 |
| 2011 | 128 | 0.13 | 141 | 0.14 |
| 2012 | 99 | 0.10 | 110 | 0.11 |
| 2013 | 113 | 0.13 | 122 | 0.14 |
| 2014 | 142 | 0.14 | 155 | 0.16 |
| 2015 | 117 | 0.12 | 130 | 0.13 |
| **Total** | **2,728** | **0.17** | **2,943** | **0.18** |

[1] The periods for which encounters were averaged (as number per year) were 0 to 365 days after diagnosis for AGW and carcinoma *in situ* and 180 days before to 730 days after diagnosis for invasive cancer

Supplementary Table 152 **Excess number of ER visits[1,2] (per year) due to HPV-related diseases (for patients compared to their matches) among Manitoba males by year of diagnosis according to different estimates of the attributable fraction**

| Year | Volesky et al.[3] | | Saraiya et al.[2] | |
|---|---|---|---|---|
| | Total | Per case | Total | Per case |
| 1997 | 1 | 0.00 | 2 | 0.00 |
| 1998 | 21 | 0.04 | 23 | 0.04 |
| 1999 | 22 | 0.05 | 24 | 0.05 |
| 2000 | 94 | 0.20 | 96 | 0.20 |
| 2001 | 48 | 0.10 | 50 | 0.11 |
| 2002 | 88 | 0.16 | 91 | 0.17 |
| 2003 | 78 | 0.14 | 80 | 0.14 |
| 2004 | 161 | 0.28 | 166 | 0.29 |
| 2005 | 112 | 0.19 | 114 | 0.19 |
| 2006 | 60 | 0.10 | 62 | 0.10 |
| 2007 | 65 | 0.11 | 65 | 0.11 |
| 2008 | 90 | 0.14 | 91 | 0.14 |
| 2009 | 47 | 0.08 | 51 | 0.08 |
| 2010 | 51 | 0.08 | 54 | 0.08 |
| 2011 | 80 | 0.12 | 83 | 0.12 |
| 2012 | 71 | 0.11 | 76 | 0.11 |
| 2013 | 60 | 0.10 | 64 | 0.11 |
| 2014 | 130 | 0.19 | 131 | 0.19 |
| 2015 | 4 | 0.01 | 4 | 0.01 |
| **Total** | **1,287** | **0.11** | **1,327** | **0.12** |

[1] ER visits are for the Winnipeg Regional Health Authority only; [2] The periods for which encounters were averaged (as number per year) were 0 to 365 days after diagnosis for AGW and carcinoma *in situ* and 180 days before to 730 days after diagnosis for invasive cancer

Supplementary Table 153 **Excess number of prescriptions[1] (per year) due to HPV-related diseases (for patients compared to their matches) among Manitoba males by year of diagnosis according to different estimates of the attributable fraction**

| Year | **Volesky et al.**[3] | | **Saraiya et al.**[2] | |
|---|---|---|---|---|
| | **Total** | **Per case** | **Total** | **Per case** |
| 1997 | 2,706 | 3.30 | 2,807 | 3.43 |
| 1998 | 2,139 | 3.06 | 2,273 | 3.25 |
| 1999 | 855 | 1.35 | 942 | 1.49 |
| 2000 | 1,961 | 2.88 | 2,087 | 3.07 |
| 2001 | 2,000 | 3.04 | 2,170 | 3.30 |
| 2002 | 1,119 | 1.49 | 1,299 | 1.73 |
| 2003 | 2,634 | 3.42 | 2,772 | 3.60 |
| 2004 | 2,376 | 2.89 | 2,481 | 3.02 |
| 2005 | 823 | 0.96 | 856 | 1.00 |
| 2006 | 659 | 0.79 | 706 | 0.85 |
| 2007 | 2,041 | 2.54 | 1,836 | 2.29 |
| 2008 | 1,321 | 1.47 | 1,527 | 1.70 |
| 2009 | 2,255 | 2.48 | 2,338 | 2.57 |
| 2010 | 2,863 | 2.89 | 2,880 | 2.91 |
| 2011 | 3,696 | 3.68 | 4,035 | 4.02 |
| 2012 | 2,378 | 2.38 | 2,492 | 2.49 |
| 2013 | 2,766 | 3.07 | 2,916 | 3.24 |
| 2014 | 4,804 | 4.87 | 4,917 | 4.99 |
| 2015 | 3,574 | 3.56 | 3,612 | 3.60 |
| **Total** | **42,973** | **2.68** | **44,947** | **2.81** |

[1] The periods for which encounters were averaged (as number per year) were 0 to 365 days after diagnosis for AGW and carcinoma *in situ* and 180 days before to 730 days after diagnosis for invasive cancer
